# Sepsis Subphenotyping Based on Organ Dysfunction Trajectory

**DOI:** 10.1101/2021.06.14.21258918

**Authors:** Zhenxing Xu, Chengsheng Mao, Chang Su, Hao Zhang, Ilias Siempos, Lisa K Torres, Di Pan, Yuan Luo, Edward J Schenck, Fei Wang

## Abstract

**Background:** Sepsis is a heterogeneous syndrome, and the identification of clinical subphenotypes is essential. Although organ dysfunction is a defining element of sepsis, subphenotypes of differential trajectory are not well studied. We sought to identify distinct Sequential Organ Failure Assessment (SOFA) score trajectory-based subphenotypes in sepsis.

**Methods:** We created 72-hour SOFA score trajectories in patients with sepsis from four diverse intensive care unit (ICU) cohorts. We then used Dynamic Time Warping (DTW) to compute heterogeneous SOFA trajectory similarities and hierarchical agglomerative clustering (HAC) to identify trajectory-based subphenotypes. Patient characteristics were compared between subphenotypes and a random forest model was developed to predict subphenotype membership at 6 and 24 hours after being admitted to the ICU. The model was tested on three validation cohorts. Sensitivity analyses were performed with alternative clustering methodologies.

**Results:** A total of 4678, 3665, 12282, and 4804 unique sepsis patients were included in development and three validation cohorts, respectively. Four subphenotypes were identified in the development cohort: Rapidly Worsening (n=612, 13.1%), Delayed Worsening (n=960, 20.5%), Rapidly Improving (n=1932, 41.3%) and Delayed Improving (n=1174, 25.1%). Baseline characteristics, including the pattern of organ dysfunction varied between subphenotypes. Rapidly Worsening was defined by a higher comorbidity burden, acidosis, and visceral organ dysfunction. Rapidly Improving was defined by vasopressor use without acidosis. Outcomes differed across the subphenotypes, Rapidly Worsening had the highest in-hospital mortality (28.3%, p-value<0.001), despite a lower SOFA (mean: 4.5) at ICU admission compared to Rapidly Improving (mortality:5.5%, mean SOFA: 5.5). An overall prediction accuracy of 0.78 (95% CI, [0.77, 0.8]) was obtained at 6 hours after ICU admission, which increased to 0.87 (95% CI, [0.86, 0.88]) at 24 hours. Similar subphenotypes were replicated in three validation cohorts. The majority of patients with sepsis have an improving phenotype with a lower mortality risk, however they make up over 20% of all deaths due to their larger numbers.

**Conclusions:** Four novel, clinically-defined, trajectory-based sepsis subphenotypes were identified and validated. Identifying trajectory-based subphenotypes has immediate implications for the powering and predictive enrichment of clinical trials. Understanding the pathophysiology of these differential trajectories may reveal unanticipated therapeutic targets and identify more precise populations and endpoints for clinical trials.

## Introduction

Sepsis is defined as a dysregulated immunological response to infection that results in acute organ dysfunction.^1,2^ The morbidity and mortality of sepsis remain high despite decades of research and numerous failed clinical trials.^3,4^ Recent research has highlighted that sepsis is a complex and heterogeneous syndrome, which includes a multidimensional array of clinical and biological features.^5^ Identifying rigorous sepsis subphenotypes that present with similar prognostic markers and pathophysiologic features has the potential to improve therapy.^6-9^

Recent sepsis subphenotyping studies used static measurements available soon after admission to the emergency department or intensive care unit (ICU) to characterize patients.^5,10-12^ However, due to the stochastic nature of infection and variable presentation to health care after developing symptoms, static assessments of sepsis subphenotypes may be incomplete, ignoring the dynamic nature of organ failure in sepsis.^13^

More recently, subphenotypes characterized by dynamic patient temperature trajectories have been identified in sepsis. The differential pattern of temperature change may represent a varied underlying inflammatory response to infection.^1^ The trajectory of the Sequential Organ Failure Assessment (SOFA) score after ICU admission have been used to predict outcomes and improve prognostic stratification in sepsis.^13,14^ In a recent study, Sanchez-Pinto et al.^15^ leveraged a matrix factorization based approach to identify multiple organ dysfunction syndrome subphenotypes according to longitudinal pediatric SOFA (pSOFA) scores, but their approach was focusing on the subphenotypes captured by the “motifs”, or frequent subsequence patterns, of the SOFA trajectories, which may not characterize the long term trends encoded in those trajectories well. However, whether the trajectory of multisystem organ failure is associated with distinct phenotypic patterns in sepsis remains largely unexplored. Identifying distinct subphenotypes of organ dysfunction trajectory in sepsis can refine our understanding of the natural history of sepsis in the ICU in response to standard of care treatment and define patterns of disease that may benefit from novel therapeutic strategies.^16^

The objective of this study was to develop and evaluate sepsis subphenotypes. The first goal was to determine whether distinct SOFA score trajectory-based subphenotypes in patients with sepsis can be identified through the electronic health record. The second goal was to understand whether those different subphenotypes are associated with the patterns of biomarkers and clinical outcomes. The third goal was to determine whether the identified subphenotypes can be predicted by using patient baseline characteristics and early-stage clinical features.

## Methods

### Overview

We did a cohort study on datasets that contained granular patient level data from a total of 221 hospitals in the United States, whose overall workflow is illustrated in Figure 1. Our goal was to derive sepsis subphenotypes of patients in ICU according to their SOFA organ dysfunction trajectories using dynamic time warping (DTW)^17^ and hierarchical agglomerative clustering (HAC)^18^. We then characterized these subphenotypes using comprehensive patient information including demographics, comorbidities, use of mechanical ventilation, type of ICU unit, admission source, organ source of sepsis, and examined their associated clinical outcomes as well as clinical biomarkers. We further built multiple random forest models to predict the derived subphenotypes from different time points’ patient clinical characteristics. To ensure replicability, the same analysis pipeline was conducted in three validation cohorts.

**Figure 1.**
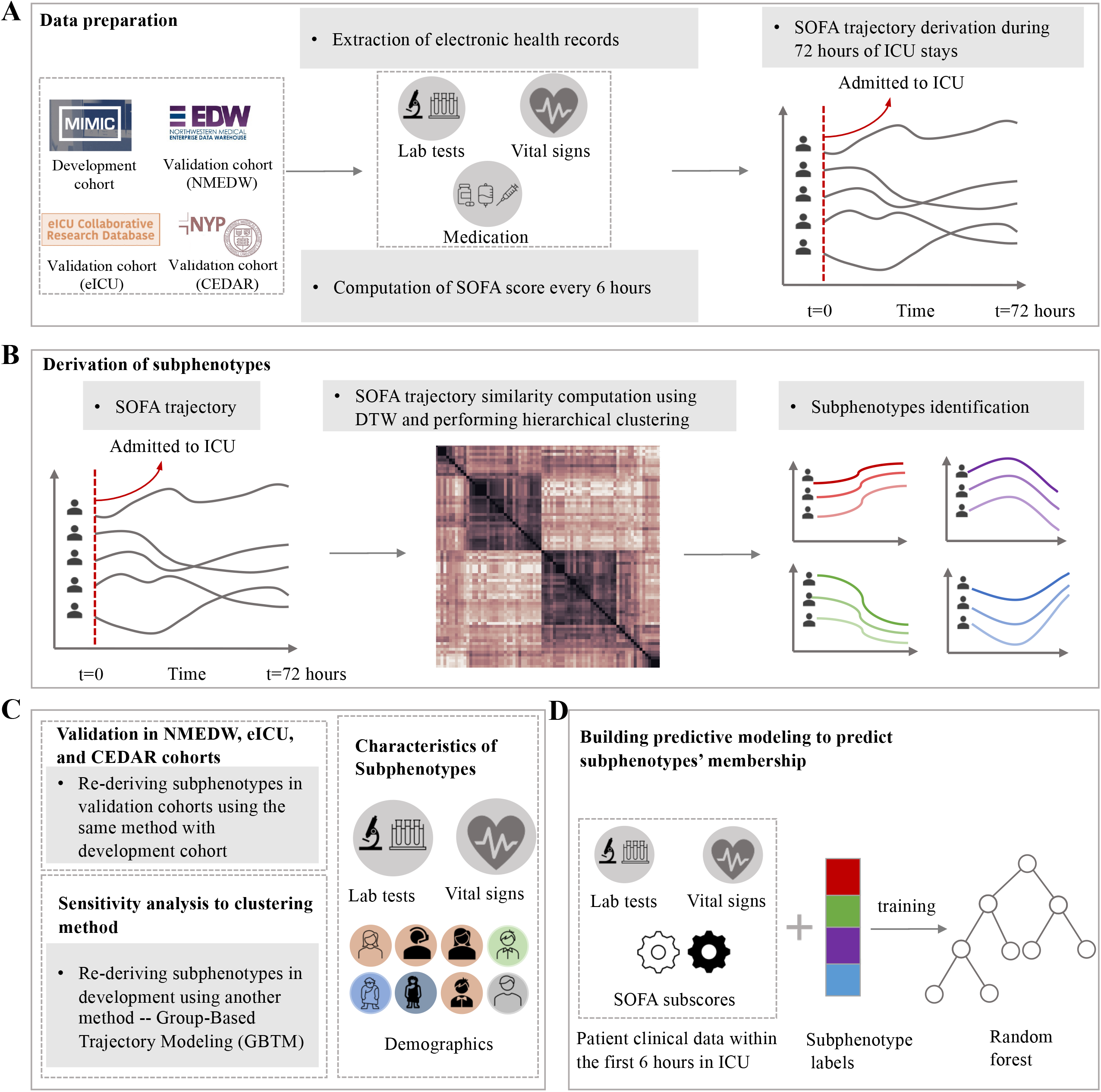
Workflow of study. (A) The MIMIC-III dataset was used as development cohort and NMEDW, eICU, and CEDAR datasets were used as validation cohorts. Electronic health records including lab tests, vital signs, and medication were extracted to compute the SOFA score every 6 hours during 72 hours after admission to ICU. (B) Each patient was represented as a 72-hour SOFA score trajectory. Dynamic Time Warping (DTW) was used to compute heterogeneous SOFA trajectory similarities and HAC was applied to identify subphenotypes based on trajectory similarities. (C) To re-derive subphenotypes in three validation cohorts and consider sensitivity analysis to clustering method, specifically, use another method (Group-Based Trajectory Modeling, GBTM) to generate subphenoytpes. Statistical analysis were performed among subphenotypes in terms of demographic factors, lab tests and vital signs. (D) The predictive model of subphenotypes at successive time points (hours 6, 24, 36, 48, 60) after ICU admission was constructed based on a random forest classifier by using patients’ clinical data including lab tests, vital signs, and SOFA subscores.

### Definition of sepsis and study population

The development cohort (Medical Information Mart for Intensive Care III database: MIMIC-III) was from Beth Israel Deaconess Medical Center (BIDMC) with admissions dating from 2001-2012. which has 673 licensed beds, including 493 medical/surgical beds, 77 critical care beds, and 62 OB/GYN beds.^19^ The first validation cohort was from Northwestern Medicine Enterprise Data Warehouse (NMEDW), which is a network of eleven hospitals located in northern Illinois with 2,554 beds in total, with ICU admissions dating from 2012-2019.^20^ The second validation cohort was from the eICU collaborative research database, which combined multi-center data from patients who were admitted to one of 335 units at 208 hospitals located throughout the US between 2014 and 2015.^21^ The third validation cohort was from Weill Cornell Critical carE Database for Advanced Research (CEDAR) with ICU admissions dating from 2001-2020, which was built on NewYork-Presbyterian/Weill Cornell Medical Center (NYP/WCMC), including 862 beds in total.^22^ The inclusion-exclusion cascade for the patients are shown in Supplemental Figure S1, where Sepsis-3 criteria are defined as in Singer et al.^2^

### SOFA score computation and model descriptions

The SOFA score was derived from six organ-specific subscores including respiration, coagulation, liver, cardiovascular, CNS, renal^16^, which was obtained every 6 hours within the first 72 hours of ICU admission. For each 6-hour period, the worst variable value was used to compute the SOFA subscores. To obtain the urine output during 6 hours, we divided daily urine output by 4. The lowest GCS for each 6 hour period was used irrespective of sedation. Missing values (Supplemental Table S14) were imputed using last observation carried forward (LOCF) and next observation carried backward (NOCB).^23^ If there was no any value during the first 72 hours, we used 0 to fill.

After the SOFA scores were derived, each patient is represented as a vector of 12 SOFA scores from the first 6 hours to the last 6 hours across the 72 hours period after ICU admission. Then, DTW and HAC were used to derive subphenotypes.^17^ In particular, DTW was used to evaluate the similarities between pairwise patient SOFA trajectories (Supplemental Figure S19 and S20). This method can capture the differences among the evolution heterogeneity in terms of the temporal curves, which can assess similarity between patients robustly. HAC was then used to perform clustering among patients based on the similarities obtained from DTW. Multiple clustering indices (Supplemental Appendix 7) were calculated to determine the optimal numbers of subphenotypes.

### Subphenotype reproducibility and prediction

To ensure the robustness of the derived subphenotypes, we re-derived them with group-based trajectory modeling (GBTM), which is one type of latent class analysis (LCA) that assigns each patient a probability of belonging to each particular subphenotype on the basis of maximum likelihood estimation.^24^

We trained a random forest model to predict the derived subphenotypes from the baseline patient clinical collected characteristics at successive time points after ICU admission, with the goal of examining whether the trajectory subphenotypes could be predicted early. Candidate predictors included demographics, comorbidities, SOFA subscores, lab tests, and vital signs. Predictor contributions were evaluated with the Shapley additive explanations (SHAP) strategy.^25^

### Statistical Analysis

Data were analyzed using tslearn package 0.3.1 and scikit-learn package 0.22.2 with Python 3.7. Survival analysis to 28 days was performed using Kaplan-Meier curves. Statistical significance was set at p < 0.05, and all tests were 2-tailed. The detailed descriptions about statistical testing are shown in Supplemental Appendix 2.

## Results

### Cohort characteristics

Our development cohort MIMIC-III had 4,678 sepsis patients with the median age 65.9 years (Interquartile Range (IQR) [53.7-77.9]), which included 2,625 male (56.1%) and 3,367 white (71.9%) patients. The overall in-hospital mortality rate was 10.9%, and the median ICU length-of-stay was 2.8 days (IQR [1.6-5.6]). There were 1,893 patients (40.5%) treated with mechanical ventilation during the first three days. The mean baseline SOFA score obtained from the first 6 hours after ICU admission was 4.96 (Standard Deviation (SD): 2.8). Most of the patients (2,611, 55.8%) were in the medical intensive care unit (MICU). The overall demographic distributions of the validation cohorts from NMEDW(n=3,665) and eICU^21^ (n=12,282) are similar to the development cohort. Patients in validation cohort CEDAR (n=4,804) were older (median age 77 years (IQR [66.0-88.0]) compared to development cohort. The overall in-hospital mortality rates of patients in NMEDW, eICU, and CEDAR were 14.0%, 10.5%, and 199%, respectively. The median length-of-stay were 3.8 days (IQR [1.9-7.9]), 2.8 days (IQR [1.7-5.1]), 4.4 days (IQR [2.7-7.9]). There were 1,524 (41.6%), 5,772 (47.0%) and 2,263 (47.1%) patients that needed mechanical ventilation in the first three days. The mean baseline SOFA scores were 5.68 (SD:2.8), 5.9 (SD:3.1), and 6.4 (SD:3.1) in validation cohorts.

### Comparisons between Survivors and Nonsurvivors

In the development cohort, nonsurvivors were older than survivors, with a median age of 71.5 years (IQR, [59.9-80.9]) compared with 65.2 years for survivors (IQR, [53.2-77.4], p-value < 0.001). Nonsurvivors had higher comorbidity burden with a median Elixhauser index score ^26^7.0 (IQR [2.0-12.0]). Median ICU length-of-stay for nonsurvivors was 3.95 days (IQR [1.9-7.7]), and the rate of mechanical ventilation during the first three days was 59.8%. Nonsurvivors had higher baseline SOFA scores, with a mean value 7.1 (SD: 3.7). More nonsurvivors were admitted in MICU (Supplemental Table S1). Similar statistics in validation cohorts are shown in Supplemental Tables S2, S3, and S4.

### SOFA trajectory and the derived subphenotypes

Based on the pairwise patients’ SOFA trajectory similarity matrix obtained from DTW, we generated clustermaps with HAC (Supplemental Figure S2), where four distinct clusters were identified as subphenotypes. The number of clusters was determined according to multiple clustering indexes (Supplemental Appendix 6 and Table S5).

The overall trajectory and prevalence of each subphenotype across four cohorts are shown in Figure 2 and 3. Specifically, in the development cohort, the Rapidly Worsening subphenotype (n=612, 13.1%) was characterized by continuously increased SOFA scores from a mean (SD) of 4.5 (2.8) at admission to more than 7 at 72 hours. This subphenotype had the fewest patients. The Delayed Worsening subphenotype (n=960, 20.5%) was characterized by decreased SOFA scores within the first 48 hours from a mean (SD) of 5.2 (2.7) at baseline to 3.7 (2.8), followed by an increase over the last 24 hours. The Rapidly Improving subphenotype (n=1,932, 41.3%) was characterized by a consistent continuous improvement in SOFA scores from a mean (SD) of 5.54 (2.9) at baseline to less than 3. This was the most common subphenotype and it had the highest SOFA score at baseline. The Delayed Improving subphenotype (n=1,174, 25.1%) was characterized by an increase and then a gradual decrease in SOFA score over 72 hours. It had the lowest SOFA score at baseline with mean (SD) 4.0 (2.4). Similar trajectory trends were obtained in all three validation cohorts (Figure 2 and 3, Supplemental Appendix 3). Individual SOFA subscore trajectories for each subphenotype are provided in Supplemental Figures S3, S4, S10, and S14.

**Figure 2.**
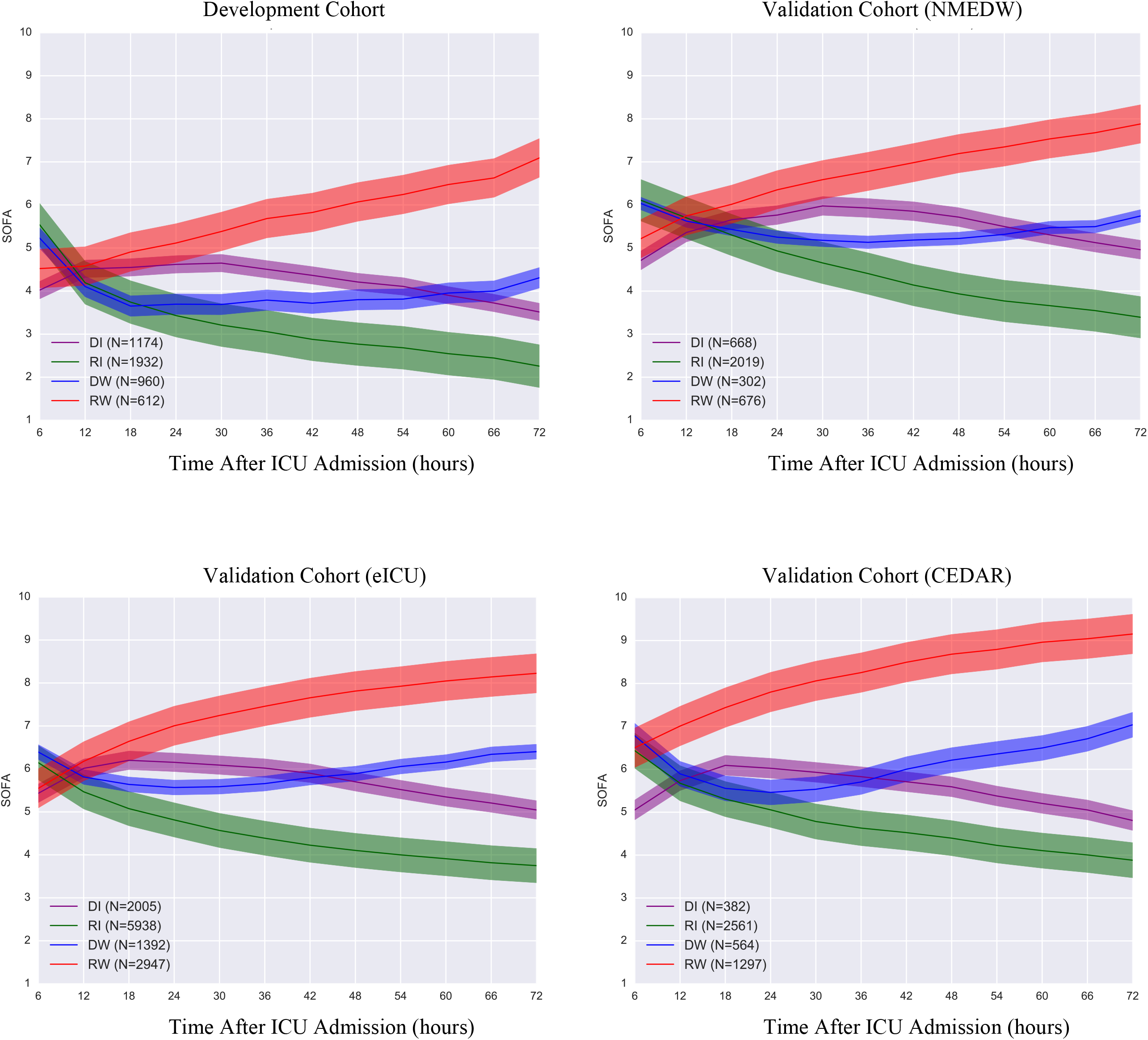
Sequential Organ Failure Assessment (SOFA) trajectories of the identified subphenotypes in development and three validation cohorts. DI: Delayed Improving; RI: Rapidly Improving; DW: Delayed Worsening; RW: Rapidly Worsening.

**Figure 3.**
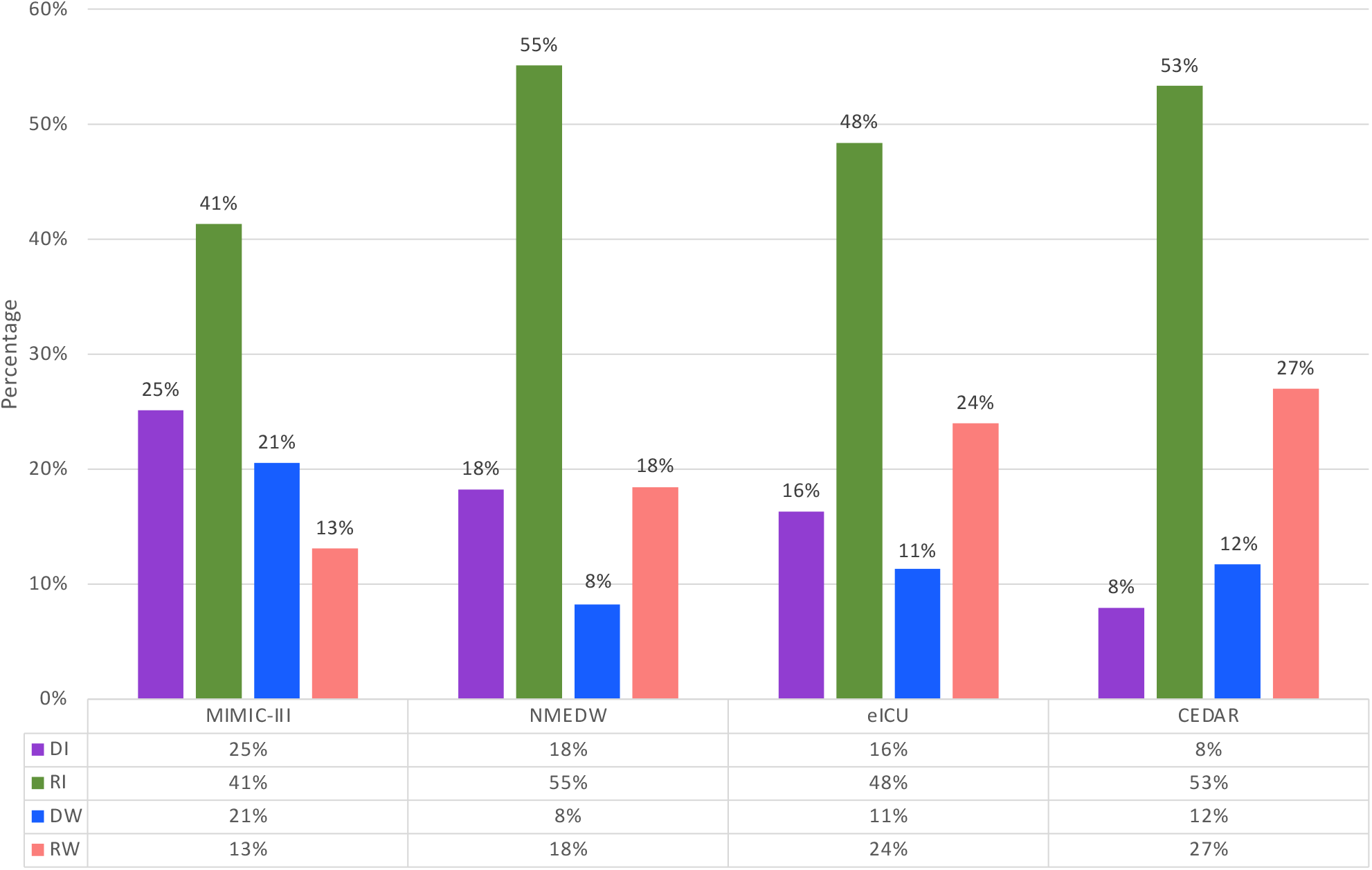
The prevalence of each subphenotype in development (MIMIC-III) and other three validation cohorts (NMEDW, eICU, CEDAR). DI: Delayed Improving; RI: Rapidly Improving; DW: Delayed Worsening; RW: Rapidly Worsening.

### Patient characteristics comparisons across subphenotypes

Patient characteristics differed across subphenotypes (Table 1, Figure 4, Figure 5, and Supplemental Table S6). Specifically, Rapidly Worsening patients had the highest rates of mechanical ventilation (46.41%), the highest median Elixhauser comorbidity burden value of 5 (IQR [0-10]) but the lowest baseline SOFA score compared to the other subphenotypes. They had the highest mortality rate (Figure 4(A) 28.3%, p-value<0.001) and a longer length of stay (Table 1, 2.9 days, p-value<0.001). Rapidly Improving patients had the lowest rate of mortality (Figure 4(A) 5.5%) and mechanical ventilation (37.9%), and the shortest length-of-stay (2.4 days). It had the highest proportion of patients meeting criteria for septic shock (15.5%, p-value=0.002). Delayed Improving and Delayed Worsening patients had lower rates of mortality (10.7%, 10.6%) and mechanical ventilation (42.5%, 39.3%) than the Rapidly Worsening subphenotype. The median age of the four subphenotypes were similar in the development cohort. Male patients were more common in all subphenotypes. Chord diagrams (Figure 5) showed the differences of subphenotypes in terms of abnormal clinical biomarkers. The Rapidly Worsening group was more likely to have patients with abnormal cardiovascular biomarkers (bicarbonate, troponin T or I, lactate) and hematologic (such as hemoglobin, INR, platelet, glucose, RDW). Patients in this subphenotype had a higher chronic comorbidity burden and had abnormal SOFA subscores including respiration, coagulation and liver. The Rapidly Improving patients were more likely to have abnormal inflammatory lab values (temperature, WBC, bands, CRP, albumin, lymphocyte percent) and abnormal cardiovascular, CNS and renal SOFA subscores. There was a lower chronic comorbidity burden in this subphenotype. Delayed Worsening group had more abnormal hematologic and respiration, coagulation, CNS, and SOFA renal subscores. Abnormal respiration, coagulation, cardiovascular SOFA subscores were strongly associated with Delayed Improving. The characteristics on validation cohorts are provided in Supplemental Appendix 4 and Tables S7, S8, S9, S10, S11, and S12. The associations between all comorbidities and subphenotypes were investigated and shown in Supplemental Tables S16, S17, S18, and S19. Multiple comorbidities such as congestive heart failure, renal failure, liver disease, cancer showed the differences among subphenotypes.

**Table 1.** Patient Characteristics among Subphenotypes in the Development Cohort.

| Characteristics | Total<br>(N=4,678) | DI<br>(N=1,174) | RI<br>(N=1,932) | DW<br>(N=960) | RW<br>(N=612) | P-value<sup>†</sup> |
| --- | --- | --- | --- | --- | --- | --- |
| Age, median (IQR) | 65.9 [53.7-77.9] | 67.25 [54.8-79.2] | 65.3 [53.3-77.2] | 66.9 [53.9-78.3] | 64.5 [52.5-76.7] | 0.204 |
| Sex, No. (%) |  |  |  |  |  |  |
| Male | 2625 (56.1) | 594 (50.6) | 1100 (56.9) | 548 (57.1) | 383 (62.6) | 0.081 |
| Female | 2053 (43.9) | 580 (49.4) | 832 (43.1) | 412 (42.9) | 229 (37.4) |  |
| Race, No. (%) |  |  |  |  |  | 0.207 |
| WHITE | 3367 (71.9) | 870 (74.1) | 1398 (72.4) | 670 (69.8) | 429 (70.1) |  |
| BLACK | 424 (9.1) | 92 (7.8) | 189 (9.8) | 101 (10.5) | 42 (6.9) |  |
| OTHER | 887 (18.9) | 212 (18.1) | 345 (17.9) | 189 (19.7) | 141 (23.0) |  |
| Elixhauser index, median (IQR) | 4.0 [0.0-9.0] | 4.0 [0.0-9.0] | 4.0 [0.0-9.0] | 4.0 [0.0-9.0] | 5.0 [0.0-10.0] | 0.015 |
| Length stay, median (IQR) | 2.8 [1.6-5.6] | 2.9 [1.8-6.2] | 2.4 [1.5-4.8] | 2.9 [1.7-5.3] | 2.9 [1.6-6.7] | <<br>0.001 |
| Mechanical ventilation at admission, No. (%) | 1893 (40.5) | 499 (42.5) | 733 (37.9) | 377 (39.3) | 284 (46.4) | <<br>0.001 |
| Baseline SOFA, mean (SD) | 4.96 (2.8) | 4.0 (2.4) | 5.5 (2.9) | 5.2 (2.7) | 4.5 (2.8) | <<br>0.001 |
| ICU unit at admission, No. (%) |  |  |  |  |  | 0.037 |
| SICU | 771 (16.5) | 185 (15.8) | 341 (17.7) | 135 (14.1) | 110 (17.9) |  |
| CCU | 443 (9.5) | 117 (9.9) | 167 (8.6) | 94 (9.8) | 65 (10.6) |  |
| TSICU | 593 (12.7) | 173 (14.7) | 226 (11.7) | 119 (12.4) | 75 (12.3) |  |
| MICU | 2611 (55.8) | 634 (54.0) | 1087 (56.3) | 569 (59.3) | 321 (52.5) |  |
| CSRU | 260 (5.6) | 65 (5.5) | 111 (5.8) | 43 (4.5) | 41 (6.7) |  |
| Admission location, No. (%) |  |  |  |  |  | 0.196 |
| Transfer from other hospital | 810 (17.3) | 213 (18.1) | 304 (15.7) | 165 (17.2) | 128 (20.9) |  |
| Emergency room | 1497 (32.0) | 355 (30.2) | 628 (32.5) | 328 (34.2) | 186 (30.4) |  |
| Clinic referral | 1985 (42.4) | 493 (41.9) | 847 (43.8) | 396 (41.3) | 249 (40.7) |  |
| Transfer from ward | 4 (0.1) | 2 (0.2) | 1 (0.1) | 1 (0.1) | 0 (0.0) |  |
| Physician referral | 367 (7.9) | 106 (9.0) | 145 (7.5) | 69 (7.2) | 47 (7.7) |  |
| Transfer from skilled nursing facility | 15 (0.3) | 5 (0.4) | 7 (0.4) | 1 (0.1) | 2 (0.3) |  |
| Infection item, No. (%) |  |  |  |  |  |  |
| Central nervous system | 56 (1.2) | 10 (0.9) | 27 (1.4) | 8 (0.8) | 11 (1.8) | 0.189 |
| Intra-abdominal | 880 (18.8) | 230 (19.6) | 363 (18.8) | 172 (17.9) | 115 (18.8) | 0.808 |
| Pneumonia | 1257 (26.9) | 328 (27.9) | 494 (25.6) | 262 (27.3) | 173 (28.3) | 0.385 |
| Septicemia bacteremia | 1587 (33.9) | 359 (30.6) | 717 (37.1) | 300 (31.3) | 211 (34.5) | <<br>0.001 |
| Skin soft tissue | 276 (5.9) | 60 (5.1) | 140 (7.3) | 42 (4.4) | 34 (5.6) | 0.008 |
| Urinary tract | 1044 (22.3) | 276 (23.5) | 439 (22.7) | 228 (23.8) | 101 (16.5) | 0.003 |
| **Septic shock, No. (%)** | **635 (13.6)** | **148 (12.6)** | **299 (15.5)** | **101 (10.5)** | **87 (14.2)** | **0.002** |

**Figure 4.**
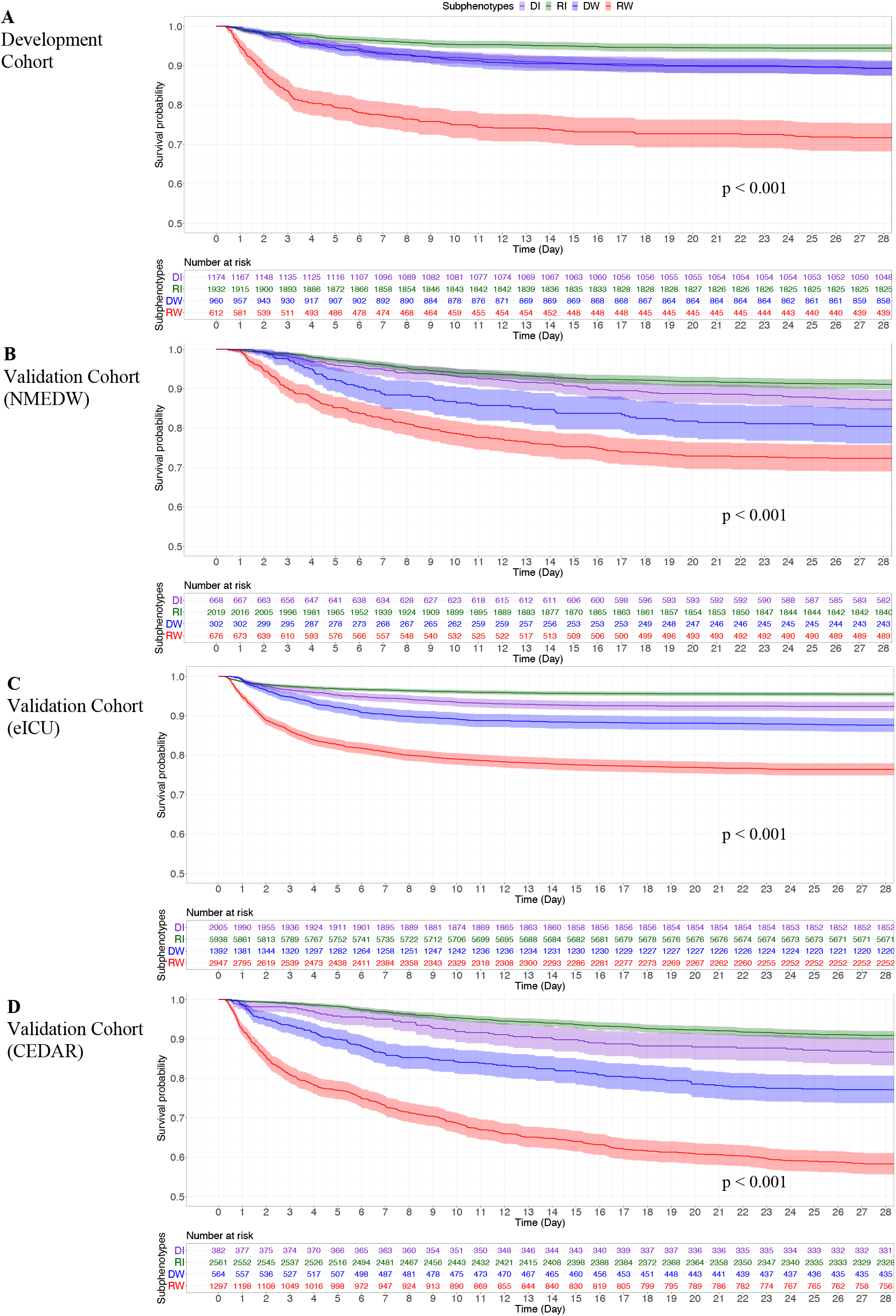
Survival analysis in terms of the identified subphenotypes in development and three validation cohorts. DI: Delayed Improving; RI: Rapidly Improving; DW: Delayed Worsening; RW: Rapidly Worsening. The (A), (B), (C), and (D) show the survival analysis results in development and three validation cohorts, respectively.

**Figure 5.**
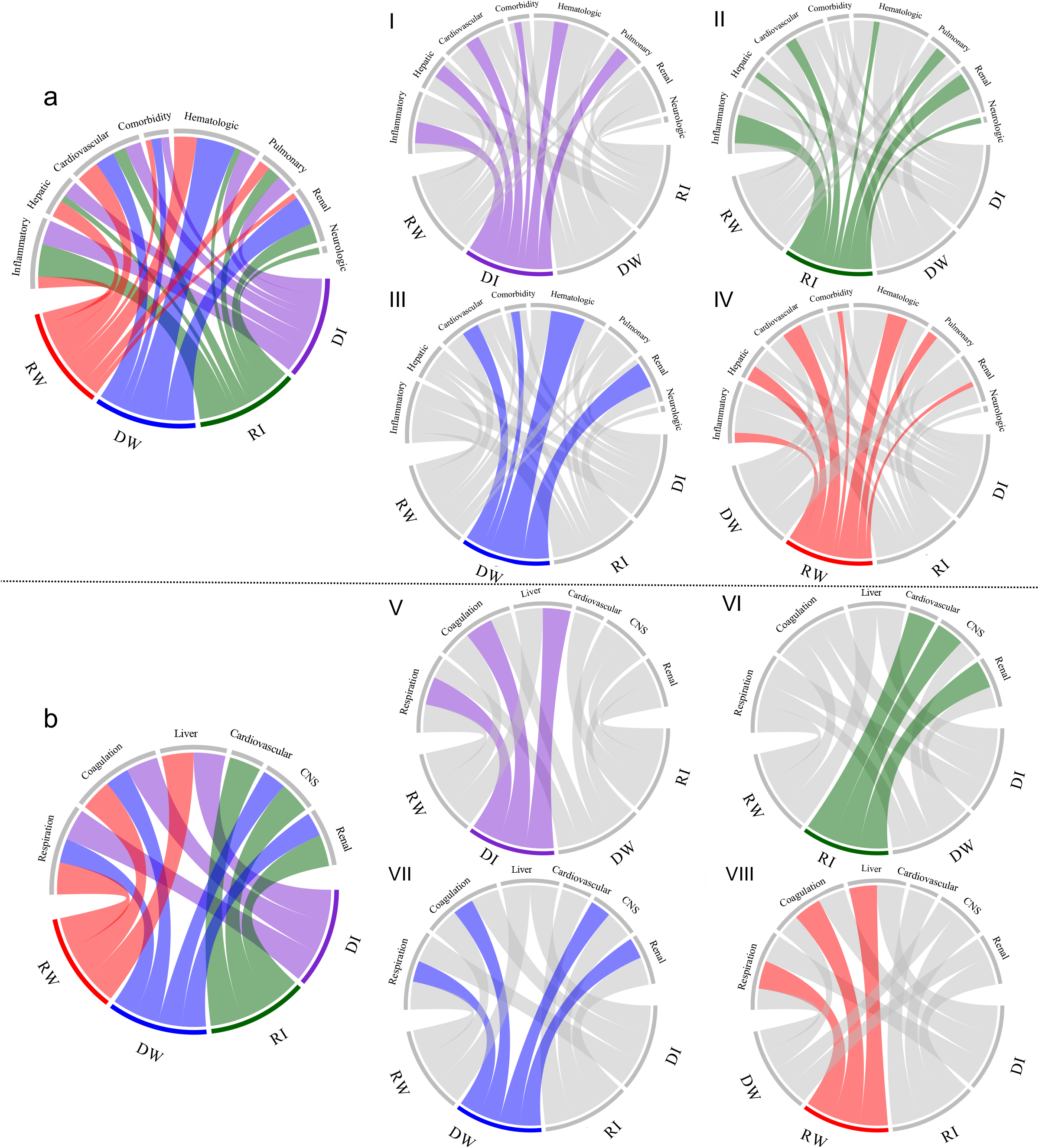
Chord diagrams showing abnormal variables by subphenotype in development cohort. a: abnormal biomarkers vs. all subphenotypes; I: abnormal biomarkers vs. DI; II: abnormal biomarkers vs. RI; III: abnormal biomarkers vs. DW; IV: abnormal biomarkers vs. RW; b: abnormal subscores vs. all subphenotypes; V: abnormal subscores vs. DI; VI: abnormal subscores vs. RI; VII: abnormal subscores vs. DW; VIII: abnormal subscores vs. RW. DI: Delayed Improving; RI: Rapidly Improving; DW: Delayed Worsening; RW: Rapidly Worsening.

### Subphenotype reproducibility and prediction

Sensitivity analysis with another clustering approach GBTM confirmed the four subphenotypes with the data from development cohort (Supplemental Figure S8). Patients’ memberships of the four subphenotypes re-derived by GBTM were highly consistent with those obtained from HAC (Supplemental Figure S9), and thus we did not find substantial changes in clinical characteristics of those subphenotypes derived from the sensitivity analysis (Supplemental Table S13).

We trained random forest models for predicting subphenotypes according to early-stage patient characteristics. Overall, with the first 6 hours after ICU admission, the models obtained the accuracy of 0.78 (95% Confidence Interval [CI] [0.77, 0.8]) in development cohort and 0.79 (95% CI [0.78, 0.8]), 0.81 (95% CI [0.8, 0.84]), and 0.82 (95% CI [0.81, 0.84]) in NMEDW, eICU, and CEDAR validation cohorts respectively. Predictor contributions on four cohorts are shown in Figure 6 and Supplemental Figures S5, S11, and S15, which demonstrated different patterns when predicting different subphenotypes. For example, creatinine, bicarbonate, RDW, and BUN contributes more for predicting the Rapidly Improving group, while platelet, INR, AST and lactate contributed more to the prediction of the Rapidly Worsening group. The prediction performance at successive time points are shown in Supplemental Figure S18. The accuracy increased to 0.87 (95% CI [0.86, 0.88]) in development cohort and 0.86 (95% CI [0.85, 0.88]), 0.86 (95% CI [0.85, 0.87]), and 0.84 (95% CI [0.83, 0.85]) in NMEDW, eICU, and CEDAR validation cohorts at the 24 hours after ICU admission, respectively.

**Figure 6.**
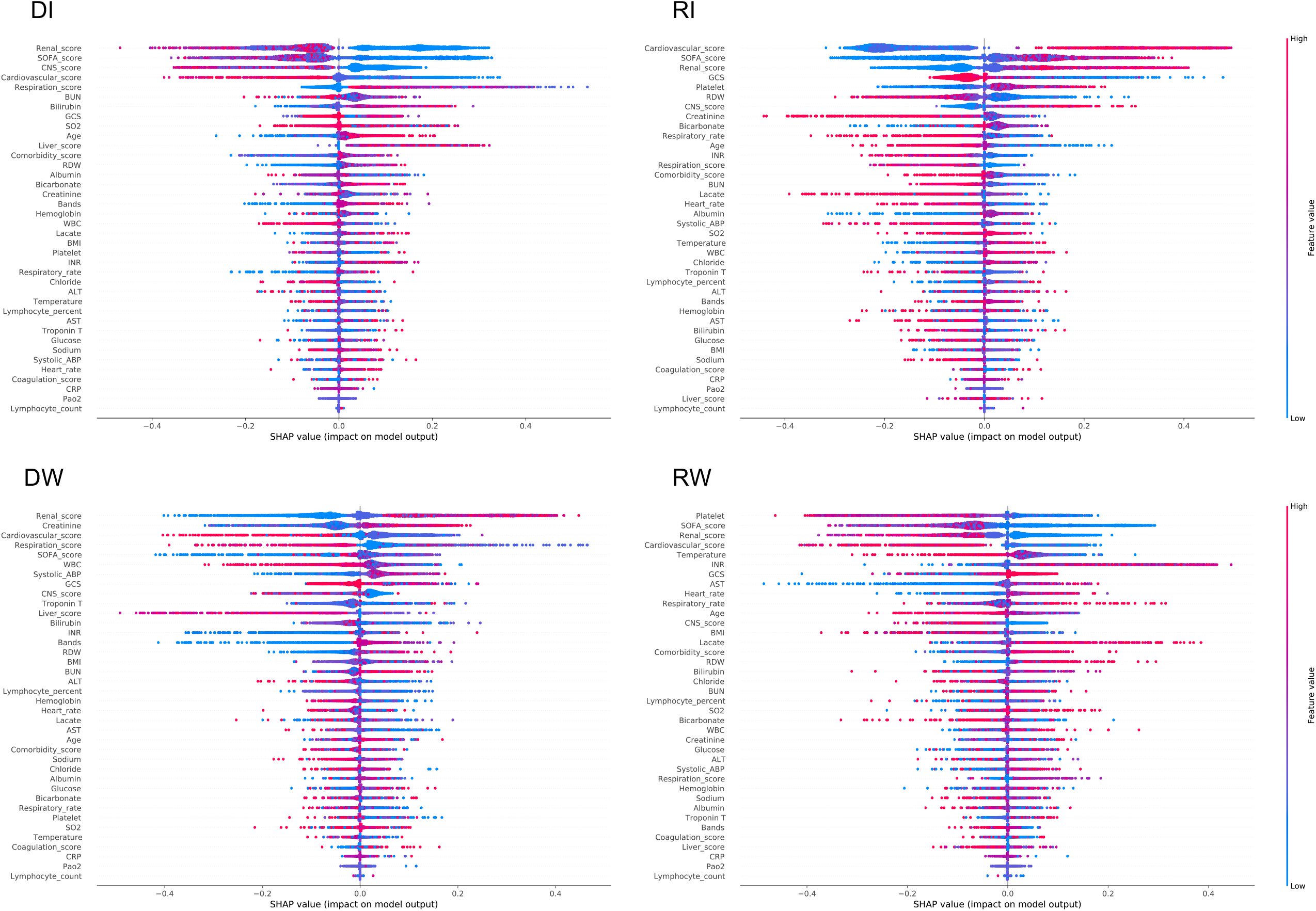
SHAP value-based predictor contribution to the subphenotype prediction of the predictive model in development cohort. Features’ importance is ranked based on SHAP values. In this figure, each point represented a single observation. The horizontal location showed whether the effect of that value was associated with a positive (a SHAP value greater than 0) or negative (a SHAP value less than 0) impact on prediction. Color showed whether the original value of that variable was high (in red) or low (in blue) for that observation. For example, in RW, a low Platelets value had a positive impact on the RW subphenotype prediction; the “low” came from the blue color, and the “positive” impact was shown on the horizontal axis. DI: Delayed Improving; RI: Rapidly Improving; DW: Delayed Worsening; RW: Rapidly Worsening.

## Discussion

We reported four sepsis subphenotypes based on dynamic organ dysfunction trajectories using a data-driven methodology. DTW was used to calculate patients’ SOFA trajectory similarities because of its capability of capturing heterogeneous evolution among the temporal sequences robustly, based on which HAC was leveraged to identify patient groups with similar trajectories. The subphenotypes identified were Rapidly Worsening, Delayed Worsening, Rapidly Improving, and Delayed Improving. Patients in the Rapidly Worsening subphenotype had progressive organ dysfunction with the ongoing ICU stay. The two Delayed groups had unstable organ dysfunction over the study period and the Rapidly Improving group had the highest admission organ dysfunction but quickly improved. Outcomes followed SOFA trajectory across each subphenotype were irrespective of traditional baseline SOFA score and septic shock categories.

A major strength of this analysis is that we have identified time-dependent progression patterns that may be related to the differential response of specific organ dysfunction to standard of care interventions. For example, the Rapidly Improving group had cardiovascular and respiratory failure at admission that resolved over 72 hours. The Rapidly Worsening groups developed multisystem organ failure including visceral organ dysfunction, specifically liver failure in addition to cardiovascular and respiratory failure. These differential patterns suggest varying time-dependent, treatment responsive organ dysfunction pathophysiology in sepsis. The cardiovascular and respiratory subscores are driven by the vasopressor dose and PaO2/FiO2 respectively, which may respond to therapeutic interventions such as corticosteroids, volume resuscitation, and the application of PEEP or therapeutic suctioning.^27^ However, as demonstrated by our analysis, sepsis-related renal and liver failure may be less modifiable with our current therapeutic strategies over the past twenty years.^28,29^ Our study highlights that patterns of organ dysfunction in patients with sepsis are Rapidly Improving, Rapidly Worsening and Delayed. Each of these patterns may be due to a different pathophysiology and benefit from different treatments in the future. However, these findings have immediate implications for those designing clinical trial endpoints such as change in SOFA subscore.^30^ Moreover, enrolling patients with a Rapidly Improving phenotype into a trial evaluating a therapeutic agent to reduce the duration of organ dysfunction will unlikely reveal a difference.

It deserves noting that our Rapidly Improving patients had better outcomes across all patients studied, but still represented 21%, 36%, 21%, and 24% of all deaths in our development and validation cohorts (NMEDW, eICU, and CEDAR cohorts) respectively, despite an overall 5%, 10%, 5%, and 9% in-hospital mortality. This low mortality rate but high numbers of absolute deaths highlights that further research is needed to understand the cause of death in patients with rapidly improving organ dysfunction in sepsis.^31^ The Rapidly Worsening subphenotype was less common compared to rapidly improving and may represent patients with our classical understanding of septic shock.^32^ More recent evidence suggests that the pathophysiology of early, progressive organ dysfunction in our Rapidly Worsening patients may be due to over exuberant activation of necroinflammatory cell death pathways in multiple organs, highlighting the need for novel treatment strategies.^33-35^ The Delayed Worsening and Improving subphenotypes, had intermediate outcomes across our cohorts, and more nuanced differences in clinical characteristics. These trajectories may be influenced by non-resolving inflammation or immune paralysis.^36,37^ Further understanding of the biology underlying these subphenotypes will be critical to develop the next generation of treatments for sepsis in all its forms.

The potential for distinct pathophysiologic etiologies for the differential trajectories is supported by the differential patterns of organ dysfunction, infectious source, vital signs, inflammatory, hematologic, and cardiovascular variables at admission to the ICU. As shown in Figure 5, and Supplemental Figures S6, S7, S12, S13, S16, and S17, there were different variables associated with different groups over the course of the study. For example, those patients of Rapidly Improving were more likely to have more abnormal inflammatory markers (such as WBC, bands, albumin, temperature, lymphocyte) and more abnormal values on cardiovascular, and CNS subscores. They were also more likely to have urosepsis. There was a lower comorbidity score in patients with this subphenotype, which suggests that sepsis outcomes may be more dependent on underlying illness. The Rapidly Worsening patients had more comorbidities and distinct derangements in clinical variables associated with metabolic acidosis and hypoperfusion, e.g. a low bicarbonate and higher lactate, and disseminated intravascular coagulation, e.g. low platelets and a higher INR and respiratory failure. Both of the Delayed subphenotypes had less specific variables associated with group membership, including inflammatory, hepatic, hematologic and pulmonary associated with Delayed Improvement and hematologic, cardiovascular and renal variables associated with Delayed Worsening. These differences may be related to secular trends in therapeutics and differing case mixes in each cohort.

We built multivariable prediction models for the identified trajectory subphenotypes from patient baseline characteristics and early-stage clinical features. Several interesting findings were obtained. (1) A high comorbidity score tended to predict the subphenotypes of Rapidly Worsening because patients with high comorbidity burden were more likely to present worse organ dysfunction in ICU; (2) The roles of lab tests and vital signs were different on prediction. For example, low Platelets had a positive impact on the Rapidly Worsening prediction and high Platelets had a positive impact on the Rapidly Improving prediction. These prediction models may enhance the clinical utility of the identified subphenotypes in practice, as they can be predicted with the EHR information captured within the early hours of ICU admission, especially for Rapid Improving and Rapid Worsening subphenotypes, which has important clinical implications as discussed above. Our model can be implemented within the EHR system as a risk calculator for subphenotype assignments.

Our manuscript complements and adds to other recent study of sepsis subphenotypes. For example, Seymour et al. and Knox et al. each identified four subphenotypes that were associated with organ dysfunction patterns and clinical outcomes in patients with sepsis using a panel of baseline clinical variables.^5,10^ There is some overlap in our high risk groups, notably both include liver injury and shock. However, our work demonstrates that the difference in outcome in this group is due to progressive non-resolving organ dysfunction that calls for novel treatments. Prior work by Ferreira et al and Sakr et al used changes in the SOFA score after ICU admission to improve prognostic stratification in sepsis, but did not use these changes to establish subphenotypes. Bhavani et al. used longitudinal temperature trajectories to identify four sepsis subphenotypes, with significant variability in inflammatory markers and outcomes, highlighting the potential for novel immune signatures to be uncovered through trajectory analysis.^1^ Differential organ dysfunction trajectory may be related to the immune response but may also be explained by differences in preexisting frailty, effective source control, resuscitation and processes of care.

This study has several limitations. First, our sepsis subphenotypes were identified based on the data-driven method, which may not be directly related to underlying differences in biology. Integration of biological data may help refine our understanding of differential disease progression and the potential for therapeutics to alter the course. Second, although we used many separate hospitals in validation, all of them are located in the United States, which may limit generalizability to other locations of care. Moreover, these observational cohorts may not directly reflect sepsis clinical trial populations but are representative of academic and community hospitals across the United States. Third, we did not evaluate the effect of specific randomized interventions on SOFA score trajectory. Fourth, this identified sepsis subphenotypes only focused on patients admitted to an ICU, which is subject to differences in ICU admission practices across institutions. Last but not the least, we did not investigate the association between care processes and the subphenotypes, which would be an important topic in future research.

## Conclusion

We discovered four sepsis subphenotypes with different natural histories following admission to the ICU. Our results suggest that these subphenotypes represent a differential host pathogen response in the setting of current standard of care therapy. Understanding differential trajectory has implications for the design and predictive enrichment of therapeutic clinical trials.^38^ Further understanding of the underlying biology of subphenotypes may reveal insights into sepsis pathophysiology and improve the personalization of sepsis management.

## Data Availability

The data and code are available upon request.

## Abbreviations

ICU: Intensive Care Unit
DTW: Dynamic Time Warping
HAC: Hierarchical Agglomerative Clustering
SOFA: Sequential Organ Failure Assessment
GBTM: Group-Based Trajectory Modeling
LCA: Latent Class Analysis
EHR: Electronic Health Record
MIMIC-III: Medical Information Mart for Intensive Care III database
BIDMC: Beth Israel Deaconess Medical Center
NMEDW: Northwestern Medicine Enterprise Data Warehouse
CEDAR: Critical carE Database for Advanced Research
NYP/WCMC: NewYork-Presbyterian/Weill Cornell Medical Center
CNS: Central Nervous System
LOCF: Last Observation Carried Forward
NOCB: Next Observation Carried Backward
SHAP: Shapley Additive Explanations
MICU: Medical Intensive Care Unit
SD: Standard Deviation
WBC: White Blood Cell Count
RDW: Red Blood Cell Distribution Width
CRP: C-reactive protein
CI: Confidence Interval
AST: Aspartate Aminotransferase.

## Acknowledgements

The work of ZX, CZ, HZ and FW are supported by NSF 1750326, NIH RF1AG072449 and NIH R01MH124740. The work of ES is supported by NHLBI K23HL151876. The work of CM and YL are supported in part by NIH 1R01LM013337 and U01TR003528.

## Author contributors

ES and FW for conceptualization, investigation, writing, reviewing and editing of the manuscript. ZX for data analysis, drafting, editing and reviewing manuscript. CM for data analysis, editing and reviewing manuscript. CS, HZ, IS, LT and DP for discussion, commenting and editing the manuscript. YL, CM, ZX, FW, and ES verified the data. YL and CM had access to the raw data from the NMEDW. FW, ES, and ZX had access to the raw data from the CEDAR, eICU, and MIMIC-III.

## Availability of data and materials

The deidentified data from development cohort (MIMIC-III) and data from validation cohort (eICU) can be obtained after approval of proposal with a signed data access agreement by checking physionet (The Research Resource for Complex Physiologic Signals, https://physionet.org/). For validation cohorts (NMEDW and CEDAR), data access are not covered by our data transfer agreements. All source codes in this study are available at https://github.com/xuzhenxing2019/sepsis_subphenotype. Our implementation is based on Python 3.7 and R 3.6. More specifically, clustering models were implemented by using Python packages ‘scikit-learn 0.23.2’ (https://scikit-learn.org/stable/) and ‘scipy 1.5.3’ (https://www.scipy.org). The implementation of SHAP is based on ‘SHAP 0.35.0’ (https://shap.readthedocs.io/en/latest/). R package ‘NbClust’ (https://cran.r-project.org/web/packages/NbClust/NbClust.pdf) was used to determine the optimal number of clusters in agglomerative hierarchical clustering. Chord diagrams were created using R package ‘circlize’ (https://cran.r-project.org/web/packages/circlize/index.html). Statistical tests and survival analyses were performed based on R.

## Declarations

### Ethics approval and consent to participate

Consent obtained for use of MIMIC-III, eICU, NMEDW and CEDAR databases.

### Consent for publication

Not applicable

