## supplementary for "Sepsis Subphenotyping Based on Organ Dysfunction Trajectory"

#### Supplementary Materials

**Tables**

**Table S1.** Patient characteristics comparisons between survivors and nonsurvivors in the development cohort

**Table S2.** Patient characteristics comparisons between survivors and nonsurvivors in the NMEDW validation cohort

**Table S3.** Patient characteristics comparisons between survivors and nonsurvivors in the eICU validation cohort

**Table S4.** Patient characteristics comparisons between survivors and nonsurvivors in the CEDAR validation cohort

**Table S5.** Multiple clustering metrics statistics and cluster number determination

**Table S6.** Patient physiological characteristics comparisons among subphenotypes in the development cohort

**Table S7.** Patient basic characteristics comparisons among subphenotypes in the NMEDW validation cohort

**Table S8.** Patient physiological characteristics comparisons among subphenotypes in the NMEDW validation cohort

**Table S9.** Patient basic characteristics comparisons among subphenotypes in the eICU validation cohort

**Table S10.** Patient physiological characteristics comparisons among subphenotypes in the eICU validation cohort

**Table S11.** Patient basic characteristics comparisons among subphenotypes in the CEDAR validation cohort

**Table S12.** Patient physiological characteristics comparisons among subphenotypes in the CEDAR validation cohort

**Table S13.** Patient physiological characteristics comparisons among subphenotypes in terms of sensitivity analysis (GBTM) in the development cohort

**Table S14.** Missing data (no., %) in cohorts for subphenotyping and predicting variables.

**Table S15.** International classification of diseases, ninth revision, clinical modification codes used to identify infectious syndromes.

**Table S16.** The associations between comorbidities and subphenotypes in the MIMIC-III development cohort

**Table S17.** The associations between comorbidities and subphenotypes in the NMEDW validation cohort

**Table S18.** The associations between comorbidities and subphenotypes in the eICU validation cohort

**Table S19.** The associations between comorbidities and subphenotypes in the CEDAR validation cohort

**Figures**

**Figure S1.** Patient exclusion criteria for building development and validation cohorts

**Figure S2.** Clustergrams of hierarchical clustering. Horizontal and vertical axes represent patients. Color intensity denotes normalized pairwise patient similarity derived using Dynamic Time Warping (DTW). All clustergrams suggest optimal cluster number 4.

**Figure S3.** The trajectories of the subphenotypes in terms of subscores in the development cohort. DI: Delayed Improving; RI: Rapidly Improving; DW: Delayed Worsening; RW: Rapidly Worsening.

**Figure S4.** The trajectories of the subphenotypes in terms of subscores in the NMEDW validation cohort. DI: Delayed Improving; RI: Rapidly Improving; DW: Delayed Worsening; RW: Rapidly Worsening.

**Figure S5.** Predictor contribution to the prediction of the predictive model in the NMEDW validation cohort. DI: Delayed Improving; RI: Rapidly Improving; DW: Delayed Worsening; RW: Rapidly Worsening.

**Figure S6.** Chord diagrams showing abnormal clinical variables by subphenotype in the NMEDW validation cohort. a: abnormal biomarkers vs. all subphenotypes; I: abnormal biomarkers vs. DI; II: abnormal biomarkers vs. RI; III: abnormal biomarkers vs. DW; IV: abnormal biomarkers vs. RW. DI: Delayed Improving; RI: Rapidly Improving; DW: Delayed Worsening; RW: Rapidly Worsening.

**Figure S7.** Chord diagrams showing abnormal subscores by subphenotype in the NMEDW validation cohort. a: abnormal subscores vs. all subphenotypes; I: abnormal subscores vs. DI; II: abnormal subscores vs. RI; III: abnormal subscores vs. DW; IV: abnormal subscores vs. RW. DI: Delayed Improving; RI: Rapidly Improving; DW: Delayed Worsening; RW: Rapidly Worsening.

**Figure S8** Sequential Organ Failure Assessment (SOFA) trajectories of the subphenotypes and survival analysis in terms of the identified subphenotypes by GBTM. The (A) describes the SOFA trajectories of the subphenotypes re-derived by GBTM in development cohort. The (B) shows the survival analysis results in subphenotypes re-derived by GBTM in the development cohort. DI: Delayed Improving; RI: Rapidly Improving; DW: Delayed Worsening; RW: Rapidly Worsening.

**Figure S9.** Confusion matrices for comparing the subphenotypes obtained by DTW and HAC, and GBTM in development cohort. DI: Delayed Improving; RI: Rapidly Improving; DW: Delayed Worsening; RW: Rapidly Worsening. DTW: Dynamic Time Warping; HAC: Hierarchical Agglomerative Clustering. GBTM: Group-Based Trajectory Modeling.

**Figure S10.** The trajectories of the subphenotypes in terms of subscores in the eICU validation cohort. DI: Delayed Improving; RI: Rapidly Improving; DW: Delayed Worsening; RW: Rapidly Worsening.

**Figure S11.** Predictor contribution to the prediction of the predictive model in the eICU validation cohort. DI: Delayed Improving; RI: Rapidly Improving; DW: Delayed Worsening; RW: Rapidly Worsening.

**Figure S12.** Chord diagrams showing abnormal clinical variables by subphenotype in the eICU validation cohort. a: abnormal biomarkers vs. all subphenotypes; I: abnormal biomarkers vs. DI; II: abnormal biomarkers vs. RI; III: abnormal biomarkers vs. DW; IV: abnormal biomarkers vs. RW. DI: Delayed Improving; RI: Rapidly Improving; DW: Delayed Worsening; RW: Rapidly Worsening.

**Figure S13.** Chord diagrams showing abnormal subscores by subphenotype in the eICU validation cohort. a: abnormal subscores vs. all subphenotypes; I: abnormal subscores vs. DI; II: abnormal subscores vs. RI; III: abnormal subscores vs. DW; IV: abnormal subscores vs. RW. DI: Delayed Improving; RI: Rapidly Improving; DW: Delayed Worsening; RW: Rapidly Worsening.

**Figure S14.** The trajectories of the subphenotypes in terms of subscores in the CEDAR validation cohort. DI: Delayed Improving; RI: Rapidly Improving; DW: Delayed Worsening; RW: Rapidly Worsening.

**Figure S15.** Predictor contribution to the prediction of the predictive model in the CEDAR validation cohort. DI: Delayed Improving; RI: Rapidly Improving; DW: Delayed Worsening; RW: Rapidly Worsening.

**Figure S16.** Chord diagrams showing abnormal clinical variables by subphenotype in the CEDAR validation cohort. a: abnormal biomarkers vs. all subphenotypes; I: abnormal biomarkers vs. DI; II: abnormal biomarkers vs. RI; III: abnormal biomarkers vs. DW; IV: abnormal biomarkers vs. RW. DI: Delayed Improving; RI: Rapidly Improving; DW: Delayed Worsening; RW: Rapidly Worsening.

**Figure S17.** Chord diagrams showing abnormal subscores by subphenotype in the CEDAR validation cohort. a: abnormal subscores vs. all subphenotypes; I: abnormal subscores vs. DI; II: abnormal subscores vs. RI; III: abnormal subscores vs. DW; IV: abnormal subscores vs. RW. DI: Delayed Improving; RI: Rapidly Improving; DW: Delayed Worsening; RW: Rapidly Worsening.

**Figure S18.** The accuracy of predicting four subphenotypes at successive time points (hours 6, 24, 36, 48, 60) after ICU admission in development and validation cohorts.

**Figure S19.** The illustration of using DTW during obtaining matched sequences. (c) shows the warping path induced by the DTW matching, where the blue cells indicate the best matching pairs for the points in the two sequences.

**Figure S20.** An example: the illustration of patient's SOFA trajectory.

**Appendix 1 The detailed description in terms of cohorts**

Study data used in our study were from the Medical Information Mart for Intensive Care III (MIMIC-III) database. The collection of MIMIC-III dataset was passive and de-identified, which was in compliance with the Health Insurance Portability and Accountability Act (HIPAA) Privacy Rule and did not produce significant impacts on patient safety. This database consisted of patients admitted to the Beth Israel Deaconess Medical Center (BIDMC, Boston, MA). This database included information such as patient demographics, vital signs, laboratory test results. A total of 23,620 ICU admissions in the Meta Vision system of MIMIC-III were analyzed. We excluded some patients such as nonadults, secondary (or greater) admissions for patients to avoid repeated measures, admissions with missing data. The final cohort contained 4,678 patients for further analysis. In addition, in the first validation cohort that is built based on patient's records from Northwestern Medicine Enterprise Data Warehouse (NMEDW) from 2012 to 2019, a total of 67,850 ICU admissions were analyzed. We finally obtained 3,665 patients for further analysis. NMEDW contains 11 hospitals, including Northwestern Memorial Hospital, Northwestern Medicine Central DuPage Hospital, etc., more details can be found in [1]. For the second validation cohort, eICU Collaborative Research Database (eICU-CRD) [9], which is a deidentified and publicly available data set that meets the safe harbor provision of the US Health Insurance Portability and Accountability Act (HIPAA). The eICU-CRD is a multi-center database sourced from the Philips eICU program, a telemedicine initiative where healthcare workers remotely monitor acutely ill patients. It comprises 200,859 patient unit encounters for 139,367 unique patients admitted between 2014 and 2015 across the United States. The patient information includes demographics, vital sign measurements, care plan documentation, the severity of illness measures, diagnosis information, treatment information, and more. For the third validation cohort, the Critical carE Database for Advanced Research (CEDAR) [10], which was built on NewYork-Presbyterian/Weill Cornell Medical Center (NYP/WCMC) from admissions dating from 2001-2020. The NYP/WCMC was an academic medical center in New York City located on the Upper East Side of Manhattan, including 862 beds in total and 118 beds for ICU units across dedicated cardiac, cardiothoracic surgery, medical, surgical, pediatric, and neurosurgical units. Figure S1 descripted the inclusion and exclusion criteria for the selection process. In particular, for each database, we excluded non-adult (Age<18). We excluded secondary (or greater) admissions for patients to avoid repeated measures. We excluded admissions to the cardiothoracic surgical service and admissions with missing data. We excluded patients if there were not met Sepsis-3 criteria in the first 6 hours since ICU admission. Specifically, we included patients if they had evidence of a suspected infection defined as the combination of administration of antibiotics (oral or parenteral) and a body fluid culture specimen obtained (blood, urine, body fluid or cerebrospinal fluid), the first of which was required within the first 6 hours of ICU presentation. For SOFA score rule, we used an absolute SOFA score of >=2. Two sepsis studies based on Sepsis-3 definition [15][16] were referred to during selecting patients in each database. The SOFA score was computed from six organ-specific subscores including respiration, coagulation, liver, cardiovascular, CNS, renal. They were computed based on parameters including PaO2, FiO2, platelets, bilirubin, MAP, dopamine, dobutamine, epinephrine, norepinephrine, GCS, creatinine, urine output. The detailed scoring rule can be referred to Singer et.al study [18].

**Appendix 2 The detailed description about statistical analysis variables and test techniques**

(1) Demographics: age, gender, and race.
(2) Biomarkers: C-reactive protein (CRP), bands, temperature, white blood cell count (WBC), SO2, PaO2, respiratory rate, bicarbonate, heart rate, lactate, systolic blood pressure, troponin T or I, blood urea nitrogen (BUN), creatinine, alanine aminotransferase (ALT), aspartate aminotransferase (AST), bilirubin, Glasgow coma scale score (GCS), hemoglobin, INR, platelets; albumin, chloride, glucose, sodium, red blood cell distribution width (RDW).
(3) The type of ICU unit: TSICU (Thoracic Surgery ICU), MICU (Medical ICU), SICU (Surgical ICU), CCU (Coronary Care Unit), CSRU (Cardiac Surgery ICU).
(4) Infection sources: Central nervous system, Intra-Abdominal, Pneumonia, Septicemia bacteremia, Skin soft tissue, Urinary tract. These infection items are defined based on ICD-9 code (see Table S15). More details can be referred to [17].
(5) Admission location: Transfer from other hospital, Emergency room, Clinic referral, Transfer from ward, Physician referral, Transfer from skilled nursing facility.
(6) Multiple statistical test techniques were performed to discover association analyses in terms of those variables. Specifically, one-way analysis of variance (ANOVA, with Tukey HSD post hoc test), Kruskal–Wallis test (with Dunn post hoc test), student's t-test, Mann-Whitney test, Chi-square test, and Fisher's exact test were considered to use when appropriate. In addition, multiple testing utilizing false discovery rate (FDR) estimation was considered to correct the p-values. The techniques of analysis of covariance

(ANCOVA) were used for the between-subphenotype comparisons, which were based on general linear model.

**Appendix 3 The detailed analysis in terms of subphenotypes in the validation cohorts**
In the NMEDW validation cohort, four subphenotypes ("Delayed Improving", n=668, 18.2%), ("Rapidly Improving", n=2,019, 55.1%), ("Delayed Worsening", n=302, 8.2%), ("Rapidly Worsening" (n=676, 18.4%). In the eICU validation cohort, four subphenotypes ("Delayed Improving", n=2005, 16.3%), ("Rapidly Improving", n=5938, 48.4%), ("Delayed Worsening", n=1392, 11.3%), ("Rapidly Worsening" (n=2947, 23.9%). In the CEDAR cohort, four subphenotypes ("Delayed Improving", n=382, 7.9%), ("Rapidly Improving", n=2561, 53.3%), ("Delayed Worsening", n=564, 11.7%), ("Rapidly Worsening" (n=1297, 26.9%). We obtained the similar results with development cohort. Some specific similar and different characteristics in terms of trends derived on development and validation cohorts are summarized as follows: (1) four similar trend were obtained, including increasing continuously, decreasing firstly and keeping stable and slight increasing finally, decreasing sharply, increasing firstly and then decreasing gradually; (2) most of the patients kept decreasing and stable (e.g., "Delayed Improving", "Rapidly Improving", and "Delayed Worsening"). (3) The range of SOFA scores at baseline among these subphenotypes were different. For the development cohort, the range of average SOFA score at the baseline among the four subphenotypes was from 4.0 to 5.5. For the NMEDW validation cohort, the range was from 4.7 to 6.1. For the eICU validation cohort, the range was from 5.4 to 6.4. The fluctuation in these cohorts was smaller than the SOFA score in the development cohort. For the CEDAR validation cohort, the range was from 5.1 to 6.9. The fluctuation in this cohort was larger than the SOFA score in the development cohort and other validation cohorts (NMEDW and eICU).

**Appendix 4 The detailed analysis in terms of physiological characteristics**
Patients physiological characteristics mainly covered inflammation, pulmonary, cardiovascular/Hemodynamic, renal, hepatic, neurological, and hematologic. Specifically, vital signs including temperature, respiratory rate, heart rate, GCS, systolic ABP, and BMI. Lab tests including bands, C-reactive protein, WBC, SO2, PaO2, Bicarbonate, lactate, troponin, BUN, creatinine, ALT, AST, bilirubin, hemoglobin, INR, platelets, albumin, chloride, glucose, sodium, lymphocyte count, RDW, and lymphocyte percentage. We screened the vital signs and lab tests from their values in the first 6 hours to compare these subphenotypes. For the development cohort, the significant lab tests included ALT, AST, bicarbonate, bilirubin, BUN, chloride, creatinine, hemoglobin, INR, lactate, platelet, SO2, WBC, RDW. The significant vital signs included respiratory rate, heart rate, systolic ABP, temperature. Patients in Rapidly Worsening had the highest ALT, AST, bilirubin, heart rate, INR, lactate and the lowest bicarbonate and platelet. Patients in Rapidly Improving had the highest hemoglobin, platelet, WBC, temperature and the lowest heart rate, RDW, SO2, and systolic ABP. More information was shown in Table S6. For the NMEDW validation dataset, the significant lab tests and vital signs include systolic ABP, platelet, RDW, AST, GCS, respiratory rate, WBC. For the eICU validation dataset, the significant lab tests and vital signs include Albumin, ALT, AST, Bicarbonate, Bilirubin, BMI, BUN, Chloride, Creatinine, GCS, Glucose, Heart rate, INR, Lactate, Lymphocyte, Platelet, RDW, respiratory rate, SO2, systolic ABP, and WBC. For the CEDAR validation dataset, the significant lab tests and vital signs include Albumin, Bicarbonate, BUN, Creatinine, GCS, Glucose, Heart rate, Hemoglobin, INR, Lactate, Platelet, Respiratory rate, SO2, systolic ABP, Temperature, and WBC. More details are shown in Tables S8, S10 and S12.

**Appendix 5 Chord plots**
Chord plots were used to visualize the patterns of abnormal clinical variables and six sub-SOFA scores by subphenotype, which was obtained using the "Circlize" package in R. These clinical variables were divided into several groups. In particular,

- ❖ Inflammatory markers: temperature, WBC, bands, CRP, albumin, lymphocyte percent.
- ❖ Hepatic markers: bilirubin, AST, ALT.
- ❖ Cardiovascular markers: heart rate, systolic ABP, bicarbonate, troponin T or I, lactate.
- ❖ Renal markers: creatinine, BUN, chloride, sodium.
- ❖ Hematologic markers: hemoglobin, INR, platelet, glucose, RDW.
- ❖ Pulmonary markers: respiratory rate, SO2, Pao2.
- ❖ Neurologic marker: GCS.
- ❖ Comorbidity: Comorbidity score based on Elixhauser.

For each subphenotype, if a clinical variable median value (e.g., AST) is greater than the median of the development cohort, a ribbon is connected between the subphenotype and the group (e.g., Hepatic markers). Subphenotypes are demonstrated in different colors. If there are more clinical variables abnormal for that subphenotype, the ribbon would be more broader.

**Appendix 6 The determination of the number of optimal clusters**

Multiple clustering indexes were used to determine the optimal numbers of subphenotypes. We used 25 clustering index from Nbclust [2] to choose the number of clusters. The optimal number of clusters for each clustering index was shown in Table S5. Among all indices in development cohort:

- 6 indices proposed 2 as the best number of clusters;
- 3 indices proposed 3 as the best number of clusters;
- 11 indices proposed 4 as the best number of clusters;
- 3 indices proposed 5 as the best number of clusters;
- 2 indices proposed 6 as the best number of clusters.

Among all indices in NMEDW validation cohort:

- 9 indices proposed 2 as the best number of clusters;
- 3 indices proposed 3 as the best number of clusters;
- 10 indices proposed 4 as the best number of clusters;
- 1 indice proposed 5 as the best number of clusters;
- 2 indices proposed 6 as the best number of clusters.

Among all indices in eICU validation cohort:

- 3 indices proposed 2 as the best number of clusters;
- 4 indices proposed 3 as the best number of clusters;
- 12 indices proposed 4 as the best number of clusters;
- 3 indices proposed 5 as the best number of clusters;
- 3 indices proposed 6 as the best number of clusters.

Among all indices in CEDAR validation cohort:

- 0 indices proposed 2 as the best number of clusters;
- 7 indices proposed 3 as the best number of clusters;
- 14 indices proposed 4 as the best number of clusters;
- 2 indices proposed 5 as the best number of clusters;
- 2 indices proposed 6 as the best number of clusters.

According to the majority rule, the best number of clusters was set as 4 in experiment.

**Appendix 7 Dynamic Time Warping (DTW) and Hierarchical Agglomerative Clustering (HAC)**

Dynamic time warping (DTW) is one of most popular techniques to measure the similarity between two temporal sequences [3]. In DTW, the warping technique of one-to-many match is used to obtain the optimal match of two sequences so that the troughs and peaks with the same pattern are completely matched, and there is no left out for both temporal sequences. Once the matched sequences are obtained, the Euclidean Distance is used to compute the distance of two sequences. Due to its invariance against warping in the time axis, DTW has been widely utilized as a similarity measure in various domains such as speech recognition and provides more meaningful discrepancy measurements between two temporal sequences than other distance measures [4]. Previous clinical study in terms of COVID-19 has shown the DTW can be used to assess longitudinal changes in organ dysfunction and help clinicians or doctors to obtain important insights about pathophysiology [8].

Technically, DTW is a technique that compares sequences A and B through matching, where for every point in A it finds a best matching point in B. The distance between A and B is the sum of the distances between all matched point pairs. Mathematically, let $x^i = \{x_1^i, x_2^i, \ldots, x_{T_i}^i\}$ be the sequence of the *i-th* patient (1≤ $i$ ≤N). $T_i$ is the sequence length and $x_k^i$ is its value at the *k-th* timestamp. In particular, given a sequence pair $(x^i, x^j)$, (see Figure

S19) for every data point $x_k^i \in x^i$ $(1 \leq k \leq T_i)$, DTW identifies a matching point $x_{k'}^j \in x^j$ $(1 \leq k' \leq T_j)$ and according to some ground distance metric $G(x_k^i, x_{k'}^j)$, then the distance of the two sequence, $D(x^i, x^j) = \sum_{k,k'} G(x_k^i, x_{k'}^j)$. The matching process needs to satisfy 1) if $k = 1$, then $k' = 1$, which means the first points of the two sequences should match each other; 2) if $k = T_i$, then $k' = T_j$, which means the last points of the two sequences should match each other; 3) the matching should be monotonic, i.e., if $(x_{k_1}^i, x_{k'_1}^j)$and $(x_{k_2}^i, x_{k'_2}^j)$ are two matched pairs and $k_1 < k_2$, then $k'_1 \leq k'_2$. DTW matches $(x^i, x^j)$ by minimizing the sum of differences (measured by ground distance) between the all matched point pairs on the two sequences through dynamic programming [14]. After collecting all matched point pairs from $x^i$ and $x^j$, we get the warping path $\mathcal{P}(x^i, x^j) = \{(x_k^i, x_{k'}^j)\}_{k,k'=1}^{T_i,T_j}$, and the number of entries in $\mathcal{P}(x^i, x^j)$ is its length. We can index the entries in $\mathcal{P}(x^i, x^j)$ according to the orders they appear in the path as $\mathcal{P}(x^i, x^j) = \{p_1^{ij}, p_2^{ij}, \ldots, p_{L_{ij}}^{ij}\}$ with $p_1^{ij} = (x_1^i, x_1^j)$ and $p_{L_{ij}}^{ij} = (x_{T_i}^i, x_{T_j}^j)$, and $L_{ij}$ is the path length satisfying $\max(T_i, T_j) \leq L_{ij} \leq T_i + T_j$.

In this study, after obtaining the SOFA trajectories of patients (see the Figure S20, the illustration of SOFA trajectory), each patient is represented as a vector of 12 SOFA scores from the first 6 hours to the last 6 hours across the 72 hours period after admission in ICU. DTW was used to evaluate the similarities between pairwise patient SOFA trajectories. Hierarchical Agglomerative Clustering (HAC) [13] is used to obtain subphenotypes based on the similarities of SOAF score trajectories derived by DTW. Compared to other clustering methods such as k-means, HAC has two advantages: (1) HAC is usually robust as it is not sensitive to data distribution and not necessary to have initialization procedure, which may cause uncertainty. (2) HAC typically generates a tree diagram that can visually demonstrate how the data points are agglomerated together in a hierarchical manner and provides a visible way to assistance the determination of optimal cluster number. In order to furtherly determine the number of optimal cluster number, one famous R package “NbClust” [2] was used in this study. During using the “NbClust”, we considered 25 indices. The more details in terms of these indices were shown in Table S5.

**Appendix 8 Group-Based Trajectory Modeling (GBTM)**
In order to assess the sensitivity to different clustering methods, in this study, we used GBTM to re-derive subphenotypes. The GBTM is one of the most popular latent class analysis (LCA) that assigns each patient a probability of being in each subphenotype by considering maximum likelihood estimation [5]. The GBTM has been widely used for researchers to identify underlying subgroups in terms of disease courses such as coronary artery risk development in young adults [6] [7]. During using GBTM to derive subphenotypes, the optimal subphenotype number was determined by comprehensively taking Akaike information criterion (AIC) [11] and Bayesian information criterion (BIC) [12] into account.

**Table S1. Patient characteristics comparisons between survivors and nonsurvivors in the development cohort**

| Characteristics | Total<br>N=4,678 | Survivors<br>N=4,170 | Nonsurvivors<br>N=508 | p-value† |
|---|---|---|---|---|
| **Age, median (IQR)** | 65.9 [53.7-77.9] | 65.2 [53.2-77.4] | 71.5 [59.9-80.9] | <0.001 |
| **Sex, No. (%)** | | | | 0.418 |
| Male | 2625 (56.1%) | 2349 (56.3%) | 276 (54.3%) | |
| Female | 2053 (43.9%) | 1821 (43.7%) | 232 (45.7%) | |
| **Race, No. (%)** | | | | <0.001 |
| WHITE | 3367 (71.9%) | 3022 (72.5%) | 345 (67.9%) | |
| BLACK | 424 (9.1%) | 396 (9.5%) | 28 (5.5%) | |
| OTHER | 887 (18.9%) | 752 (18.0%) | 135 (26.6%) | |
| **Elixhauser index, median (IQR)** | 4.0 [0.0-9.0] | 4.0 [0.0-8.0] | 7.0 [2.0-12.0] | <0.001 |
| **Length stay, median (IQR)** | 2.8 [1.6-5.6] | 2.7 [1.6-5.2] | 3.9 [1.9-7.7] | <0.001 |
| **Mechanical ventilation at admission, No. (%)** | 1893 (40.5%) | 1589 (38.1%) | 304 (59.8%) | <0.001 |
| **Baseline SOFA, mean (SD)** | 4.96 (2.8) | 4.7 (2.6) | 7.1 (3.7) | <0.001 |
| **ICU unit at admission, No. (%)** | | | | <0.001 |
| CCU | 443 (9.5%) | 377 (9.0%) | 66 (12.9%) | |
| CSRU | 260 (5.6%) | 253 (6.1%) | 7 (1.4%) | |
| MICU | 2611 (55.8%) | 2293 (54.9%) | 318 (62.6%) | |
| TSICU | 593 (12.7%) | 547 (13.1%) | 46 (9.1%) | |
| SICU | 771 (16.4%) | 700 (16.7%) | 71 (13.9%) | |
| **Admission location, No. (%)** | | | | <0.001 |
| Transfer from other hospital | 4 (0.1%) | 3 (0.1%) | 1 (0.2%) | |
| Emergency room | 1497 (32.0%) | 1335 (32.0%) | 162 (31.8%) | |
| Clinic referral | 1985 (42.4%) | 1779 (42.6%) | 206 (40.5%) | |
| Transfer from ward | 810 (17.3%) | 686 (16.4%) | 124 (24.4%) | |
| Physician referral | 367 (7.8%) | 355 (8.5%) | 12 (2.3%) | |
| Transfer from skilled nursing facility | 15 (0.3%) | 12 (0.2%) | 3 (0.5%) | |
| **Infection item** | | | | |
| Central nervous system | 56 (1.2%) | 54 (1.2%) | 2 (0.3%) | 0.122 |
| Intra-abdominal | 880 (18.8%) | 794 (19.0%) | 86 (16.9%) | 0.276 |
| Pneumonia | 1257 (26.8%) | 1108 (26.5%) | 149 (29.3%) | 0.203 |
| Septicemia bacteremia | 1587 (33.9%) | 1331 (31.9%) | 256 (50.3%) | <0.001 |
| Skin soft tissue | 276 (5.9%) | 263 (6.3%) | 13 (2.5%) | <0.001 |
| Urinary tract | 1044 (22.3%) | 974 (23.3%) | 70 (13.7%) | <0.001 |

Definition of abbreviations: IQR--interquartile range; SD--standard deviation; SOFA--Sequential Organ Failure Assessment; SICU--Surgical ICU; CCU-- Coronary Care Unit; TSICU--Thoracic Surgery ICU; MICU--Medical ICU; CSRU--Cardiac Surgery ICU.
†p-value calculated by Chi-square test/Fisher's exact test, or student's t-test/Mann-Whitney test where appropriate.

**Table S2. Patient characteristics comparisons between survivors and nonsurvivors in the NMEDW validation cohort**

| Characteristics | Total N=3,665 | Survivors N=3,107 | Nonsurvivors N=558 | p-value† |
|---|---|---|---|---|
| **Age, median (IQR)** | 65.0(54.0-76.0) | 65.0(54.0-75.0) | 69.0(59.0-78.0) | <0.001 |
| **Sex, No. (%)** | | | | 0.877 |
| Male | 2,011(54.8%) | 1,707(54.9%) | 304(54.4%) | |
| Female | 1,654(45.1%) | 1,400(45.0%) | 254(45.5%) | |
| **Race, No. (%)** | | | | 0.254 |
| WHITE | 2,596(70.8%) | 2,202(70.8%) | 394(70.6%) | |
| BLACK | 526(14.3%) | 455(14.6%) | 71(12.7%) | |
| OTHER | 543(14.8%) | 450(14.4%) | 93(16.6%) | |
| **Length stay, median (IQR)** | 3.8(1.9-7.9) | 3.7(1.9-7.6) | 5.0(2.6-9.6) | <0.001 |
| **Mechanical ventilation, No. (%)** | 1,524(41.5%) | 1,197(38.5%) | 327(58.6%) | <0.001 |
| **Elixhauser index, median (IQR)** | 6.0(0.0-14.0) | 6.0(0.0-14.0) | 9.0(0.0-18.0) | <0.001 |
| **Baseline SOFA, mean (SD)** | 5.6(2.8) | 5.4(2.6) | 7.7(4.0) | <0.001 |
| **Infection item** | | | | |
| Central nervous system | 47(1.2%) | 41(1.3%) | 6(1.0%) | 0.789 |
| Intra-Abdominal | 383(10.4%) | 321(10.3%) | 62(11.1%) | 0.632 |
| Pneumonia | 980(26.7%) | 832(26.7%) | 148(26.5%) | 0.942 |
| Septicemia bacteremia | 1293(35.2%) | 1034(33.2%) | 259(46.4%) | <0.001 |
| Skin soft tissue | 157(4.2%) | 146(4.7%) | 11(1.9%) | 0.005 |
| Urinary tract | 410(11.1%) | 367(11.8%) | 43(7.7%) | 0.006 |

Definition of abbreviations: IQR--interquartile range; SD--standard deviation; SOFA--Sequential Organ Failure Assessment.
†p-value calculated by Chi-square test/Fisher’s exact test, or student’s t-test/Mann-Whitney test where appropriate.

**Table S3. Patient characteristics comparisons between survivors and nonsurvivors in the eICU validation cohort**

| Characteristics | Total N=12,282 | Survivors N=10,995 | Nonsurvivors N=1,287 | p-value† |
|---|---|---|---|---|
| **Age, median (IQR)** | 67.0 [56.0-78.0] | 67.0 [55.0-78.0] | 71.0 [60.5-81.0] | <0.001 |
| **Sex, No. (%)** | | | | 0.517 |
| Male | 6438 (52.4%) | 5782 (52.5%) | 656 (50.9%) | |
| Female | 5844 (47.5%) | 5213 (47.4%) | 631 (49.0%) | |
| **Ethnicity, No. (%)** | | | | 0.571 |
| Caucasian | 9434 (76.8%) | 8428 (76.6%) | 1006 (78.1%) | |
| African American | 1317 (10.7%) | 1198 (10.9%) | 119 (9.2%) | |
| Hispanic | 593 (4.8%) | 528 (4.8%) | 65 (5.0%) | |
| Native American | 111 (0.9%) | 99 (0.9%) | 12 (0.9%) | |
| Asian | 201 (1.6%) | 178 (1.6%) | 23 (1.7%) | |
| Other/Unknown | 626 (5.1%) | 564 (5.1%) | 62 (4.8%) | |
| **Elixhauser index, median (IQR)** | 5.0 [0.0-12.0] | 3.0 [0.0-12.0] | 7.0 [0.0-18.0] | <0.001 |
| **Length stay, median (IQR)** | 2.8 [1.7-5.1] | 2.8 [1.7-4.9] | 3.18 [1.6-6.8] | 0.002 |
| **Mechanical ventilation at admission, No. (%)** | 5329 (43.3%) | 4587 (41.7%) | 742 (57.6%) | <0.001 |
| **Baseline SOFA, mean (SD)** | 5.9 (3.0) | 5.66 (2.8) | 8.72 (3.7) | <0.001 |
| **ICU unit at admission, No. (%)** | | | | 0.042 |
| SICU | 627 (5.1%) | 558 (5.0%) | 69 (5.3%) | |
| MICU | 1575 (12.8%) | 1402 (12.7%) | 173 (13.4%) | |
| CTICU | 87 (0.7%) | 82 (0.7%) | 5 (0.3%) | |
| CSICU | 160 (1.3%) | 138 (1.2%) | 22 (1.7%) | |
| MSICU | 7797 (63.4%) | 6986 (63.5%) | 811 (63.0%) | |
| CCU-CTICU | 772 (6.2%) | 684 (6.2%) | 88 (6.8%) | |
| CICU | 907 (7.3%) | 808 (7.3%) | 99 (7.6%) | |
| Neuro-ICU | 357 (2.9%) | 337 (3.0%) | 20 (1.5%) | |
| **Admission location, No. (%)** | | | | <0.001 |
| Floor | 1986 (16.1%) | 1738 (15.8%) | 248 (19.2%) | |
| SDU | 218 (1.7%) | 176 (1.6%) | 42 (3.2%) | |
| Direct Admit | 852 (6.9%) | 760 (6.9%) | 92 (7.1%) | |
| Other Hospital | 404 (3.2%) | 345 (3.1) % | 59 (4.5) % | |
| ICU | 4 (0.03%) | 4 (0.04%) | 0 (0.0%) | |
| Other ICU | 63 (0.5%) | 51 (0.4%) | 12 (0.9%) | |
| ICU to SDU | 1 (0.01%) | 1 (0.01%) | 0 (0.0%) | |
| PACU | 17 (0.14%) | 16 (0.15%) | 1 (0.08%) | |
| Recovery Room | 146 (1.1%) | 138 (1.2%) | 8 (0.6%) | |
| Chest Pain Center | 5 (0.04%) | 5 (0.05%) | 0 (0.00%) | |
| Emergency Department | 7825 (63.7%) | 7073 (64.3%) | 752 (58.4%) | |
| Acute Care/Floor | 284 (2.3%) | 247 (2.2%) | 37 (2.8%) | |
| Operating Room | 477 (3.8%) | 441 (4.0%) | 36 (2.8%) | |
| **Infection item, No. (%)** | | | | |
| Central nervous system | 63 (0.5%) | 58 (0.5%) | 5 (0.3%) | 0.649 |
| Intra-abdominal | 790 (6.4%) | 689 (6.2%) | 101 (7.8%) | 0.033 |

| | | | | |
|---|---|---|---|---|
| Pneumonia | 4743 (38.6%) | 4176 (37.9%) | 567 (44.0%) | <0.001 |
| Septicemia bacteremia | 7350 (59.8%) | 6460 (58.7%) | 890 (69.1%) | <0.001 |
| Skin soft tissue | 632 (5.1%) | 590 (5.3%) | 42 (3.2%) | <0.001 |
| Urinary tract | 2478 (20.1%) | 2279 (20.7%) | 199 (15.4%) | <0.001 |

Definition of abbreviations: IQR--interquartile range; SD--standard deviation; SOFA--Sequential Organ Failure Assessment; SICU-- Surgical ICU; MICU--Medical ICU; CTICU--Cardiothoracic ICU; CSICU--Cardiac Surgery ICU; MSICU--Medical-Surgical ICU; CCU--Coronary Care Unit; CICU-- Cardiac ICU; Neuro-ICU-- Neurological ICU; SDU--Step-Down Unit.

†p-value calculated by Chi-square test/Fisher's exact test, or student's t-test/Mann-Whitney test where appropriate.

**Table S4. Patient characteristics comparisons between survivors and nonsurvivors in the CEDAR validation cohort**

| Characteristics | Total N=4,804 | Survivors N=3,850 | Nonsurvivors N=954 | p-value† |
|---|---|---|---|---|
| **Age, median (IQR)** | 77.0 [66.0-88.0] | 77.0 [65.0-88.0] | 78.0 [68.0-89.0] | 0.004 |
| **Sex, No. (%)** | | | | 0.133 |
| Male | 2320 (48.2%) | 1838 (47.7%) | 482 (50.5%) | |
| Female | 2484 (51.7%) | 2012 (52.2%) | 472 (49.4%) | |
| **Race, No. (%)** | | | | 0.238 |
| WHITE | 1791 (37.2%) | 1420 (36.8%) | 371 (38.8%) | |
| BLACK | 446 (9.2%) | 350 (9.0%) | 96 (10.0%) | |
| OTHER | 2567 (53.4%) | 2080 (54.0%) | 487 (51.0%) | |
| **Length stay, median (IQR)** | 4.42(2.6-7.9) | 4.2(2.6-7.7) | 5.0(2.5-9.4) | 0.026 |
| **Mechanical ventilation, No. (%)** | 2263(47.1%) | 1624(42.1%) | 639(66.9%) | <0.001 |
| **Elixhauser index, median (IQR)** | 12.5 [0.0-27.0] | 11.0 [0.0-25.0] | 21.0 [0.0-33.0] | <0.001 |
| **Baseline SOFA, mean (SD)** | 6.38 (3.1) | 5.8 (2.8) | 8.43 (3.40) | <0.001 |
| **ICU unit at admission, No. (%)** | | | | <0.001 |
| CTICU | 656 (13.6%) | 627 (16.2%) | 29 (3.0%) | |
| SICU | 752 (15.6%) | 658 (17.0%) | 94 (9.8%) | |
| MICU | 2641 (54.9%) | 1976 (51.3%) | 665 (69.7%) | |
| CICU | 755 (15.7%) | 589 (15.3%) | 166 (17.4%) | |
| **Infection item** | | | | |
| Central nervous system | 38 (0.7%) | 30 (0.7%) | 8 (0.8%) | 0.985 |
| Intra-Abdominal | 499 (10.3%) | 375 (9.7%) | 124 (13.0%) | 0.004 |
| Pneumonia | 597 (12.4%) | 431 (11.1%) | 166 (17.4%) | <0.001 |
| Septicemia bacteremia | 1376 (28.6%) | 1001 (26.0%) | 375 (39.3%) | <0.001 |
| Skin soft tissue | 164 (3.4%) | 137 (3.5%) | 27 (2.8%) | 0.312 |
| Urinary tract | 1127 (23.4%) | 977 (25.3%) | 150 (15.7%) | <0.001 |

Definition of abbreviations: IQR--interquartile range; SD--standard deviation; SOFA--Sequential Organ Failure Assessment; CTICU--Cardiothoracic ICU; SICU-- Surgical ICU; MICU-- Medical ICU; CICU-- Cardiac ICU.
†p-value calculated by Chi-square test/Fisher's exact test, or student's t-test/Mann-Whitney test where appropriate.

**Table S5. Multiple clustering metrics statistics and cluster number determination**

| Clustering Metrics | Optimal Cluster Number in Development Cohort | Optimal Cluster Number in the NMEDW Validation Cohort | Optimal Cluster Number in the eICU Validation Cohort | Optimal Cluster Number in the CEDAR Validation Cohort |
|---|---|---|---|---|
| **KL** | 4 | 4 | 4 | 4 |
| **CH** | 2 | 2 | 4 | 4 |
| **Hartigan** | 6 | 4 | 6 | 4 |
| **CCC** | 2 | 2 | 2 | 3 |
| **Scott** | 4 | 4 | 4 | 4 |
| **Marriot** | 4 | 4 | 4 | 4 |
| **TrCovW** | 4 | 4 | 4 | 4 |
| **TraceW** | 4 | 4 | 4 | 4 |
| **Friedman** | 4 | 4 | 4 | 4 |
| **Rubin** | 4 | 4 | 4 | 4 |
| **Cindex** | 5 | 6 | 5 | 4 |
| **DB** | 5 | 3 | 3 | 3 |
| **Silhouette** | 2 | 2 | 2 | 3 |
| **Duda** | 2 | 2 | 5 | 5 |
| **PseudoT2** | 3 | 3 | 5 | 5 |
| **Beale** | 2 | 2 | 3 | 3 |
| **Ratkowsky** | 6 | 2 | 4 | 4 |
| **Ball** | 3 | 3 | 4 | 4 |
| **PtBiserial** | 4 | 4 | 3 | 3 |
| **Gap** | 2 | 2 | 4 | 4 |
| **Frey** | 4 | 2 | 4 | 4 |
| **McClain** | 4 | 6 | 3 | 3 |
| **Dunn** | 4 | 4 | 6 | 6 |
| **SDindex** | 3 | 2 | 2 | 3 |
| **SDbw** | 5 | 5 | 6 | 6 |
| **Total** | # of 2: 6<br># of 3: 3<br>**# of 4: 11**<br># of 5: 3<br># of 6: 2 | # of 2: 9<br># of 3: 3<br>**# of 4: 10**<br># of 5: 1<br># of 6: 2 | # of 2: 3<br># of 3: 4<br>**# of 4: 12**<br># of 5: 3<br># of 6: 3 | # of 2: 0<br># of 3: 7<br>**# of 4: 14**<br># of 5: 2<br># of 6: 2 |

Definition of abbreviations: KL-- Krzanowski-Lai index; CH-- Calinski-Harabasz index; CCC-- Cubic Clustering Criterion, DB-- Davies–Bouldin index; All those clustering metrics are from NbClust Package.

**Table S6. Patient physiological characteristics comparisons among subphenotypes in the development cohort**

| Characteristics | Total (N=4,678) | DI (N=1,174) | RI (N=1,932) | DW (N=960) | RW (N=612) | p-value† | Post-hoc |
|---|---|---|---|---|---|---|---|
| Albumin, g/dL, median [IQR] | 2.9 [2.5-3.3] | 2.9 [2.5-3.32] | 3.0 [2.5-3.3] | 2.9 [2.5-3.3] | 2.9 [2.4-3.3] | 0.346 | |
| ALT, IU/L, median [IQR] | 35.0 [19.0-86.0] | 33.0 [19.0-85.75] | 36.5 [20.0-89.25] | 30.0 [18.0-71.0] | 40.0 [21.0-86.25] | 0.016 | DW vs RI, RW |
| AST, IU/L, median [IQR] | 49.0 [27.0-120.0] | 48.0 [28.0-125.0] | 49.0 [27.0-115.2] | 45.0 [24.0-106.0] | 61.0 [32.5-143.5] | <0.001 | DI vs DW, RW; RW vs RI, DW |
| Bands, median [IQR] | 2.0 [0.0-8.0] | 2.0 [0.0-9.0] | 2.0 [0.0-9.0] | 2.0 [0.0-8.0] | 2.0 [0.0-8.0] | 0.712 | |
| Bicarbonate, mEq/L, median [IQR] | 23.0 [20.0-26.0] | 24.0 [21.0-26.0] | 23.0 [20.0-25.0] | 23.0 [19.0-26.0] | 22.0 [19.0-26.0] | <0.001 | DI vs RI, DW, RW |
| Bilirubin, mg/dL, median [IQR] | 0.8 [0.4-2.1] | 1.0 [0.5-2.4] | 0.8 [0.4-2.0] | 0.7 [0.4-1.6] | 1.1 [0.5-2.5] | <0.001 | DI vs RI, DW; RI vs DW; RW vs RI, DW |
| BMI, kg/m2, median [IQR] | 27.5 [23.7-32.2] | 27.4 [23.7-32.4] | 27.6 [23.9-32.5] | 27.9 [23.9-32.4] | 26.8 [23.5-30.8] | 0.227 | |
| BUN, mg/dL, median [IQR] | 22.0 [15.0-38.0] | 20.0 [14.0-32.0] | 22.0 [15.0-37.0] | 27.0 [16.0-47.0] | 23.0 [15.0-38.0] | <0.001 | DI vs RI, DW, RW; RI vs DW; DW vs RW |
| Chloride, mEq/L, median [IQR] | 106.0 [102.0-110.0] | 106.0 [101.0-109.0] | 106.0 [102.0-110.0] | 106.0 [102.0-110.0] | 106.0 [101.0-110.0] | 0.014 | DI vs RI; RI vs RW |
| Creatinine, mg/dL, median [IQR] | 1.1 [0.8-1.7] | 1.0 [0.7-1.4] | 1.1 [0.8-1.7] | 1.3 [0.9-2.4] | 1.1 [0.8-1.7] | <0.001 | DI vs RI, DW, RW; RI vs DW; DW vs RW |
| CRP, mg/dL, median [IQR] | 92.5 [40.5-174.4] | 107.0 [51.4-181.6] | 81.5 [42.2-172.1] | 82.6 [29.25-177.15] | 72.5 [41.7-143.7] | 0.825 | |
| GCS, median [IQR] | 13.0 [7.0-15.0] | 13.0 [7.0-15.0] | 13.0 [7.0-15.0] | 14.0 [7.0-15.0] | 12.0 [6.0-15.0] | 0.203 | |
| Glucose, mg/dL, median [IQR] | 131.0 [106.0-172.0] | 130.0 [106.0-167.75] | 132.0 [107.0-173.0] | 131.0 [106.0-174.0] | 134.0 [105.0-176.0] | 0.746 | |
| Heart rate, beats/min, median [IQR] | 95.0 [81.0-110.0] | 96.0 [82.0-110.0] | 93.42 [80.5-108.3] | 94.12 [80.0-110.2] | 96.83 [84.0-112.0] | 0.002 | DI vs RI, RI vs RW, DW vs RW |
| Hemoglobin, g/dL | 10.6 [9.2-12.0] | 10.57 [9.3-11.9] | 10.7 [9.3-12.1] | 10.3 [9.1-11.8] | 10.6 [9.07-12.1] | 0.041 | RI vs DW |
| INR, median [IQR] | 1.3 [1.2-1.6] | 1.3 [1.2-1.6] | 1.3 [1.1-1.5] | 1.3 [1.2-1.6] | 1.4 [1.2-1.7] | <0.001** | DI, RI, DW vs RW |
| Lactate, mmol/L, median [IQR] | 1.8 [1.2-2.8] | 1.8 [1.2-2.8] | 1.7 [1.2-2.7] | 1.7 [1.2-2.7] | 2.0 [1.3-3.3] | <0.001§ | DI, RI, DW vs RW |
| lymphocyte count, median [IQR] | 940.0 [675.0-1570.0] | 1278.0 [775.0-1854.5] | 805.0 [539.2-1172.0] | 1033.5 [866.2-1792.5] | 780.0 [460.0-1690.0] | 0.262 | |
| lymphocyte percentage, %, median [IQR] | 7.1 [3.5-12.5] | 7.5 [4.0-12.6] | 7.2 [3.5-12.4] | 6.8 [3.3-12.0] | 6.75 [3.5-13.5] | 0.283 | |
| Pao2, mm Hg | 108.0 [81.5-168.5] | 101.5 [86.5-109.5] | 110.0 [70.0-157.0] | 102.5 [83.7-162.7] | 170.0 [90.0-213.5] | 0.364 | |
| Platelet, x10(9)/L, median [IQR] | 190.0 [130.0-260.0] | 186.0 [125.0-258.2] | 199.0 [138.75-268.0] | 190.0 [132.0-252.7] | 168.0 [112.75-237.0] | <0.001 | DI vs RI, DW; RI vs DW, RW; DW vs RW |
| RDW, %, median [IQR] | 14.8 [13.8-16.3] | 14.8 [13.8-16.2] | 14.6 [13.6-16.0] | 15.0 [13.9-16.6] | 14.9 [13.8-16.8] | <0.001 | DI vs RI; RI vs DW, RW |
| Respiratory rate, breaths/min, median [IQR] | 22.0 [19.0-27.0] | 23.0 [19.0-27.0] | 22.0 [19.0-26.0] | 22.0 [19.0-26.75] | 22.33 [19.0-27.0] | 0.023 | DI vs RI; RI vs RW |
| SO2, %, median [IQR] | 94.0 [80.0-97.0] | 94.0 [83.0-97.0] | 93.0 [78.0-97.0] | 95.0 [78.38-97.25] | 94.0 [82.0-98.0] | 0.014 | DI vs RI |

| | | | | | | | |
|---|---|---|---|---|---|---|---|
| **Sodium, mmol/L, median [IQR]** | 139.0 [136.0-142.0] | 139.0 [136.0-141.0] | 139.0 [136.0-141.0] | 139.0 [136.0-142.0] | 139.0 [136.0-142.0] | 0.973 | |
| **Systolic ABP, mm Hg, median [IQR]** | 101.0 [91.0-115.0] | 102.0 [92.0-118.0] | 100.0 [91.0-112.0] | 102.0 [91.7-115.0] | 102.0 [91.0-117.0] | <0.001 | DI vs RI; RI vs DW, RW; |
| **Temperature, degrees C, median [IQR]** | 36.8 [36.3-37.4] | 36.8 [36.3-37.4] | 36.8 [36.3-37.5] | 36.8 [36.2-37.4] | 36.78 [36.2-37.3] | 0.030 | RI vs DW, RW |
| **Troponin T, ng/mL, median [IQR]** | 0.1 [0.0-0.3] | 0.1 [0.0-0.3] | 0.1 [0.0-0.3] | 0.1 [0.0-0.2] | 0.1 [0.0-0.3] | 0.404 | |
| **WBC, x10(9)/L, median [IQR]** | 11.1 [7.5-15.8] | 10.6 [7.1-14.9] | 11.4 [7.9-16.4] | 10.9 [7.4-15.3] | 11.0 [7.3-16.2] | <0.001 | DI vs RI, RI vs DW |

Definition of abbreviations: IQR--interquartile range; SD--standard deviation; ALT-- alanine aminotransferase; AST-- alanine aminotransferase; BMI-- body mass index, BUN-- blood urea nitrogen; CRP-- C-reactive protein; GCS-- Glasgow coma scale score; INR-- international normalized ratio; RDW-- red blood cell distribution width; ABP-- Ambulatory Blood Pressure; WBC--white blood cell count; DI-- Delayed Improving; RI-- Rapidly Improving; DW-- Delayed Worsening; RW-- Rapidly Worsening.
† p-value calculated by analysis of variance (ANOVA)/Kruskal–Wallis test.
** False discovery rate corrected p-value < 0.001
§ Age adjusted (analysis of covariance [ANCOVA]) p-value < 0.001

**Table S7. Patient basic characteristics comparisons among subphenotypes in the NMEDW validation cohort**

| Characteristics | Total N=3665 | DI N=668 | RI N=2,019 | DW N=302 | RW N=676 | p-value[†] |
|---|---|---|---|---|---|---|
| **Age, median (IQR)** | 65.0(54.0-76.0) | 66.0(55.0-77.0) | 66.0(55.0-76.0) | 66.0(54.2-76.7) | 64.0(52.0-74.0) | 0.027 |
| **Sex, No. (%)** | | | | | | 0.463 |
| Male | 2011(54.8%) | 366(54.7%) | 1116(55.2%) | 174(57.6%) | 355(52.5%) | |
| Female | 1654(45.1%) | 302(45.2%) | 903(44.7%) | 128(42.3%) | 321(47.4%) | |
| **Race, No. (%)** | | | | | | <0.001 |
| WHITE | 2596(70.8%) | 472(70.6%) | 1481(73.3%) | 205(67.8%) | 438(64.7%) | |
| BLACK | 526(14.3%) | 93(13.9%) | 281(13.9%) | 42(13.9%) | 110(16.2%) | |
| OTHER | 543(14.8%) | 103(15.4%) | 257(12.7%) | 55(18.2%) | 128(18.9%) | |
| **Length stay, median (IQR)** | 3.8(1.9-7.9) | 4.4(2.0-9.1) | 3.2(1.8-6.1) | 4.2(2.1-9.2) | 5.44(2.3-11.5) | <0.001 |
| **Mechanical ventilation at admission, No. (%)** | 1524(41.5%) | 252(37.7%) | 882(43.6%) | 111(36.7%) | 279(41.2%) | 0.014 |
| **Elixhauser index, median (IQR)** | 6.0(0.0-14.0) | 7.00(0.0-15.0) | 5.0(0.0-12.0) | 8.0(0.0-17.0) | 9.00(0.0-17.0) | <0.001 |
| **Baseline SOFA, mean (SD)** | 5.6(2.8) | 4.71(2.3) | 6.1(2.9) | 6.0(2.8) | 5.22(2.5) | <0.001 |
| **Infection item, No. (%)** | | | | | | |
| Central nervous system | 47(1.2%) | 11(1.6%) | 18(0.8%) | 7(2.3%) | 11(1.6%) | 0.097 |
| Intra-Abdominal | 383(10.4%) | 68(10.1%) | 198(9.8%) | 36(11.9%) | 81(11.9%) | 0.342 |
| Pneumonia | 980(26.7%) | 207(30.9%) | 478(23.6%) | 81(26.8%) | 214(31.6%) | <0.001 |
| Septicemia bacteremia | 1293(35.2%) | 227(33.9%) | 667(33.0%) | 124(41.0%) | 275(40.6%) | <0.001 |
| Skin soft tissue | 157(4.2%) | 24(3.5%) | 93(4.6%) | 16(5.3%) | 24(3.5%) | 0.402 |
| Urinary tract | 410(11.1%) | 91(13.6%) | 224(11.0%) | 37(12.2%) | 58(8.5%) | 0.030 |
| **Septic shock, No. (%)** | 366(9.9%) | 54(8.0%) | 229(11.3%) | 36(11.9%) | 47(6.9%) | 0.002 |

Definition of abbreviations: IQR--interquartile range; SD--standard deviation; SOFA--Sequential Organ Failure Assessment; DI-- Delayed Improving; RI--Rapidly Improving; DW-- Delayed Worsening; RW-- Rapidly Worsening.

[†]p-value calculated by Chi-square test/Fisher's exact test, or student's t-test/Mann-Whitney test where appropriate.

**Table S8. Patient physiological characteristics comparisons among subphenotypes in the NMEDW validation cohort**

| Characteristics | Total N=3665 | DI N=668 | RI N=2,019 | DW N=302 | RW N=676 | p-value† | Post-hoc |
|---|---|---|---|---|---|---|---|
| **Albumin, g/dL, median [IQR]** | 2.9(2.4-3.4) | 3.0(2.4-3.4) | 3.0(2.4-3.5) | 2.8(2.4-3.4) | 2.9(2.4-3.4) | 0.174 | |
| **ALT, IU/L, median [IQR]** | 26.0(15.0-58.0) | 27.0(16.0-55.0) | 25.0(15.0-54.0) | 24.5(16.0-51.2) | 30.0(16.2-73.5) | 0.061 | |
| **AST, IU/L, median [IQR]** | 36.0(21.0-81.0) | 38.0(21.2-79.0) | 34.0(21.0-72.0) | 31.5(19.0-65.0) | 42.0(24.0-100.0) | 0.003 | RW vs RI, DW |
| **Bands, median [IQR]** | 6.0(2.0-12.0) | 5.0(2.0-7.0) | 8.0(3.0-13.2) | 7.0(2.0-8.0) | 6.0(2.0-18.0) | 0.256 | |
| **Bicarbonate, mEq/L, median [IQR]** | 22.0(19.0-26.0) | 23.0(19.0-26.0) | 22.0(19.0-25.0) | 22.0(17.0-25.0) | 22.0(18.0-26.0) | 0.336 | |
| **Bilirubin, mg/dL, median [IQR]** | 0.8(0.5-1.7) | 0.9(0.5-1.9) | 0.8(0.5-1.6) | 0.8(0.5-1.4) | 0.9(0.5-1.9) | 0.056 | |
| **BMI, kg/m2, median [IQR]** | 27.6(23.2-33.2) | 27.2(23.4-32.1) | 27.8(23.3-33.5) | 27.6(22.9-32.4) | 27.6(23.1-33.2) | 0.481 | |
| **BUN, mg/dL, median [IQR]** | 22.0(13.0-40.0) | 22.0(13.0-41.0) | 22.0(13.0-40.0) | 22.5(13.7-37.2) | 23.0(14.0-39.0) | 0.935 | |
| **Chloride, mEq/L, median [IQR]** | 104.0(100.0-109.0) | 104.0(100.0-108.0) | 105.0(100.0-109.0) | 105.0(100.5-109.0) | 104.0(100.0-109.0) | 0.218 | |
| **Creatinine, mg/dL, median [IQR]** | 1.2(0.8-2.1) | 1.1(0.7-2.1) | 1.25(0.8-2.1) | 1.2(0.8-2.0) | 1.2(0.8-1.9) | 0.350 | |
| **CRP, mg/dL, median [IQR]** | 14.3(6.2-28.7) | 14.3(6.1-24.9) | 15.1(5.1-36.1) | 13.6(8.0-22.9) | 12.7(6.6-26.8) | 0.974 | |
| **GCS, median [IQR]** | 13.0(8.0-15.0) | 15.0(10.0-15.0) | 10.0(7.0-15.0) | 11.0(7.0-15.0) | 14.0(10.0-15.0) | <0.001 | DI vs RI, DW; RI vs DW, RW; DW vs RW |
| **Glucose, mg/dL, median [IQR]** | 144.0(112.5-196.0) | 138.0(111.5-185.5) | 145.0(114.0-194.0) | 149.0(117.0-215.0) | 146.0(110.0-200.0) | 0.364 | |
| **Heart rate, beats/min, median [IQR]** | 96.0(82.0-111.0) | 96.0(80.0-113.0) | 96.0(82.2-110.0) | 97.5(78.0-111.7) | 100.0(85.0-115.0) | 0.179 | |
| **Hemoglobin, g/dL** | 10.0(8.3-11.9) | 9.9(8.3-11.8) | 10.2(8.5-12.0) | 9.7(8.0-11.8) | 9.6(8.0-11.9) | 0.109 | |
| **INR, median [IQR]** | 1.3(1.2-1.7) | 1.4(1.2-1.7) | 1.3(1.2-1.6) | 1.4(1.2-1.6) | 1.4(1.1-1.8) | 0.427 | |
| **Lactate, mmol/L, median [IQR]** | 15.6(9.4-37.1) | 14.6(9.1-31.5) | 19.1(9.6-34.9) | 20.7(8.5-51.2) | 19.3(11.1-62.1) | 0.810 | |
| **lymphocyte count, median [IQR]** | 900.0(500.0-1500.0) | 865.0(500.0-1600.0) | 1000.0(500.0-1500.0) | 800.0(400.0-1300.0) | 870.0(400.0-1600.0) | 0.177 | |
| **lymphocyte percentage, %, median [IQR]** | 10.0(5.0-18.0) | 9.0(5.0-20.0) | 10.0(5.0-17.0) | 8.0(5.0-15.0) | 10.0(5.0-18.0) | 0.602 | |
| **Pao2, mm Hg** | 99.0(73.0-146.0) | 95.0(71.1-148.5) | 100.0(74.0-142.0) | 93.5(71.8-136.5) | 103.0(73.0-152.7) | 0.697 | |
| **Platelet, x10(9)/L, median [IQR]** | 176.0(107.0-254.0) | 163.0(100.5-241.0) | 187.0(113.0-262.0) | 186.0(121.0-270.5) | 162.0(93.0-236.0) | <0.001** | DI vs RI DW; RW vs RI, DW |
| **RDW, %, median [IQR]** | 15.5(14.1-17.6) | 15.5(14.1-17.8) | 15.3(14.0-17.4) | 15.7(14.1-17.6) | 15.9(14.5-17.9) | <0.001 | RI vs RW |

| | | | | | | | |
|---|---|---|---|---|---|---|---|
| **Respiratory rate, breaths/min, median [IQR]** | 25.0(20.0-32.0) | 26.0(20.0-32.0) | 24.00(20.0-31.0) | 26.0(20.0-32.0) | 26.0(20.0-34.0) | <0.001§ | DI vs RI; RI vs RW |
| **SO2, %, median [IQR]** | 93.0(90.0-96.0) | 93.0(90.0-96.0) | 94.0(90.0-96.0) | 94.0(90.0-96.0) | 93.0(90.0-96.0) | 0.072 | |
| **Sodium, mmol/L, median [IQR]** | 137.0(134.0-140.0) | 137.0(133.0-140.0) | 137.0(134.0-141.0) | 137.0(133.5-140.0) | 137.0(134.0-140.0) | 0.170 | |
| **Systolic ABP, mm Hg, median [IQR]** | 93.0(79.0-110.0) | 98.0(82.0-117.0) | 91.0(78.0-108.0) | 92.0(77.0-106.0) | 95.0(81.0-113.0) | <0.001 | DI vs RI, DW, RW; RI vs RW; DW vs RW |
| **Temperature, degrees C, median [IQR]** | 37.1(36.7-37.9) | 37.1(36.7-37.9) | 37.1(36.7-37.8) | 37.1(36.6-37.8) | 37.1(36.6-37.9) | 0.822 | |
| **Troponin I, ng/mL, median [IQR]** | 0.04(0.02-0.14) | 0.04(0.02-0.16) | 0.04(0.02-0.11) | 0.05(0.02-0.15) | 0.04(0.02-0.15) | 0.381 | |
| **WBC, x10(9)/L, median [IQR]** | 12.0(8.0-17.3) | 11.8(7.8-16.6) | 12.4(8.3-17.3) | 11.4(8.0-18.1) | 11.5(6.9-17.1) | <0.001 | RI vs RW |

Definition of abbreviations: IQR--interquartile range; SD--standard deviation; ALT-- alanine aminotransferase; AST-- alanine aminotransferase; BMI-- body mass index, BUN-- blood urea nitrogen; CRP-- C-reactive protein; GCS-- Glasgow coma scale score; RDW-- red blood cell distribution width; WBC--white blood cell count; DI-- Delayed Improving; RI--Rapidly Improving; DW-- Delayed Worsening; RW-- Rapidly Worsening.

† p-value calculated by analysis of variance (ANOVA)/Kruskal–Wallis test

** False discovery rate corrected p-value < 0.001

§ Age adjusted (analysis of covariance [ANCOVA]) p-value < 0.001

#### Table S9. Patient basic characteristics comparisons among subphenotypes in the eICU validation cohort

| Characteristics | Total N=12,282 | DI N=2,005 | RI N=5,938 | DW N=1,392 | RW N=2,947 | p-value† |
|---|---|---|---|---|---|---|
| **Age, median (IQR)** | 67.0 [56.0-78.0] | 68.0 [55.0-79.0] | 67.0 [55.0-78.0] | 68.0 [57.0-79.0] | 68.0 [56.0-80.0] | 0.049 |
| **Sex, No. (%)** | | | | | | 0.979 |
| Male | 6438 (52.4%) | 1054 (52.5%) | 3116 (52.4%) | 729 (52.3%) | 1539 (52.2%) | |
| Female | 5844 (47.5%) | 951 (47.4%) | 2822 (47.5%) | 663 (47.6%) | 1408 (47.7%) | |
| **Ethnicity, No. (%)** | | | | | | 0.052 |
| Caucasian | 9434 (76.8%) | 1554 (77.5%) | 4584 (77.2%) | 1076 (77.3%) | 2220 (75.3%) | |
| African American | 1317 (10.7%) | 212 (10.5%) | 623 (10.4%) | 165 (11.8%) | 317 (10.7%) | |
| Hispanic | 593 (4.8%) | 87 (4.3%) | 264 (4.4%) | 63 (4.5%) | 179 (6.0%) | |
| Native American | 111 (0.9%) | 13 (0.6%) | 55 (0.9%) | 16 (1.1%) | 27 (0.9%) | |
| Asian | 201 (1.6%) | 32 (1.6%) | 104 (1.7%) | 14 (1.0%) | 51 (1.7%) | |
| Other/Unknown | 626 (5.1%) | 107 (5.3%) | 308 (5.1%) | 58 (4.1%) | 153 (5.1%) | |
| **Elixhauser index, median (IQR)** | 5.0 [0.0-12.0] | 5.0 [0.0-14.0] | 3.0 [0.0-11.0] | 7.0 [0.0-15.0] | 6.0 [0.0-14.0] | <0.001 |
| **Length stay, median (IQR)** | 2.8 [1.7-5.1] | 3.3 [2.1-5.7] | 2.4 [1.5-4.0] | 3.4 [1.9-6.0] | 3.0 [1.6-6.6] | <0.001 |
| **Baseline SOFA, mean (SD)** | 5.9 (3.08) | 5.4 (2.7) | 6.1 (3.1) | 6.3 (3.1) | 5.5 (2.9) | <0.001 |
| **Mechanical ventilation at admission, No. (%)** | 5329 (43.3%) | 877 (43.7%) | 2590 (43.6%) | 609 (43.7%) | 1253 (42.5%) | 0.751 |
| **ICU unit at admission, No. (%)** | | | | | | 0.381 |
| SICU | 627 (5.1%) | 104 (5.1%) | 295 (4.9%) | 90 (6.4%) | 138 (4.6%) | |
| MICU | 1575 (12.8%) | 261 (13.0%) | 770 (12.9%) | 168 (12.0%) | 376 (12.7%) | |
| CTICU | 87 (0.7%) | 15 (0.7%) | 33 (0.5%) | 14 (1.0%) | 25 (0.8%) | |
| CSICU | 160 (1.3%) | 27 (1.3%) | 72 (1.2%) | 15 (1.1%) | 46 (1.5%) | |
| MSICU | 7797 (63.4%) | 1267 (63.1%) | 3809 (64.1%) | 858 (61.6%) | 1863 (63.2%) | |
| CCU-CTICU | 772 (6.2%) | 131 (6.5%) | 373 (6.2%) | 85 (6.1%) | 183 (6.2%) | |
| CICU | 907 (7.3%) | 143 (7.1%) | 410 (6.9%) | 122 (8.7%) | 232 (7.8%) | |
| Neuro ICU | 357 (2.9%) | 57 (2.8%) | 176 (2.9%) | 40 (2.8%) | 84 (2.8%) | |
| **Admission location, No. (%)** | | | | | | <0.001 |
| ICU | 4 (0.03%) | 2 (0.1%) | 1 (0.02%) | 0 (0.0%) | 1 (0.03%) | |
| ICU to SDU | 1 (0.01%) | 0 (0.0%) | 0 (0.0%) | 0 (0.0%) | 1 (0.03%) | |
| Floor | 1986 (16.1%) | 360 (17.9%) | 824 (13.8%) | 225 (16.1%) | 577 (19.5%) | |
| Other Hospital | 404 (3.2%) | 76 (3.7%) | 180 (3.0%) | 61 (4.3%) | 87 (2.9%) | |
| Operating Room | 477 (3.8%) | 84 (4.1%) | 264 (4.4%) | 60 (4.3%) | 69 (2.3%) | |
| PACU | 17 (0.14%) | 2 (0.1%) | 11 (0.1%) | 2 (0.1%) | 2 (0.07%) | |
| Acute Care/Floor | 284 (2.3%) | 42 (2.0%) | 128 (2.1%) | 28 (2.0%) | 86 (2.9%) | |
| Direct Admit | 852 (6.9%) | 156 (7.7%) | 397 (6.6%) | 106 (7.6%) | 193 (6.5%) | |
| Other ICU | 63 (0.5%) | 13 (0.6%) | 34 (0.5%) | 5 (0.3%) | 11 (0.3%) | |
| SDU | 218 (1.7%) | 35 (1.7%) | 99 (1.6%) | 31 (2.2%) | 53 (1.8%) | |
| Recovery Room | 146 (1.1%) | 24 (1.2%) | 86 (1.4%) | 16 (1.1%) | 20 (0.6%) | |
| Emergency Department | 7825 (63.7%) | 1210 (60.3%) | 3911 (65.8%) | 858 (61.6%) | 1846 (62.6%) | |
| **Infection item, No. (%)** | | | | | | |
| Central nervous system | 63 (0.5%) | 17 (0.8%) | 20 (0.3%) | 8 (0.5%) | 18 (0.6%) | 0.033 |
| Intra-Abdominal | 790 (6.4%) | 139 (6.9%) | 341 (5.7%) | 112 (8.0%) | 198 (6.7%) | 0.007 |
| Pneumonia | 4743 (38.6%) | 834 (41.6%) | 2137 (35.9%) | 545 (39.1%) | 1227 (41.6%) | <0.001 |
| Septicemia bacteremia | 7350 (59.8%) | 1118 (55.7%) | 3582 (60.3%) | 827 (59.4%) | 1823 (61.8%) | <0.001 |
| Skin soft tissue | 632 (5.1%) | 112 (5.5%) | 304 (5.1%) | 68 (4.8%) | 148 (5.0%) | 0.779 |
| Urinary tract | 2478 (20.1%) | 388 (19.3%) | 1231 (20.7%) | 314 (22.5%) | 545 (18.4%) | 0.007 |
| **Septic shock, No. (%)** | 1649 (13.4%) | 250 (12.4%) | 719 (12.1%) | 166 (11.9%) | 514 (17.4%) | <0.001 |

Definition of abbreviations: IQR--interquartile range; SD--standard deviation; SOFA--Sequential Organ Failure Assessment; SICU-- Surgical ICU; MICU--Medical ICU; CTICU--Cardiothoracic ICU; CSICU--Cardiac Surgery ICU; MSICU--Medical-Surgical ICU; CCU--Coronary Care Unit; CICU-- Cardiac ICU; Neuro-ICU--Neurological ICU; SDU--Step-Down Unit; DI-- Delayed Improving; RI--Rapidly Improving; DW--Delayed Worsening; RW-- Rapidly Worsening.

†p-value calculated by Chi-square test/Fisher's exact test, or student's t-test/Mann-Whitney test where appropriate.

**Table S10. Patient physiological characteristics comparisons among subphenotypes in the eICU validation cohort**

| Characteristics | Total N=12,282 | DI N=2,005 | RI N=5,938 | DW N=1,392 | RW N=2,947 | p-value† | Post-hoc |
|---|---|---|---|---|---|---|---|
| Albumin, g/dL, median [IQR] | 2.7 [2.1-3.3] | 2.7 [2.0-3.3] | 2.8 [2.2-3.4] | 2.7 [2.0-3.3] | 2.6 [2.0-3.2] | <0.001 | RI vs RW |
| ALT, IU/L, median [IQR] | 23.0 [13.0-44.7] | 22.0 [11.0-42.0] | 23.0 [12.0-42.0] | 24.0 [13.0-47.0] | 25.0 [13.0-50.0] | <0.001 | RI vs DW, RW |
| AST, IU/L, median [IQR] | 29.0 [17.0-59.0] | 28.0 [16.0-54.0] | 28.0 [16.0-54.0] | 31.0 [17.0-66.0] | 34.0 [18.0-74.5] | <0.001 | RW vs DI, RI |
| Bands, median [IQR] | 11.0 [4.0-20.0] | 10.0 [5.0-19.0] | 10.0 [4.0-20.0] | 11.0 [5.0-19.0] | 11.0 [4.0-21.0] | 0.126 | |
| Bicarbonate, mEq/L, median [IQR] | 21.4 [17.4-25.0] | 22.0 [18.0-26.0] | 21.6 [18.0-25.0] | 21.8 [17.0-25.05] | 21.0 [16.0-24.9] | <0.001 | RI vs RW |
| Bilirubin, mg/dL, median [IQR] | 0.7 [0.4-1.2] | 0.7 [0.4-1.2] | 0.7 [0.4-1.2] | 0.7 [0.4-1.3] | 0.8 [0.5-1.4] | <0.001 | RW vs DI, RI, DW |
| BMI, kg/m2, median [IQR] | 27.1 [22.8-33.1] | 26.9 [22.6-33.3] | 27.5 [23.2-33.4] | 26.8 [22.6-32.4] | 26.6 [22.3-32.6] | <0.001 | RI vs RW |
| BUN, mg/dL, median [IQR] | 30.0 [19.0-49.0] | 28.0 [17.0-45.0] | 31.0 [19.0-50.0] | 32.0 [20.0-52.0] | 30.0 [19.0-48.0] | <0.001 | DI vs DW |
| Chloride, mEq/L, median [IQR] | 103.0 [99.0-108.0] | 103.0 [99.0-108.0] | 103.0 [99.0-108.0] | 104.0 [99.0-109.0] | 103.0 [99.0-108.0] | 0.035 | DI vs DW |
| Creatinine, mg/dL, median [IQR] | 1.5 [0.9-2.5] | 1.3 [0.8-2.3] | 1.5 [1.0-2.6] | 1.6 [1.0-2.5] | 1.5 [0.9-2.6] | <0.001 | DI vs RI, DW, RW |
| CRP, mg/dL, median [IQR] | 16.6 [6.2-31.6] | 13.0 [6.1-26.5] | 14.9 [5.9-29.1] | 15.9 [6.6-30.7] | 21.9 [9.6-46.5] | 0.160 | |
| GCS, median [IQR] | 14.0 [9.0-15.0] | 14.0 [10.0-15.0] | 13.0 [8.0-15.0] | 12.0 [8.0-15.0] | 14.0 [11.0-15.0] | <0.001 | DI vs DW |
| Glucose, mg/dL, median [IQR] | 141.0 [112.0-194.0] | 138.0 [109.0-188.0] | 143.0 [113.0-198.0] | 143.0 [112.7-198.0] | 138.0 [109.0-190.0] | <0.001 | DI vs DW |
| Heart rate, beats/min, median [IQR] | 106.0 [89.0-123.0] | 107.0 [90.0-124.0] | 103.0 [87.0-121.0] | 106.0 [89.0-122.0] | 109.0 [91.0-126.0] | <0.001 | RI vs DI, DW, RW |
| Hemoglobin, g/dL | 10.8 [9.1-12.6] | 10.8 [9.0-12.6] | 11.0 [9.3-12.6] | 10.6 [9.0-12.5] | 10.7 [9.0-12.6] | 0.075 | |
| INR, median [IQR] | 1.3 [1.1-1.7] | 1.3 [1.1-1.7] | 1.2 [1.1-1.6] | 1.3 [1.1-1.7] | 1.3 [1.1-1.8] | <0.001 | RI vs RW |
| Lactate, mmol/L, median [IQR] | 1.7 [0.6-3.2] | 1.5 [0.0-2.9] | 1.6 [0.5-2.9] | 1.75 [0.8-3.4] | 2.0 [0.9-4.0] | <0.001[§] | DI vs RW |
| lymphocyte percentage, %, median [IQR] | 6.8 [2.7-12.0] | 6.7 [3.0-12.3] | 7.0 [3.0-12.0] | 7.0 [3.0-12.1] | 6.0 [2.0-12.0] | <0.001 | RW vs DI, RI, DW |
| Pao2, mm Hg | 77.9 [61.0-108.0] | 77.0 [60.0-107.0] | 79.0 [61.4-108.0] | 78.9 [61.0-112.0] | 75.5 [60.4-105.0] | 0.194 | |
| Platelet, x10(9)/L, median [IQR] | 191.0 [129.0-266.0] | 187.0 [133.0-266.0] | 200.0 [136.0-278.0] | 184.0 [125.0-252.0] | 176.0 [117.0-248.0] | <0.001[**] | RI vs DI, DW, RW |
| RDW, %, median [IQR] | 15.3 [14.1-17.1] | 15.5 [14.3-17.4] | 15.2 [14.0-16.7] | 15.6 [14.2-17.4] | 15.6 [14.3-17.5] | <0.001 | RI vs DI, DW, RW |
| Respiratory rate, breaths/min, median [IQR] | 26.0 [21.0-33.0] | 27.0 [22.0-33.0] | 26.0 [20.0-32.0] | 26.0 [21.0-32.0] | 28.0 [22.0-35.0] | <0.001 | RI vs DW |
| SO2, %, median [IQR] | 93.0 [89.0-96.0] | 92.0 [88.0-96.0] | 93.0 [89.0-96.0] | 93.0 [88.0-96.0] | 92.0 [88.0-96.0] | <0.001 | RI vs DI, RW |
| Sodium, mmol/L, median [IQR] | 138.0 [135.0-141.0] | 138.0 [135.0-141.0] | 138.0 [135.0-141.0] | 138.0 [135.0-142.0] | 138.0 [134.0-141.0] | 0.170 | |
| Systolic ABP, mm Hg, median [IQR] | 89.0 [76.0-104.0] | 90.0 [77.0-106.0] | 90.0 [76.0-104.0] | 88.0 [76.0-102.0] | 88.0 [74.0-104.0] | 0.031 | DI vs DW |
| Temperature, degrees C, median [IQR] | 37.2 [36.8-38.1] | 37.22 [36.8-38.1] | 37.2 [36.8-38.1] | 37.2 [36.7-38.0] | 37.2 [36.7-38.0] | 0.072 | |
| Troponin I, ng/mL, median [IQR] | 0.1 [0.04-0.38] | 0.1 [0.03-0.4] | 0.1 [0.03-0.33] | 0.1 [0.04-0.58] | 0.1 [0.04-0.4] | 0.104 | |

| | | | | | | | |
|---|---|---|---|---|---|---|---|
| **WBC, x10(9)/L, median [IQR]** | 13.8 [9.0-19.5] | 13.4 [8.8-19.3] | 14.0 [9.5-19.4] | 13.3 [8.8-19.4] | 13.6 [8.3-19.9] | 0.003 | RI vs DW |

Definition of abbreviations: IQR--interquartile range; SD--standard deviation; ALT-- alanine aminotransferase; AST-- alanine aminotransferase; BMI-- body mass index, BUN-- blood urea nitrogen; CRP-- C-reactive protein; GCS-- Glasgow coma scale score; RDW-- red blood cell distribution width; WBC--white blood cell count; DI-- Delayed Improving; RI--Rapidly Improving; DW-- Delayed Worsening; RW-- Rapidly Worsening.

† p-value calculated by analysis of variance (ANOVA)/Kruskal–Wallis test

** False discovery rate corrected p-value < 0.001

§ Age adjusted (analysis of covariance [ANCOVA]) p-value < 0.001

**Table S11. Patient basic characteristics comparisons among subphenotypes in the CEDAR validation cohort**

| **Characteristics** | **Total N=4,804** | **DI N=382** | **RI N=2,561** | **DW N=564** | **RW N=1,297** | **p-value†** |
|---|---|---|---|---|---|---|
| **Age, median (IQR)** | 77.0 [66.0-88.0] | 77.0 [65.0-89.0] | 78.0 [65.0-88.0] | 79.0 [68.0-90.0] | 76.0 [65.0-87.0] | 0.010 |
| **Sex, No. (%)** | | | | | | 0.257 |
| Male | 2320 (48.2%) | 193 (50.5%) | 1204 (47.0%) | 274 (48.5%) | 649 (50.0%) | |
| Female | 2484 (51.7%) | 189 (49.4%) | 1357 (52.9%) | 290 (51.4%) | 648 (49.9%) | |
| **Race, No. (%)** | | | | | | 0.964 |
| WHITE | 1791 (37.2%) | 140 (36.6%) | 959 (37.4%) | 210 (37.2%) | 482 (37.1%) | |
| BLACK | 446 (9.2%) | 39 (10.2%) | 228 (8.9%) | 51 (9.0%) | 128 (9.8%) | |
| OTHER | 2567 (53.4%) | 203 (53.1%) | 1374 (53.6%) | 303 (53.7%) | 687 (52.9%) | |
| **Length stay, median (IQR)** | 4.4 [2.6-7.9] | 4.6 [2.8-7.8] | 3.9 [2.4-6.7] | 4.7 [2.8-7.9] | 5.7 [2.8-10.8] | <0.001 |
| **Mechanical ventilation at admission, No. (%)** | 2263(47.1%) | 151(39.5%) | 1174(45.8%) | 270(47.8%) | 668(51.5%) | 0.014 |
| **Elixhauser index, median (IQR)** | 12.5 [0.0-27.0] | 14.0 [0.0-29.0] | 11.0 [0.0-24.0] | 15.0 [0.0-26.0] | 16.0 [0.0-30.0] | <0.001 |
| **Baseline SOFA, mean (SD)** | 6.3 (3.1) | 5.0 (2.8) | 6.4 (3.0) | 6.7 (3.2) | 6.5 (3.2) | <0.001 |
| **ICU unit at admission, No. (%)** | | | | | | <0.001 |
| CTICU | 656 (13.6%) | 31 (8.1%) | 382 (14.9%) | 126 (22.3%) | 117 (9.0%) | |
| SICU | 752 (15.6%) | 60 (15.7%) | 492 (19.2%) | 56 (9.9%) | 144 (11.1%) | |
| MICU | 2641 (54.9%) | 213 (55.7) % | 1314 (51.3%) | 295 (52.3%) | 819 (63.1%) | |
| CICU | 755 (15.7%) | 78 (20.4%) | 373 (14.5%) | 87 (15.4%) | 217 (16.7%) | |
| **Infection item, No. (%)** | | | | | | |
| Central nervous system | 38 (0.7%) | 3 (0.7%) | 23 (0.9%) | 6 (1.0%) | 6 (0.4%) | 0.441 |
| Intra-Abdominal | 499 (10.3%) | 39 (10.2%) | 256 (10.0%) | 54 (9.5%) | 150 (11.5%) | 0.428 |
| Pneumonia | 597 (12.4%) | 52 (13.6%) | 291 (11.3%) | 63 (11.1%) | 191 (14.7%) | 0.016 |
| Septicemia bacteremia | 1376 (28.6%) | 101 (26.4%) | 684 (26.7%) | 155 (27.4%) | 436 (33.6%) | <0.001 |
| Skin soft tissue | 164 (3.4%) | 17 (4.4%) | 76 (2.9%) | 24 (4.2%) | 47 (3.6%) | 0.243 |
| Urinary tract | 1127 (23.4%) | 93 (24.3%) | 657 (25.6%) | 127 (22.5%) | 250 (19.2%) | <0.001 |
| **Septic shock, No. (%)** | 564 (11.7%) | 47 (12.3%) | 247 (9.6%) | 58 (10.2%) | 212 (16.3%) | <0.001 |

Definition of abbreviations: IQR--interquartile range; SD--standard deviation; SOFA--Sequential Organ Failure Assessment; CTICU--Cardiothoracic ICU; SICU-- Surgical ICU; MICU-- Medical ICU; CICU-- Cardiac ICU; DI-- Delayed Improving; RI--Rapidly Improving; DW--Delayed Worsening; RW-- Rapidly Worsening.

†p-value calculated by Chi-square test/Fisher's exact test, or student's t-test/Mann-Whitney test where appropriate.

**Table S12. Patient physiological characteristics comparisons among subphenotypes in the CEDAR validation cohort**

| Characteristics | Total N=4,804 | DI N=382 | RI N=2,561 | DW N=564 | RW N=1,297 | p-value† | Post-hoc |
|---|---|---|---|---|---|---|---|
| Albumin, g/dL, median [IQR] | 2.6 [2.1-3.2] | 2.7 [2.2-3.2] | 2.7 [2.3-3.2] | 2.6 [2.1-3.1] | 2.5 [1.9-3.0] | <0.001 | DI vs RW |
| ALT, IU/L, median [IQR] | 26.0 [17.0-53.0] | 26.0 [17.0-46.75] | 26.0 [17.0-49.75] | 24.0 [16.0-56.0] | 30.0 [18.0-64.0] | 0.113 | |
| AST, IU/L, median [IQR] | 38.0 [25.0-75.0] | 35.0 [24.0-62.0] | 36.0 [24.0-67.0] | 39.0 [26.0-93.0] | 42.0 [26.0-93.0] | 0.073 | |
| Bands, median [IQR] | 12.0 [5.0-21.0] | 13.5 [7.0-22.0] | 12.0 [5.0-21.0] | 12.0 [7.0-23.75] | 11.0 [6.0-21.0] | 0.340 | |
| Bicarbonate, mEq/L, median [IQR] | 20.4 [16.4-23.8] | 20.7 [16.7-23.9] | 21.0 [17.6-24.4] | 20.4 [16.7-23.4] | 18.7 [14.0-22.8] | <0.001 | RI vs RW |
| Bilirubin, mg/dL, median [IQR] | 1.1 [0.7-1.7] | 1.0 [0.7-1.8] | 1.1 [0.7-1.7] | 1.0 [0.7-1.6] | 1.1 [0.7-1.8] | 0.189 | |
| BMI, kg/m2, median [IQR] | 25.7 [22.0-30.4] | 25.0 [21.7-28.7] | 25.8 [22.2-30.6] | 26.3 [22.0-30.6] | 25.3 [21.7-30.1] | 0.053 | |
| BUN, mg/dL, median [IQR] | 27.0 [16.0-48.0] | 27.0 [16.0-48.2] | 25.0 [15.0-45.0] | 26.5 [16.0-46.7] | 31.0 [18.0-53.0] | <0.001 | RI vs DW |
| Chloride, mEq/L, median [IQR] | 106.0 [101.0-110.0] | 105.0 [100.0-109.0] | 106.0 [101.0-109.0] | 106.0 [102.0-110.0] | 105.0 [101.0-110.0] | 0.064 | |
| Creatinine, mg/dL, median [IQR] | 1.3 [0.9-2.3] | 1.3 [0.9-2.6] | 1.3 [0.8-2.1] | 1.3 [0.9-2.1] | 1.5 [0.9-2.4] | 0.042 | RW vs RI, DW |
| CRP, mg/dL, median [IQR] | 15.8 [7.8-26.8] | 19.1 [7.9-28.9] | 13.9 [7.1-26.6] | 18.7 [7.5-27.4] | 13.54 [8.25-26.56] | 0.926 | |
| GCS, median [IQR] | 12.0 [7.0-15.0] | 15.0 [10.0-15.0] | 10.0 [6.0-15.0] | 10.0 [3.0-15.0] | 14.0 [8.0-15.0] | <0.001 | RW vs RI, DW; DI vs RI, DW |
| Glucose, mg/dL, median [IQR] | 144.0 [113.0-189.0] | 139.0 [109.2-187.0] | 146.0 [114.0-189.0] | 146.0 [114.0-198.0] | 141.0 [111.0-187.0] | 0.032 | RW vs RI, DW |
| Heart rate, beats/min, median [IQR] | 110.0 [92.0-130.0] | 108.0 [93.0-129.0] | 106.0 [90.0-126.0] | 108.0 [91.0-128.0] | 119.0 [99.0-139.0] | <0.001 | RW vs DI, RI, DW |
| Hemoglobin, g/dL | 9.9 [8.3-11.6] | 9.8 [8.0-11.5] | 10.1 [8.6-11.7] | 9.7 [8.3-11.5] | 9.6 [8.0-11.4] | <0.001 | RI vs RW |
| INR, median [IQR] | 1.3 [1.1-1.6] | 1.2 [1.1-1.5] | 1.2 [1.1-1.5] | 1.3 [1.1-1.6] | 1.4 [1.2-1.7] | <0.001 | RW vs DI, RI |
| Lactate, mmol/L, median [IQR] | 2.5 [1.3-4.8] | 2.2 [1.1-4.4] | 2.1 [1.2-4.0] | 3.0 [1.6-5.4] | 3.2 [1.5-6.4] | <0.001[§] | RW vs DI, RI; DW vs DI, RI |
| lymphocyte percentage, %, median [IQR] | 8.4 [4.5-15.4] | 8.3 [4.5-13.5] | 8.7 [4.7-15.3] | 8.5 [4.7-15.5] | 8.0 [4.0-16.2] | 0.563 | |
| Pao2, mm Hg | 92.0 [70.0-129.0] | 89.0 [68.0-141.0] | 96.0 [73.0-132.0] | 92.0 [70.0-132.5] | 84.0 [67.0-118.2] | 0.283 | |
| Platelet, x10(9)/L, median [IQR] | 154.0 [93.0-229.0] | 164.0 [104.0-225.0] | 163.0 [101.7-242.0] | 149.0 [87.5-223.0] | 137.0 [66.5-204.0] | <0.001[**] | RW vs DI, RI, DW; RI vs DW |
| Respiratory rate, breaths/min, median [IQR] | 27.0 [22.0-33.0] | 28.0 [23.0-34.0] | 26.0 [22.0-31.0] | 26.0 [21.0-32.0] | 30.0 [24.0-36.0] | <0.001 | RW vs RI, DW |
| SO2, %, median [IQR] | 94.0 [90.0-96.0] | 94.0 [91.0-96.0] | 94.0 [91.0-96.0] | 94.0 [90.0-96.0] | 92.0 [87.0-95.0] | <0.001 | RW vs DI, RI, DW |
| Sodium, mmol/L, median [IQR] | 138.0 [135.0-141.0] | 138.0 [134.0-141.0] | 138.0 [135.0-141.0] | 139.0 [136.0-142.0] | 138.0 [134.0-141.0] | 0.172 | |
| Systolic ABP, mm Hg, median [IQR] | 88.0 [73.0-101.0] | 89.0 [74.0-103.0] | 91.0 [78.0-103.0] | 87.0 [70.0-99.0] | 79.0 [66.0-96.0] | <0.001 | RW vs DI, RI, DW |
| Temperature, degrees C, median [IQR] | 37.2 [36.8-38.1] | 37.2 [36.8-38.4] | 37.2 [36.8-38.0] | 37.1 [36.8-37.9] | 37.3 [36.8-38.4] | <0.001 | DW vs RW |
| Troponin I, ng/mL, median [IQR] | 0.1 [0.04-0.4] | 0.1 [0.03-0.8] | 0.08 [0.04-0.4] | 0.1 [0.05-0.4] | 0.1 [0.04-0.4] | 0.050 | |

| | | | | | | |
|---|---|---|---|---|---|---|
| **WBC, x10(9)/L, median [IQR]** | 11.5 [7.1-17.2] | 11.0 [7.0-16.0] | 12.0 [8.0-17.3] | 12.1 [7.7-17.8] | 10.1 [4.7-16.7] | <0.001 | DW vs RW |

Definition of abbreviations: IQR--interquartile range; SD--standard deviation; ALT-- alanine aminotransferase; AST-- alanine aminotransferase; BMI-- body mass index, BUN-- blood urea nitrogen; CRP-- C-reactive protein; GCS-- Glasgow coma scale score; WBC--white blood cell count; DI-- Delayed Improving; RI--Rapidly Improving; DW-- Delayed Worsening; RW-- Rapidly Worsening.

† p-value calculated by analysis of variance (ANOVA)/Kruskal–Wallis test

** False discovery rate corrected p-value < 0.001

§ Age adjusted (analysis of covariance [ANCOVA]) p-value < 0.001

**Table S13. Patient physiological characteristics comparisons among subphenotypes in terms of sensitivity analysis (GBTM) in the development cohort**

| Characteristics | Total (N=4,678) | DI (N=817) | RI (N=2,402) | DW (N=735) | RW (N=724) | p-value† | Post-hoc |
|---|---|---|---|---|---|---|---|
| **Albumin, g/dL, median [IQR]** | 2.9 [2.5-3.3] | 2.9 [2.5-3.4] | 2.9 [2.5-3.3] | 3.0 [2.5-3.3] | 2.9 [2.5-3.3] | 0.581 | |
| **ALT, IU/L, median [IQR]** | 35.0 [19.0-86.0] | 33.0 [20.0-79.5] | 36.0 [19.0-89.0] | 32.0 [18.0-78.7] | 38.5 [20.0-82.2] | 0.36 | |
| **AST, IU/L, median [IQR]** | 49.0 [27.0-120.0] | 47.0 [29.0-128.0] | 47.0 [26.0-111.0] | 47.0 [24.0-127.0] | 58.0 [30.5-142.0] | 0.002 | DI, RI, DW vs RW |
| **Bands, median [IQR]** | 2.0 [0.0-8.0] | 2.0 [0.0-8.2] | 2.0 [0.0-9.0] | 3.0 [0.0-8.0] | 2.0 [0.0-8.0] | 0.838 | |
| **Bicarbonate, mEq/L, median [IQR]** | 23.0 [20.0-26.0] | 24.0 [21.0-26.0] | 23.0 [20.0-25.0] | 23.0 [19.0-25.0] | 22.0 [19.0-26.0] | <0.001 | DI vs RI, DW, RW |
| **Bilirubin, mg/dL, median [IQR]** | 0.8 [0.4-2.1] | 1.0 [0.5-2.3] | 0.8 [0.4-2.0] | 0.7 [0.4-1.7] | 1.0 [0.5-2.6] | <0.001 | DI vs RI, DW; RW vs RI, DW |
| **BMI, kg/m2, median [IQR]** | 27.5 [23.7-32.2] | 27.1 [23.5-32.0] | 27.7 [23.8-32.6] | 27.8 [24.2-32.2] | 26.8 [23.4-31.4] | 0.318 | |
| **BUN, mg/dL, median [IQR]** | 22.0 [15.0-38.0] | 19.0 [13.0-30.0] | 22.0 [15.0-37.0] | 28.0 [16.0-49.0] | 24.0 [15.0-41.0] | <0.001 | DI vs RI, DW, RW; RI vs DW; DW vs RW |
| **Chloride, mEq/L, median [IQR]** | 106.0 [102.0-110.0] | 106.0 [101.0-109.0] | 106.0 [102.0-110.0] | 106.0 [102.0-110.0] | 106.0 [101.0-110.0] | 0.027 | DI vs RI; |
| **Creatinine, mg/dL, median [IQR]** | 1.1 [0.8-1.7] | 0.9 [0.7-1.3] | 1.1 [0.8-1.7] | 1.3 [0.9-2.5] | 1.1 [0.8-1.8] | <0.001 | DI vs RI, DW, RW; RI vs DW; DW vs RW |
| **CRP, mg/dL, median [IQR]** | 92.5 [40.5-174.4] | 125.4 [57.3-186.6] | 80.9 [29.2-171.3] | 101.0 [49.7-165.0] | 83.0 [42.6-164.8] | 0.524 | |
| **GCS, median [IQR]** | 13.0 [7.0-15.0] | 14.0 [7.0-15.0] | 13.0 [7.0-15.0] | 13.0 [7.0-15.0] | 13.0 [6.0-15.0] | 0.051 | |
| **Glucose, mg/dL, median [IQR]** | 131.0 [106.0-172.0] | 129.0 [105.0-167.0] | 132.0 [107.0-173.0] | 131.0 [105.0-172.0] | 135.0 [106.0-175.0] | 0.321 | |
| **Heart rate, beats/min, median [IQR]** | 95.0 [81.0-110.0] | 96.0 [82.0-110.0] | 94.0 [80.6-109.0] | 95.0 [80.63-110.0] | 96.0 [83.8-112.0] | 0.043 | RI, DW vs RW |
| **Hemoglobin, g/dL** | 10.6 [9.2-12.0] | 10.6 [9.3-11.9] | 10.6 [9.3-12.0] | 10.4 [9.1-11.9] | 10.5 [9.1-12.01] | 0.559 | |
| **INR, median [IQR]** | 1.3 [1.2-1.6] | 1.3 [1.1-1.6] | 1.3 [1.1-1.5] | 1.3 [1.2-1.7] | 1.4 [1.2-1.7] | <0.001** | DI vs DW, RW; RI vs DW, RW |
| **Lactate, mmol/L, median [IQR]** | 1.8 [1.2-2.8] | 1.7 [1.2-2.8] | 1.7 [1.1-2.7] | 1.8 [1.2-3.0] | 2.1 [1.3-3.2] | <0.001§ | DI, RI, DW vs RW |
| **lymphocyte count, median [IQR]** | 940.0 [675.0-1570.0] | 1161.0 [1020.0-2016.0] | 840.0 [584.5-1144.0] | 1095.0 [431.2-1823.2] | 782.0 [537.5-1502.5] | 0.422 | |
| **lymphocyte percentage, %, median [IQR]** | 7.1 [3.5-12.5] | 7.6 [4.0-12.4] | 7.1 [3.5-12.6] | 7.0 [3.5-12.2] | 6.7 [3.4-13.1] | 0.608 | |
| **Pao2, mm Hg** | 108.0 [81.5-168.5] | 101.0 [81.5-128.5] | 106.0 [71.0-146.5] | 138.0 [91.2-181.5] | 135.0 [89.5-202.7] | 0.189 | |
| **Platelet, x10(9)/L, median [IQR]** | 190.0 [130.0-260.0] | 184.0 [126.0-259.0] | 199.0 [137.0-265.0] | 189.5 [131.0-257.2] | 168.0 [115.0-238.0] | <0.001 | DI vs RI, RW; RI, DW vs RW |
| **RDW, %, median [IQR]** | 14.8 [13.8-16.3] | 14.8 [13.8-16.2] | 14.7 [13.7-16.1] | 14.9 [13.8-16.7] | 14.9 [13.8-16.8] | <0.001 | RI vs DW, RW |
| **Respiratory rate, breaths/min, median [IQR]** | 22.0 [19.0-27.0] | 22.6 [19.0-27.0] | 22.0 [19.0-26.0] | 22.0 [19.0-26.1] | 23.0 [19.0-27.5] | 0.085 | |
| **SO2, %, median [IQR]** | 94.0 [80.0-97.0] | 94.6 [83.0-97.0] | 93.0 [78.0-97.0] | 95.0 [82.0-97.2] | 94.0 [81.0-97.0] | 0.064 | |

| | | | | | | | |
|---|---|---|---|---|---|---|---|
| **Sodium, mmol/L, median [IQR]** | 139.0 [136.0-142.0] | 139.0 [136.0-141.0] | 139.0 [136.0-141.0] | 139.0 [136.0-142.0] | 139.0 [136.0-142.0] | 0.085 | |
| **Systolic ABP, mm Hg, median [IQR]** | 101.0 [91.0-115.0] | 102.5 [92.0-118.0] | 100.0 [91.0-113.0] | 102.0 [91.5-115.0] | 102.0 [91.3-117.0] | <0.001 | DI vs RI; RI vs DW, RW; |
| **Temperature, degrees C, median [IQR]** | 36.8 [36.3-37.4] | 36.8 [36.3-37.3] | 36.8 [36.3-37.5] | 36.8 [36.2-37.4] | 36.7 [36.2-37.3] | 0.011 | RI vs DW, RW |
| **Troponin T, ng/mL, median [IQR]** | 0.1 [0.03-0.3] | 0.1 [0.03-0.2] | 0.1 [0.03-0.3] | 0.1 [0.04-0.3] | 0.1 [0.03-0.3] | 0.129 | |
| **WBC, x10(9)/L, median [IQR]** | 11.1 [7.5-15.8] | 10.4 [7.0-14.5] | 11.4 [7.8-16.3] | 11.0 [7.4-15.0] | 10.9 [7.4-15.8] | <0.001 | DI vs RI, RI vs DW |

Definition of abbreviations: IQR--interquartile range; SD--standard deviation; ALT-- alanine aminotransferase; AST-- alanine aminotransferase; BMI-- body mass index, BUN-- blood urea nitrogen; CRP-- C-reactive protein; GCS-- Glasgow coma scale score; INR-- international normalized ratio; RDW-- red blood cell distribution width; ABP-- Ambulatory Blood Pressure; WBC--white blood cell count; DI-- Delayed Improving; RI-- Rapidly Improving; DW-- Delayed Worsening; RW-- Rapidly Worsening.

† p-value calculated by analysis of variance (ANOVA)/Kruskal–Wallis test

** False discovery rate corrected p-value < 0.001

§ Age adjusted (analysis of covariance [ANCOVA]) p-value < 0.001

**Table S14. Missing data (no., %) in cohorts for subphenotyping and predicting variables at the first six hours**

| Variable | Development Cohort (MIMIC-III) | Validation Cohort (NMEDW) | Validation Cohort (eICU) | Validation Cohort (CEDAR) |
|---|---|---|---|---|
| No. of patients | 4,678 | 3,665 | 12,282 | 4,804 |
| Age | 349 (7.4%) | 0 (0.0%) | 0 (0.0%) | 0 (0.0%) |
| Albumin | 2239 (47.8%) | 917 (25.0%) | 3055 (24.8%) | 1103 (22.9%) |
| ALT | 1746 (37.3%) | 984 (26.8%) | 3040 (24.7%) | 1164 (24.2%) |
| AST | 1747 (37.3%) | 976 (26.6%) | 2972 (24.1%) | 1162 (24.1%) |
| Bands | 3425 (73.2%) | 3083 (84.1%) | 9550 (81.0%) | 3779 (78.6%) |
| Bicarbonate | 24 (0.5%) | 295 (8.0%) | 533 (4.3%) | 261 (5.4%) |
| Bilirubin | 1729 (36.9%) | 593 (16.1%) | 3232 (26.3%) | 1120 (23.3%) |
| PaO2 | 1532 (32.7%) | 475 (12.9%) | 475 (33.1%) | 1398 (29.1%) |
| BMI | 2088 (44.6%) | 698 (19.0%) | 115 (0.9%) | 1786 (37.1%) |
| BUN | 26 (0.5%) | 12 (0.3%) | 603 (4.9%) | 247 (5.1%) |
| Chloride | 23 (0.4%) | 16 (0.4%) | 601 (4.8%) | 243 (5.0%) |
| Creatinine | 24 (0.5%) | 10 (0.2%) | 605 (4.9%) | 243 (5.0%) |
| CRP | 4522 (96.6%) | 3398 (92.7%) | 11737 (95.5%) | 4566 (95.0%) |
| GCS | 2 (0.04%) | 121 (3.3%) | 2949 (24.0%) | 238 (4.9%) |
| Gender | 0 (0.0%) | 0 (0.0%) | 0 (0.0%) | 0 (0.0%) |
| Glucose | 22 (0.4%) | 14 (0.3%) | 165 (1.3%) | 261 (5.4%) |
| Heart rate | 0 (0.0%) | 2 (0.05%) | 626 (5.0%) | 87 (1.8%) |
| Hemoglobin | 31 (0.6%) | 17 (0.4%) | 357 (2.9%) | 296 (2.1%) |
| INR | 511 (10.9%) | 977 (26.6%) | 3555 (28.9%) | 716 (14.9%) |
| Lactate | 1393 (29.7%) | 723 (19.7%) | 3240 (26.3%) | 1322 (27.5%) |
| Lymphocyte percent | 2012 (43.0%) | 2206 (60.1%) | 3025 (24.6%) | 1971 (41.0%) |
| Platelet | 32 (0.6%) | 10 (0.2%) | 439 (3.5%) | 35 (0.7%) |
| Race | 0 (0.0%) | 0 (0.0%) | 0 (0.0%) | 0 (0.0%) |
| RDW | 34 (0.7%) | 25 (0.6%) | 808(6.5%) | ----- |
| Respiratory rate | 0 (0.0%) | 120 (3.2%) | 866 (7.0%) | 88 (1.8%) |
| SO2 | 8 (0.17%) | 1 (0.03%) | 269 (2.1%) | 87 (1.8%) |
| Sodium | 23 (0.4%) | 14 (0.3%) | 269 (2.1%) | 18 (0.3%) |
| Systolic ABP | 1 (0.02%) | 7 (0.19%) | 748 (6.0%) | 90 (1.8%) |
| Temperature | 12 (0.2%) | 2 (0.05%) | 39 (3.0%) | 90 (1.8%) |
| Troponin | 3147 (67.2%) | 1922 (52.4%) | 7758 (63.1%) | 2798 (58.2%) |
| WBC | 32 (0.6%) | 22 (0.6%) | 407 (3.3%) | 60 (1.2%) |

Definition of abbreviations: ALT-- alanine aminotransferase; AST-- alanine aminotransferase; BMI-- body mass index, BUN-- blood urea nitrogen; CRP-- C-reactive protein; GCS-- Glasgow coma scale score; INR-- international normalized ratio; RDW-- red blood cell distribution width; ABP: Ambulatory Blood Pressure; WBC--white blood cell count.

**Table S15. International classification of diseases, ninth revision, clinical modification codes used to identify infectious syndromes.**

| **Infectious Syndrome** | **ICD-9-CM Codes** |
|---|---|
| Pneumonia | 480.0-480.9, 481, 482.0-482.9, 483.0-483.8, 484.1-484.8, 485, 486 |
| Urinary Tract Infections | 590.00, 590.01, 590.10, 590.11, 590.2, 590.3, 590.80, 590.81, 590.9, 595.0, 595.2, 595.3, 595.4, 595.89, 595.9, 597.0, 597.80, 597.89, 598.00, 598.01, 599.0 |
| Intra-abdominal Infections | 008.45, 009.0-009.3, 540.0-540.9, 541, 542, 543.9, 562.01, 562.03, 562.11, 562.13, 567.0-567.9, 569.5, 569.61, 569.71, 569.83, 572.0-572.8, 574.00-574.91, 575.0-575.9, 576.0-576.9, 614.0-614.9 |
| Skin/Soft Tissue Infections | 680-686, 035, 376.01, 728.86 |
| Central Nervous System | 006.5, 013.00-013.06, 013.10-013.16, 013.20-013.26, 013.30-013.36, 013.40-013.46, 013.50-013.56, 013.60-013.66, 013.80-013.86, 013.90-013.96, 036.0, 036.1, 049.8, 052.0, 052.2, 053.0, 053.14, 054.3, 054.72, 055.0, 056.01, 064, 072.1, 072.2, 091.81, 094.2, 098.82, 100.81, 112.83, 114.2, 130.0, 320.0-320.3, 320.7, 320.81, 323.01, 323.02, 323.1, 323.2, 323.41, 323.42, 324.0, 324.1, 324.9 |
| Septicemia/Bacteremia | 038.0-038.9, 790.7 |

**Table S16. The associations between comorbidities and subphenotypes in the MIMIC-III development cohort**

| Comorbidity Name | Total (N=4,678) | DI (N=1,174) | RI (N=1,932) | DW (N=960) | RW (N=612) | p-value |
|---|---|---|---|---|---|---|
| Congestive heart failure, No. (%) | 1327 (28.3%) | 372 (31.6%) | 502 (25.9%) | 302 (31.4%) | 151 (24.6%) | <0.001 |
| Valvular disease, No. (%) | 544 (11.6%) | 173 (14.7%) | 199 (10.3%) | 107 (11.1%) | 65 (10.6%) | 0.002 |
| Pulmonary circulation disorder, No. (%) | 514 (10.9%) | 161 (13.7%) | 177 (9.1%) | 109 (11.3%) | 67 (10.9%) | 0.001 |
| Peripheral vascular disorder, No. (%) | 469 (10.0%) | 123 (10.4%) | 177 (9.1%) | 117 (12.1%) | 52 (8.5%) | 0.038 |
| Hypertension, No. (%) | 2894 (61.8%) | 734 (62.5%) | 1165 (60.3%) | 619 (64.4%) | 376 (61.4%) | 0.168 |
| Paralysis, No. (%) | 173 (3.7%) | 43 (3.6%) | 68 (3.5%) | 36 (3.7%) | 26 (4.2%) | 0.872 |
| Other neurological, No. (%) | 903 (19.3%) | 232 (19.7%) | 372 (19.2%) | 191 (19.9%) | 108 (17.6%) | 0.693 |
| Chronic pulmonary disease, No. (%) | 1263 (27.0%) | 345 (29.3%) | 496 (25.6%) | 261 (27.1%) | 161 (26.3%) | 0.152 |
| Diabetes w/o chronic complications, No. (%) | 1147 (24.5%) | 300 (25.5%) | 473 (24.4%) | 248 (25.8%) | 126 (20.5%) | 0.083 |
| Diabetes w/ chronic complications, No. (%) | 380 (8.1%) | 80 (6.8%) | 149 (7.7%) | 104 (10.8%) | 47 (7.6%) | 0.005 |
| Hypothyroidism, No. (%) | 659 (14.0%) | 171 (14.5%) | 273 (14.1%) | 137 (14.2%) | 78 (12.7%) | 0.762 |
| Renal failure, No. (%) | 1075 (22.9%) | 212 (18.0%) | 408 (21.1%) | 327 (34.0%) | 128 (20.9%) | <0.001 |
| Liver disease, No. (%) | 852 (18.2%) | 233 (19.8%) | 289 (14.9%) | 163 (16.9%) | 167 (27.2%) | <0.001 |
| Chronic Peptic ulcer disease, No. (%) | 93 (1.9%) | 28 (2.3%) | 37 (1.9%) | 16 (1.6%) | 12 (1.9%) | 0.679 |
| HIV_AIDS, No. (%) | 41 (0.8%) | 8 (0.6%) | 20 (1.0%) | 10 (1.0%) | 3 (0.4%) | 0.489 |
| Lymphoma, No. (%) | 118 (2.5%) | 31 (2.6%) | 44 (2.2%) | 28 (2.9%) | 15 (2.4%) | 0.763 |
| Metastatic cancer, No. (%) | 359 (7.6%) | 86 (7.3%) | 151 (7.8%) | 75 (7.8%) | 47 (7.6%) | 0.963 |
| Solid tumor without metastasis, No. (%) | 536 (11.4%) | 132 (11.2%) | 226 (11.7%) | 105 (10.9%) | 73 (11.9%) | 0.907 |
| Rheumatoid arthritis/collagen vascular diseases, No. (%) | 244 (5.2%) | 55 (4.6%) | 99 (5.1%) | 56 (5.8%) | 34 (5.5%) | 0.663 |
| Coagulation deficiency, No. (%) | 1007 (21.5%) | 249 (21.2%) | 366 (18.9%) | 205 (21.3%) | 187 (30.5%) | <0.001 |
| Obesity, No. (%) | 383 (8.1%) | 107 (9.1%) | 151 (7.8%) | 78 (8.1%) | 47 (7.6%) | 0.591 |
| Weight loss, No. (%) | 408 (8.7%) | 111 (9.4%) | 165 (8.5%) | 78 (8.1%) | 54 (8.8%) | 0.727 |
| Fluid and electrolyte disorders, No. (%) | 2537 (54.2%) | 615 (52.3%) | 1031 (53.3%) | 547 (56.9%) | 344 (56.2%) | 0.108 |
| Blood loss anemia, No. (%) | 103 (2.2%) | 40 (3.4%) | 28 (1.4%) | 23 (2.4%) | 12 (1.9%) | 0.004 |
| Deficiency anemias, No. (%) | 214 (4.5%) | 63 (5.3%) | 83 (4.3%) | 42 (4.3%) | 26 (4.2%) | 0.519 |
| Alcohol abuse, No. (%) | 562 (12.0%) | 157 (13.3%) | 217 (11.2%) | 96 (10.0%) | 92 (15.0%) | 0.007 |
| Drug abuse, No. (%) | 285 (6.0%) | 62 (5.2%) | 130 (6.7%) | 56 (5.8%) | 37 (6.0%) | 0.418 |
| Psychoses, No. (%) | 112 (2.3%) | 26 (2.2%) | 51 (2.6%) | 28 (2.9%) | 7 (1.1%) | 0.118 |
| Depression, No. (%) | 788 (16.8%) | 189 (16.1%) | 336 (17.3%) | 171 (17.8%) | 92 (15.0%) | 0.399 |

**Table S17. The associations between comorbidities and subphenotypes in the NMEDW validation cohort**

| Comorbidity Name | Total N=3,665 | DI N=668 | RI N=2,019 | DW N=302 | RW N=676 | p-value |
|---|---|---|---|---|---|---|
| Congestive heart failure, No. (%) | 664 (18.1%) | 127 (19.0%) | 338 (16.7%) | 67 (22.1%) | 132 (19.5%) | 0.065 |
| Valvular disease, No. (%) | 257 (7.0%) | 49 (7.3%) | 129 (6.3%) | 23 (7.6%) | 56 (8.2%) | 0.368 |
| Pulmonary circulation disorder, No. (%) | 266 (7.2%) | 46 (6.8%) | 138 (6.8%) | 16 (5.3%) | 66 (9.7%) | 0.034 |
| Peripheral vascular disorder, No. (%) | 251 (6.8%) | 45 (6.7%) | 125 (6.1%) | 20 (6.6%) | 61 (9.0%) | 0.093 |
| Hypertension, No. (%) | 1005 (27.4%) | 202 (30.2%) | 543 (26.8%) | 95 (31.4%) | 165 (24.4%) | 0.037 |
| Paralysis, No. (%) | 91 (2.4%) | 12 (1.8%) | 45 (2.2%) | 11 (3.6%) | 23 (3.4%) | 0.118 |
| Other neurological, No. (%) | 841 (22.9%) | 154 (23.0%) | 418 (20.7%) | 84 (27.8%) | 185 (27.3%) | 0.001 |
| Chronic pulmonary disease, No. (%) | 549 (14.9%) | 98 (14.6%) | 299 (14.8%) | 52 (17.2%) | 100 (14.7%) | 0.728 |
| Diabetes w/o chronic complications, No. (%) | 423 (11.5%) | 78 (11.6%) | 224 (11.0%) | 41 (13.5%) | 80 (11.8%) | 0.640 |
| Diabetes w/ chronic complications, No. (%) | 255 (6.9%) | 49 (7.3%) | 132 (6.5%) | 28 (9.2%) | 46 (6.8%) | 0.359 |
| Hypothyroidism, No. (%) | 198 (5.4%) | 43 (6.4%) | 101 (5.0%) | 14 (4.6%) | 40 (5.9%) | 0.435 |
| Renal failure, No. (%) | 519 (14.1%) | 105 (15.7%) | 237 (11.7%) | 58 (19.2%) | 119 (17.6%) | <0.001 |
| Liver disease, No. (%) | 371 (10.1%) | 73 (10.9%) | 165 (8.1%) | 31 (10.2%) | 102 (15.0%) | <0.001 |
| Chronic Peptic ulcer disease, No. (%) | 49 (1.3%) | 11 (1.6%) | 21 (1.0%) | 5 (1.6%) | 12 (1.7%) | 0.384 |
| HIV_AIDS, No. (%) | 27 (0.7%) | 3 (0.4%) | 17 (0.8%) | 3 (0.9%) | 4 (0.5%) | 0.676 |
| Lymphoma, No. (%) | 94 (2.5%) | 19 (2.8%) | 41 (2.0%) | 7 (2.3%) | 27 (3.9%) | 0.044 |
| Metastatic cancer, No. (%) | 173 (4.7%) | 40 (5.9%) | 80 (3.9%) | 15 (4.9%) | 38 (5.6%) | 0.101 |
| Solid tumor without metastasis, No. (%) | 305 (8.3%) | 67 (10.0%) | 153 (7.5%) | 18 (5.9%) | 67 (9.9%) | 0.037 |
| Rheumatoid arthritis/collagen vascular diseases, No. (%) | 90 (2.4%) | 23 (3.4%) | 39 (1.9%) | 9 (2.9%) | 19 (2.8%) | 0.125 |
| Coagulation deficiency, No. (%) | 470 (12.8%) | 97 (14.5%) | 206 (10.2%) | 47 (15.5%) | 120 (17.7%) | <0.001 |
| Obesity, No. (%) | 167 (4.5%) | 29 (4.3%) | 94 (4.6%) | 14 (4.6%) | 30 (4.4%) | 0.986 |
| Weight loss, No. (%) | 434 (11.8%) | 92 (13.7%) | 193 (9.5%) | 41 (13.5%) | 108 (15.9%) | <0.001 |
| Fluid and electrolyte disorders, No. (%) | 1397 (38.1%) | 273 (40.8%) | 676 (33.4%) | 137 (45.3%) | 311 (46.0%) | <0.001 |
| Blood loss anemia, No. (%) | 108 (2.9%) | 22 (3.2%) | 45 (2.2%) | 12 (3.9%) | 29 (4.2%) | 0.026 |
| Deficiency anemias, No. (%) | 126 (3.4%) | 23 (3.4%) | 60 (2.9%) | 15 (4.9%) | 28 (4.1%) | 0.216 |
| Alcohol abuse, No. (%) | 221 (6.0%) | 45 (6.7%) | 111 (5.5%) | 20 (6.6%) | 45 (6.6%) | 0.521 |
| Drug abuse, No. (%) | 96 (2.6%) | 17 (2.5%) | 55 (2.7%) | 7 (2.3%) | 17 (2.5%) | 0.971 |
| Psychoses, No. (%) | 66 (1.8%) | 10 (1.5%) | 43 (2.1%) | 7 (2.3%) | 6 (0.8%) | 0.156 |
| Depression, No. (%) | 210 (5.7%) | 40 (5.9%) | 114 (5.6%) | 23 (7.6%) | 33 (4.8%) | 0.392 |

**Table S18. The associations between comorbidities and subphenotypes in the eICU validation cohort**

| Comorbidity Name | Total N=12,282 | DI N=2,005 | RI N=5,938 | DW N=1,392 | RW N=2,947 | p-value |
|---|---|---|---|---|---|---|
| Congestive heart failure, No. (%) | 1518 (12.3%) | 288 (14.3%) | 628 (10.5%) | 205 (14.7%) | 397 (13.4%) | <0.001 |
| Valvular disease, No. (%) | 119 (0.9%) | 33 (1.6%) | 35 (0.5%) | 20 (1.4%) | 31 (1.1%) | <0.001 |
| Pulmonary circulation disorder, No. (%) | 181 (1.4%) | 35 (1.7%) | 78 (1.3%) | 19 (1.3%) | 49 (1.6%) | 0.406 |
| Peripheral vascular disorder, No. (%) | 86 (0.7%) | 16 (0.8%) | 26 (0.4%) | 11 (0.7%) | 33 (1.1%) | 0.003 |
| Hypertension, No. (%) | 1456 (11.8%) | 270 (13.4%) | 633 (10.6%) | 193 (13.8%) | 360 (12.2%) | <0.001 |
| Paralysis, No. (%) | 74 (0.6%) | 17 (0.8%) | 40 (0.6%) | 6 (0.4%) | 11 (0.3%) | 0.122 |
| Other neurological, No. (%) | 1427 (11.6%) | 261 (13.0%) | 665 (11.2%) | 184 (13.2%) | 317 (10.7%) | 0.015 |
| Chronic pulmonary disease, No. (%) | 1601 (13.0%) | 311 (15.5%) | 791 (13.3%) | 189 (13.5%) | 310 (10.5%) | <0.001 |
| Diabetes w/o chronic complications, No. (%) | 444 (3.6%) | 65 (3.2%) | 242 (4.0%) | 46 (3.3%) | 91 (3.0%) | 0.067 |
| Hypothyroidism, No. (%) | 508 (4.1%) | 89 (4.4%) | 234 (3.9%) | 66 (4.7%) | 119 (4.0%) | 0.495 |
| Renal failure, No. (%) | 1755 (14.2%) | 297 (14.8%) | 755 (12.7%) | 247 (17.7%) | 456 (15.4%) | <0.001 |
| Liver disease, No. (%) | 670 (5.4%) | 103 (5.1%) | 239 (4.0%) | 85 (6.1%) | 243 (8.2%) | <0.001 |
| Chronic Peptic ulcer disease, No. (%) | 76 (0.6%) | 14 (0.7%) | 30 (0.5%) | 17 (1.2%) | 15 (0.5%) | 0.017 |
| Lymphoma, No. (%) | 127 (1.0%) | 28 (1.4%) | 41 (0.6%) | 19 (1.3%) | 39 (1.3%) | 0.004 |
| Metastatic cancer, No. (%) | 174 (1.4%) | 31 (1.5%) | 68 (1.1%) | 12 (0.8%) | 63 (2.1%) | 0.001 |
| Solid tumor without metastasis, No. (%) | 500 (4.0%) | 91 (4.5%) | 199 (3.3%) | 52 (3.7%) | 158 (5.3%) | <0.001 |
| Rheumatoid arthritis/collagen vascular diseases, No. (%) | 97 (0.7%) | 15 (0.7%) | 40 (0.6%) | 12 (0.8%) | 30 (1.0%) | 0.374 |
| Coagulation deficiency, No. (%) | 1108 (9.0%) | 200 (9.9%) | 396 (6.6%) | 159 (11.4%) | 353 (11.9%) | <0.001 |
| Obesity, No. (%) | 211 (1.7%) | 60 (2.9%) | 85 (1.4%) | 14 (1.0%) | 52 (1.7%) | <0.001 |
| Weight loss, No. (%) | 777 (6.3%) | 164 (8.1%) | 313 (5.2%) | 118 (8.4%) | 182 (6.1%) | <0.001 |
| Fluid and electrolyte disorders, No. (%) | 2783 (22.6%) | 473 (23.5%) | 1273 (21.4%) | 350 (25.1%) | 687 (23.3%) | 0.009 |
| Blood loss anemia, No. (%) | 21 (0.1%) | 4 (0.2%) | 11 (0.1%) | 0 (0.0%) | 6 (0.2%) | 0.434 |
| Deficiency anemias, No. (%) | 6 (0.1%) | 4 (0.2%) | 1 (0.02%) | 1 (0.0%) | 0 (0.0%) | 0.007 |
| Alcohol abuse, No. (%) | 318 (2.5%) | 51 (2.5%) | 133 (2.2%) | 42 (3.0%) | 92 (3.1%) | 0.065 |
| Drug abuse, No. (%) | 23 (0.1%) | 7 (0.3%) | 12 (0.2%) | 1 (0.1%) | 3 (0.1%) | 0.170 |
| Psychoses, No. (%) | 80 (0.6%) | 14 (0.7%) | 40 (0.6%) | 10 (0.7%) | 16 (0.5%) | 0.862 |
| Depression, No. (%) | 50 (0.4%) | 8 (0.4%) | 16 (0.2%) | 4 (0.2%) | 22 (0.7%) | 0.009 |

**Table S19. The associations between comorbidities and subphenotypes in the CEDAR validation cohort**

| Comorbidity Name | Total N=4,804 | DI N=382 | RI N=2,561 | DW N=564 | RW N=1,297 | p-value |
|---|---|---|---|---|---|---|
| Congestive heart failure, No. (%) | 1100 (22.9%) | 94 (24.6%) | 575 (22.4%) | 157 (27.8%) | 274 (21.1%) | 0.012 |
| Valvular disease, No. (%) | 732 (15.2%) | 50 (13.0%) | 408 (15.9%) | 114 (20.2%) | 160 (12.3%) | <0.001 |
| Pulmonary circulation disorder, No. (%) | 690 (14.3%) | 61 (15.9%) | 361 (14.1%) | 103 (18.2%) | 165 (12.7%) | 0.013 |
| Peripheral vascular disorder, No. (%) | 398 (8.2%) | 31 (8.1%) | 203 (7.9%) | 56 (9.9%) | 108 (8.3%) | 0.483 |
| Hypertension, No. (%) | 1776 (36.9%) | 130 (34.0%) | 986 (38.5%) | 221 (39.1%) | 439 (33.8%) | 0.014 |
| Paralysis, No. (%) | 148 (3.0%) | 7 (1.8%) | 85 (3.3%) | 23 (4.0%) | 33 (2.5%) | 0.132 |
| Other neurological, No. (%) | 786 (16.3%) | 63 (16.4%) | 386 (15.0%) | 86 (15.2%) | 251 (19.3%) | 0.007 |
| Chronic pulmonary disease, No. (%) | 835 (17.3%) | 66 (17.2%) | 471 (18.3%) | 118 (20.9%) | 180 (13.8%) | <0.001 |
| Diabetes w/o chronic complications, No. (%) | 678 (14.1%) | 47 (12.3%) | 404 (15.7%) | 85 (15.0%) | 142 (10.9%) | <0.001 |
| Diabetes w/ chronic complications, No. (%) | 328 (6.8%) | 18 (4.7%) | 191 (7.4%) | 40 (7.0%) | 79 (6.0%) | 0.141 |
| Hypothyroidism, No. (%) | 464 (9.6%) | 36 (9.4%) | 251 (9.8%) | 58 (10.2%) | 119 (9.1%) | 0.877 |
| Renal failure, No. (%) | 932 (19.4%) | 77 (20.1%) | 464 (18.1%) | 116 (20.5%) | 275 (21.2%) | 0.111 |
| Liver disease, No. (%) | 402 (8.3%) | 36 (9.4%) | 165 (6.4%) | 48 (8.5%) | 153 (11.8%) | <0.001 |
| Chronic Peptic ulcer disease, No. (%) | 76 (1.5%) | 11 (2.8%) | 41 (1.6%) | 9 (1.6%) | 15 (1.1%) | 0.130 |
| Lymphoma, No. (%) | 217 (4.5%) | 24 (6.2%) | 95 (3.7%) | 29 (5.1%) | 69 (5.3%) | 0.028 |
| Metastatic cancer, No. (%) | 289 (6.0%) | 26 (6.8%) | 127 (4.9%) | 27 (4.7%) | 109 (8.4%) | <0.001 |
| Solid tumor without metastasis, No. (%) | 381 (7.9%) | 38 (9.9%) | 188 (7.3%) | 38 (6.7%) | 117 (9.0%) | 0.087 |
| Rheumatoid arthritis/collagen vascular diseases, No. (%) | 150 (3.1%) | 12 (3.1%) | 79 (3.0%) | 10 (1.7%) | 49 (3.7%) | 0.154 |
| Coagulation deficiency, No. (%) | 931 (19.3%) | 68 (17.8%) | 396 (15.4%) | 121 (21.4%) | 346 (26.6%) | <0.001 |
| Obesity, No. (%) | 286 (5.95%) | 17 (4.4%) | 168 (6.5%) | 38 (6.7%) | 63 (4.8%) | 0.085 |
| Weight loss, No. (%) | 529 (11.0%) | 44 (11.5%) | 290 (11.3%) | 56 (9.9%) | 139 (10.7%) | 0.766 |
| Fluid and electrolyte disorders, No. (%) | 2099 (43.6%) | 162 (42.4%) | 1063 (41.5%) | 259 (45.9%) | 615 (47.4%) | 0.003 |
| Blood loss anemia, No. (%) | 45 (0.9%) | 4 (1.0%) | 22 (0.8%) | 7 (1.2%) | 12 (0.9%) | 0.854 |
| Deficiency anemias, No. (%) | 132 (2.7%) | 11 (2.8%) | 68 (2.6%) | 26 (4.6%) | 27 (2.0%) | 0.023 |
| Alcohol abuse, No. (%) | 172 (3.5%) | 10 (2.6%) | 92 (3.5%) | 21 (3.7%) | 49 (3.7%) | 0.751 |
| Drug abuse, No. (%) | 110 (2.2%) | 6 (1.5%) | 67 (2.6%) | 11 (1.9%) | 26 (2.0%) | 0.413 |
| Psychoses, No. (%) | 55 (1.1%) | 4 (1.0%) | 38 (1.4%) | 6 (1.0%) | 7 (0.5%) | 0.076 |
| Depression, No. (%) | 214 (4.4%) | 15 (3.9%) | 119 (4.6%) | 23 (4.0%) | 57 (4.3%) | 0.880 |

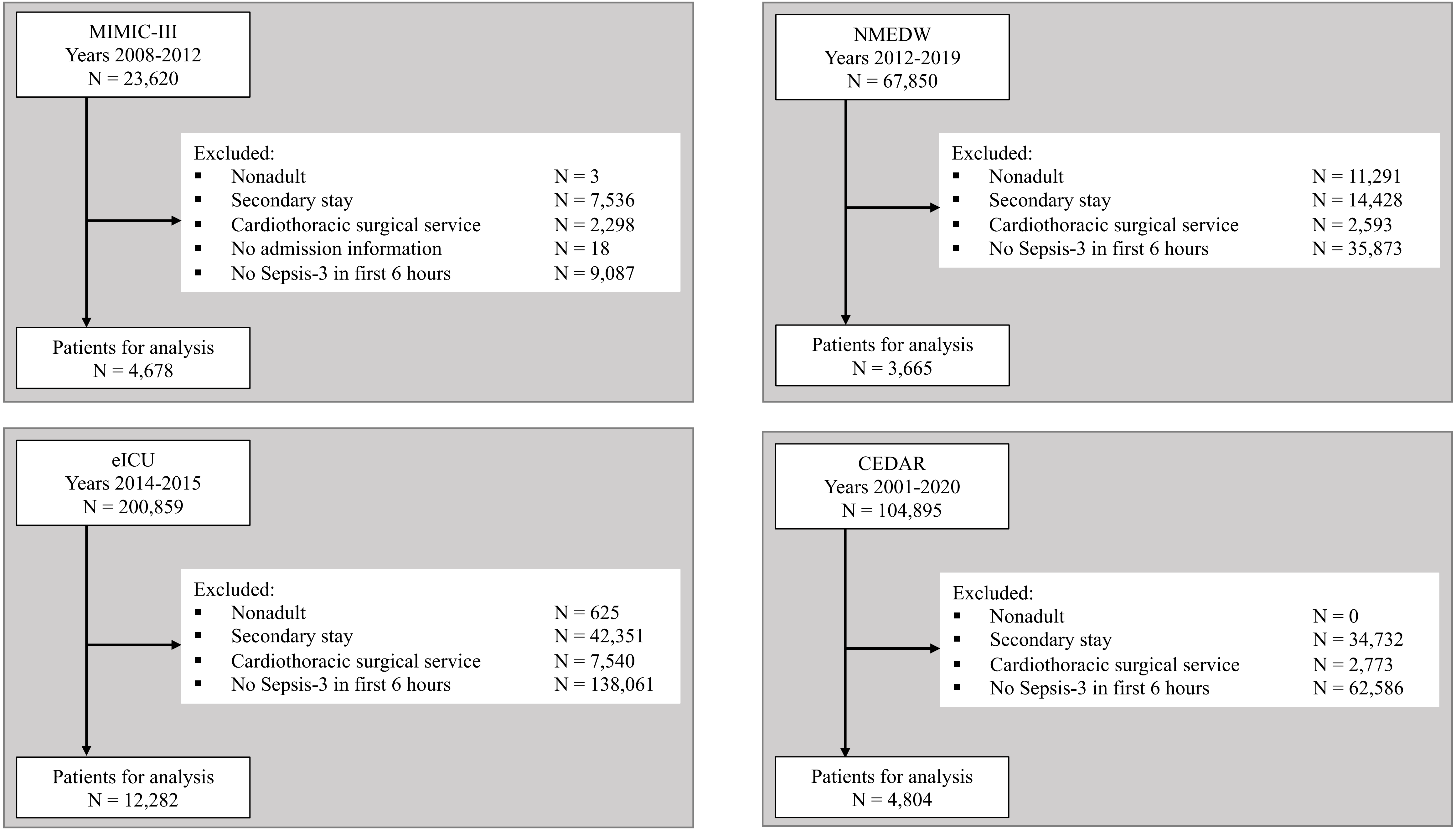

MIMIC-III
Years 2008-2012
N = 23,620
Excluded:
Nonadult N = 3
Secondary stay N = 7,536
Cardiothoracic surgical service N = 2,298
No admission information N = 18
No Sepsis-3 in first 6 hours N = 9,087
Patients for analysis
N = 4,678
NMEDW
Years 2012-2019
N = 67,850
Excluded:
Nonadult N = 11,291
Secondary stay N = 14,428
Cardiothoracic surgical service N = 2,593
No Sepsis-3 in first 6 hours N = 35,873
Patients for analysis
N = 3,665
eICU
Years 2014-2015
N = 200,859
Excluded:
Nonadult N = 625
Secondary stay N = 42,351
Cardiothoracic surgical service N = 7,540
No Sepsis-3 in first 6 hours N = 138,061
Patients for analysis
N = 12,282
CEDAR
Years 2001-2020
N = 104,895
Excluded:
Nonadult N = 0
Secondary stay N = 34,732
Cardiothoracic surgical service N = 2,773
No Sepsis-3 in first 6 hours N = 62,586
Patients for analysis
N = 4,804

**Figure S1. Patient exclusion criteria for building development and validation cohorts**

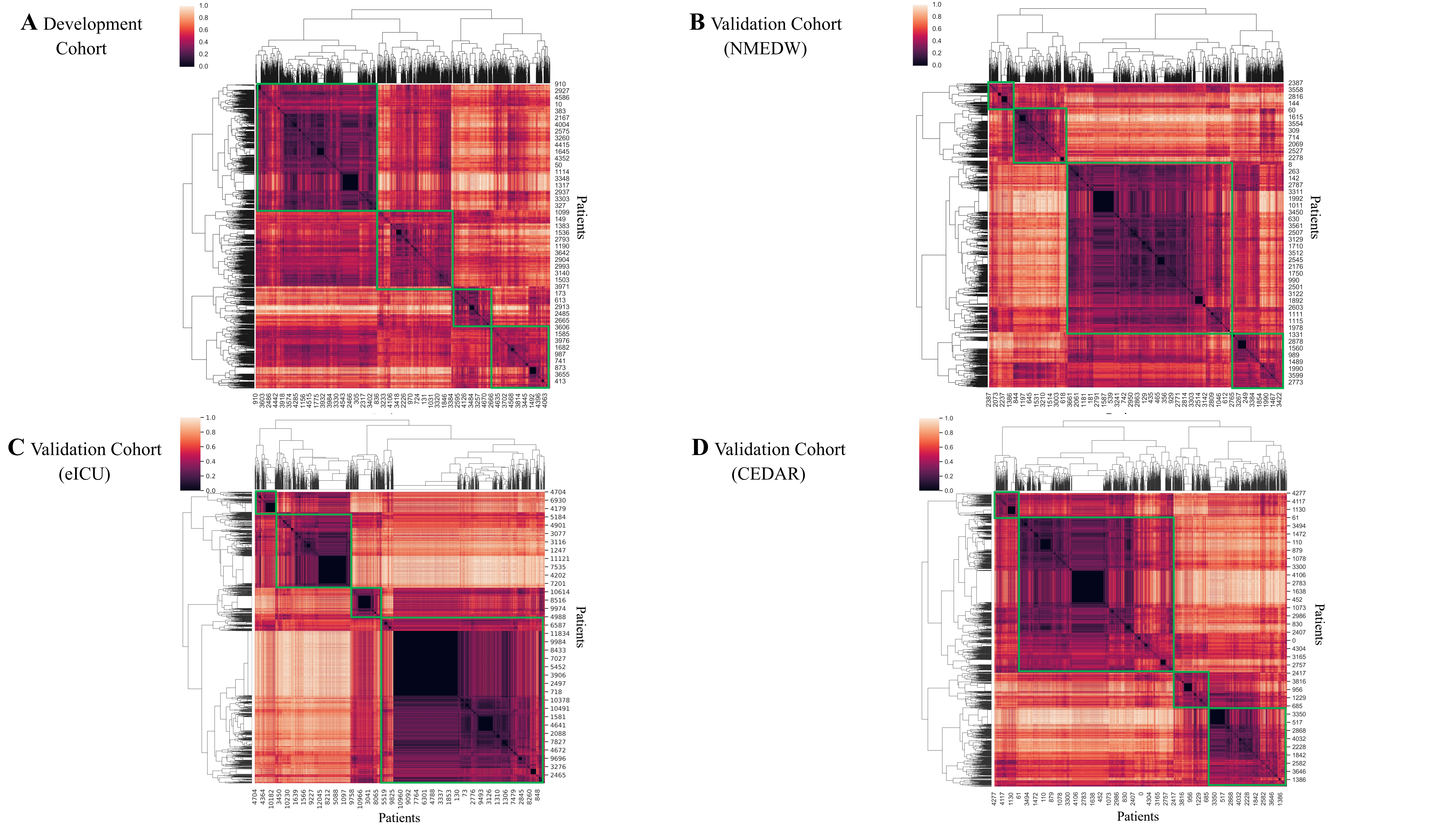

A Development Cohort
B Validation Cohort (NMEDW)
C Validation Cohort (eICU)
D Validation Cohort (CEDAR)
1.0
0.8
0.6
0.4
0.2
0.0
Patients

**Figure S2. Clustergrams of hierarchical clustering. Horizontal and vertical axes represent patients**. Color intensity denotes normalized pairwise patient similarity derived using Dynamic Time Warping (DTW). All clustergrams suggest optimal cluster number 4.

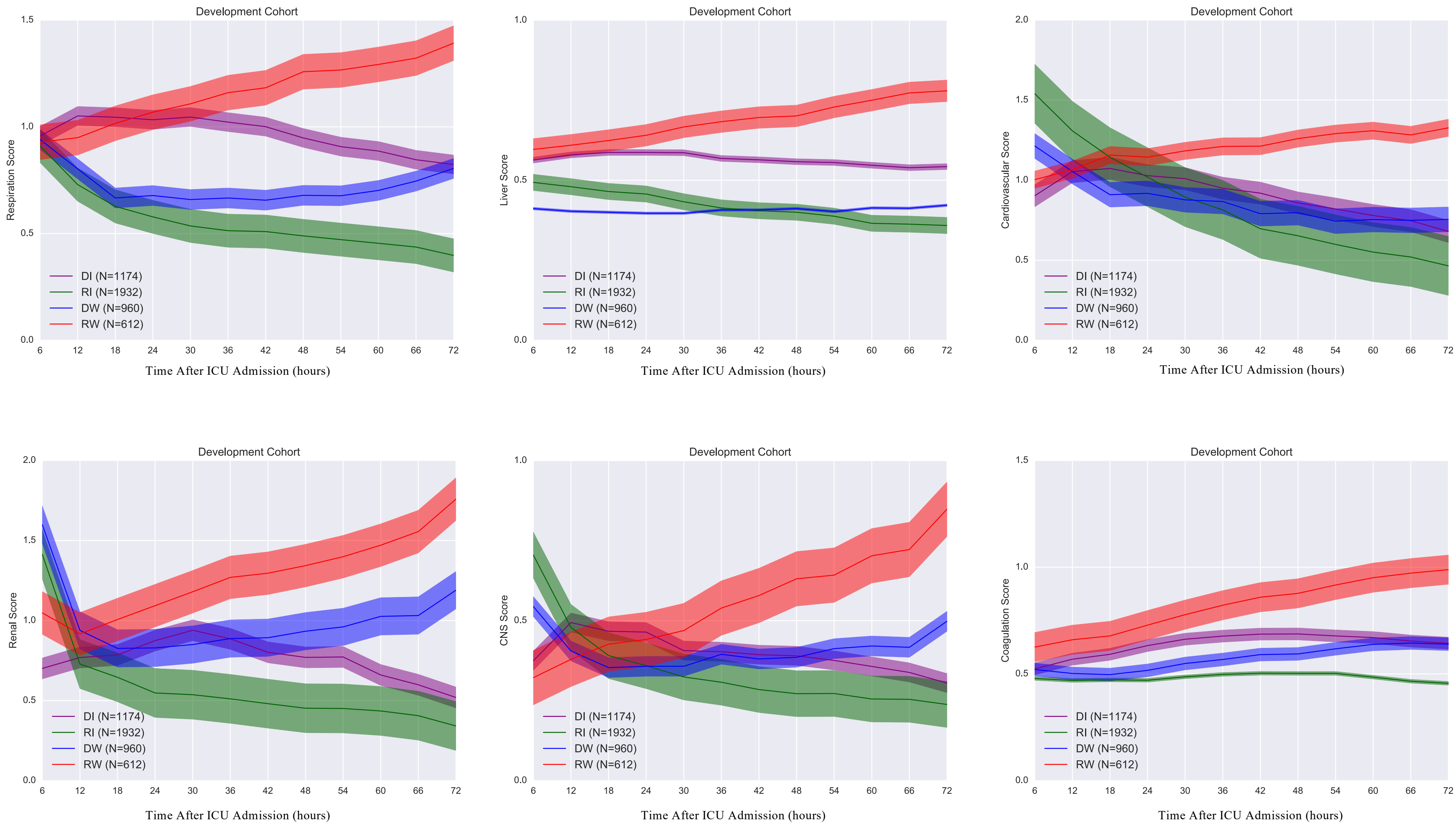
Development Cohort
Respiration Score
Liver Score
Cardiovascular Score
Renal Score
CNS Score
Coagulation Score
DI (N=1174)
RI (N=1932)
DW (N=960)
RW (N=612)
Time After ICU Admission (hours)

**Figure S3. The trajectories of the subphenotypes in terms of subscores in the development cohort**. DI: Delayed Improving; RI: Rapidly Improving; DW: Delayed Worsening; RW: Rapidly Worsening.

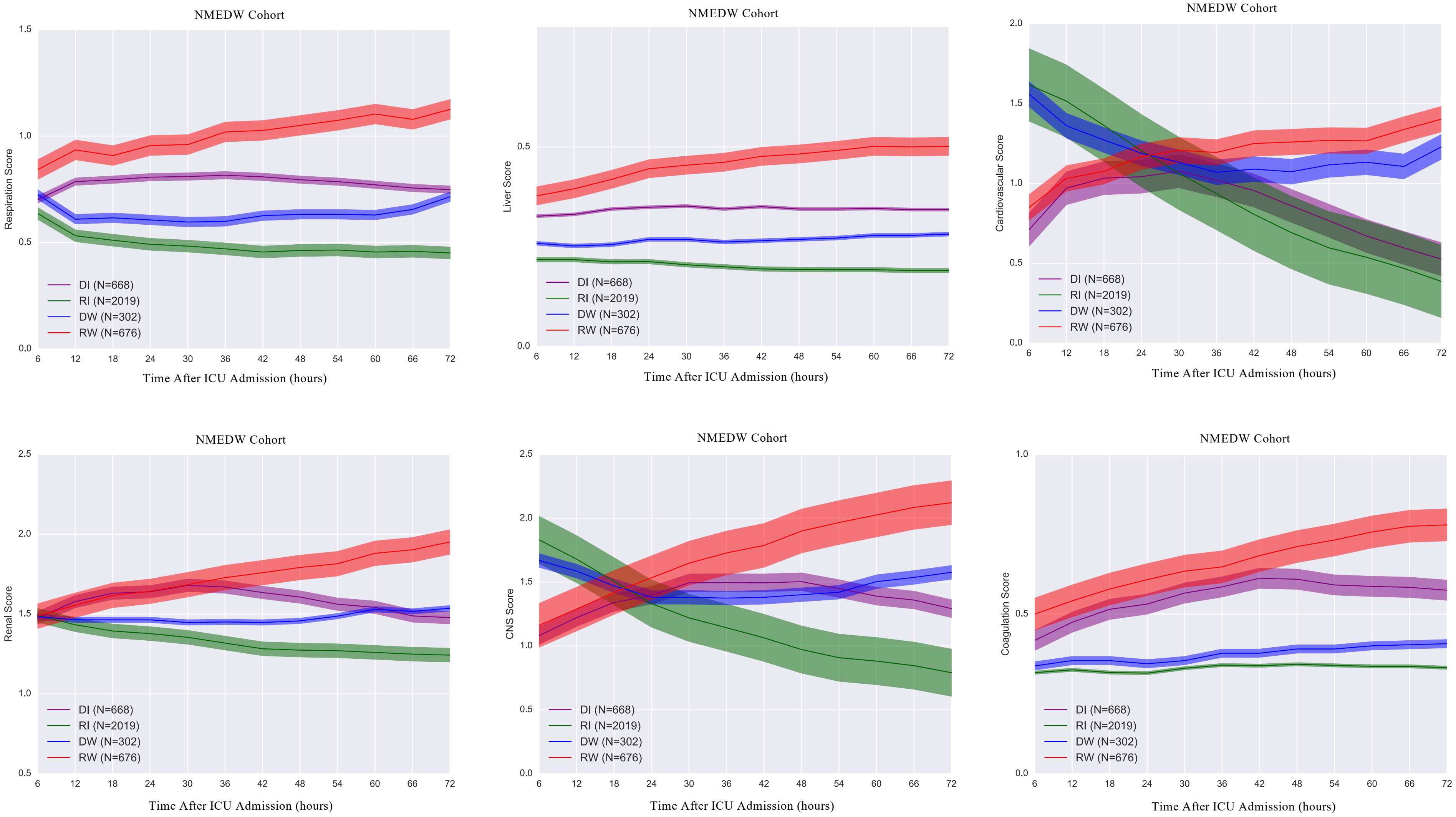
NMEDW Cohort
Respiration Score
Time After ICU Admission (hours)
DI (N=668)
RI (N=2019)
DW (N=302)
RW (N=676)
NMEDW Cohort
Liver Score
Time After ICU Admission (hours)
DI (N=668)
RI (N=2019)
DW (N=302)
RW (N=676)
NMEDW Cohort
Cardiovascular Score
Time After ICU Admission (hours)
DI (N=668)
RI (N=2019)
DW (N=302)
RW (N=676)
NMEDW Cohort
Renal Score
Time After ICU Admission (hours)
DI (N=668)
RI (N=2019)
DW (N=302)
RW (N=676)
NMEDW Cohort
CNS Score
Time After ICU Admission (hours)
DI (N=668)
RI (N=2019)
DW (N=302)
RW (N=676)
NMEDW Cohort
Coagulation Score
Time After ICU Admission (hours)
DI (N=668)
RI (N=2019)
DW (N=302)
RW (N=676)

**Figure S4. The trajectories of the subphenotypes in terms of subscores in the NMEDW validation cohort**. DI: Delayed Improving; RI: Rapidly Improving; DW: Delayed Worsening; RW: Rapidly Worsening.

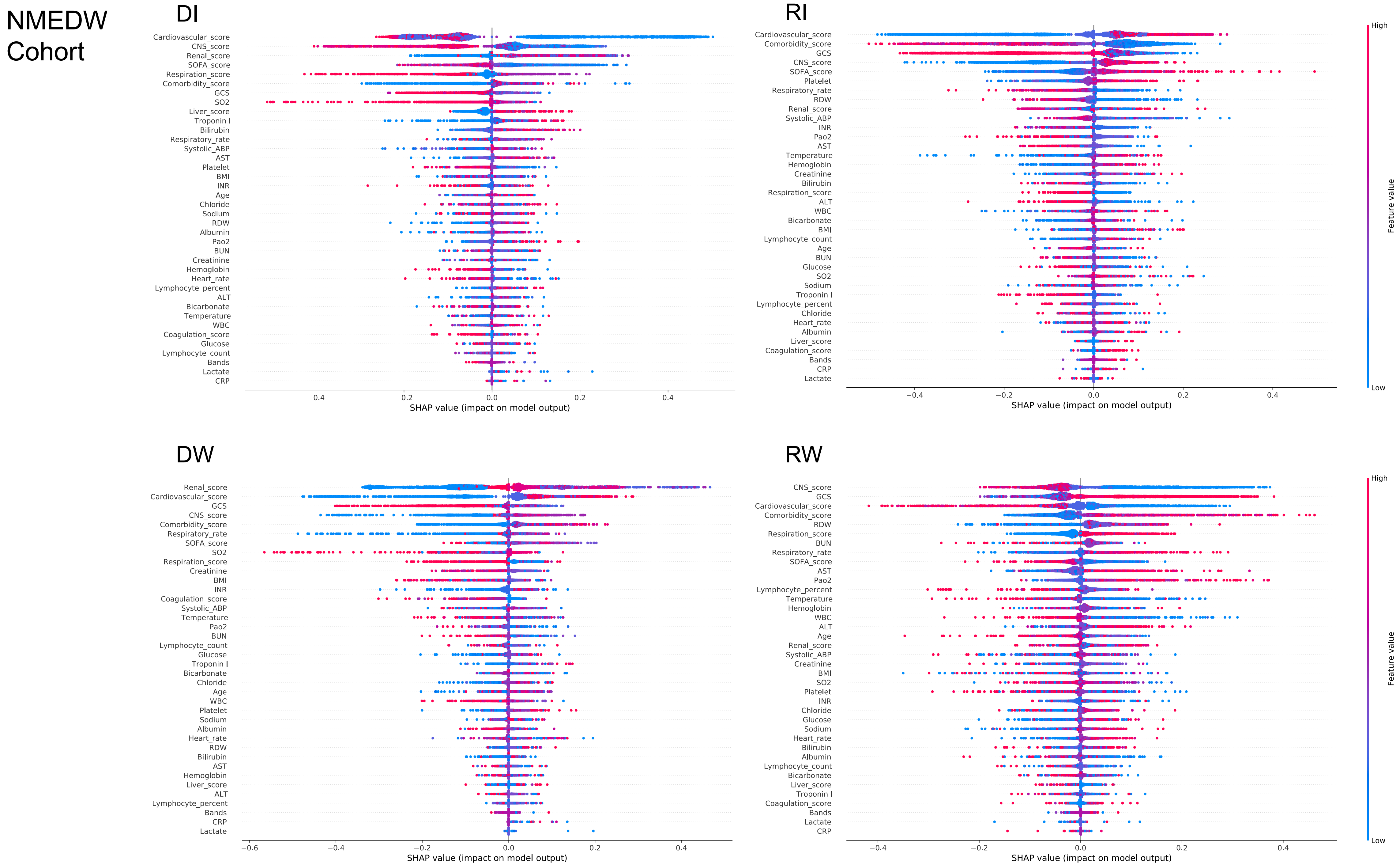
NMEDW
Cohort
DI
Cardiovascular_score
CNS_score
Renal_score
SOFA_score
Respiration_score
Comorbidity_score
GCS
SO2
Liver_score
Troponin I
Bilirubin
Respiratory_rate
Systolic_ABP
AST
Platelet
BMI
INR
Age
Chloride
Sodium
RDW
Albumin
Pao2
BUN
Creatinine
Hemoglobin
Heart_rate
Lymphocyte_percent
ALT
Bicarbonate
Temperature
WBC
Coagulation_score
Glucose
Lymphocyte_count
Bands
Lactate
CRP
−0.4 −0.2 0.0 0.2 0.4
SHAP value (impact on model output)
RI
Cardiovascular_score
Comorbidity_score
GCS
CNS_score
SOFA_score
Platelet
Respiratory_rate
RDW
Renal_score
Systolic_ABP
INR
Pao2
AST
Temperature
Hemoglobin
Creatinine
Bilirubin
Respiration_score
ALT
WBC
Bicarbonate
BMI
Lymphocyte_count
Age
BUN
Glucose
SO2
Sodium
Troponin I
Lymphocyte_percent
Chloride
Heart_rate
Albumin
Liver_score
Coagulation_score
Bands
CRP
Lactate
−0.4 −0.2 0.0 0.2 0.4
SHAP value (impact on model output)
High
Feature value
Low
DW
Renal_score
Cardiovascular_score
GCS
CNS_score
Comorbidity_score
Respiratory_rate
SOFA_score
SO2
Respiration_score
Creatinine
BMI
INR
Coagulation_score
Systolic_ABP
Temperature
Pao2
BUN
Lymphocyte_count
Glucose
Troponin I
Bicarbonate
Chloride
Age
WBC
Platelet
Sodium
Albumin
Heart_rate
RDW
Bilirubin
AST
Hemoglobin
Liver_score
ALT
Lymphocyte_percent
Bands
CRP
Lactate
−0.6 −0.4 −0.2 0.0 0.2 0.4
SHAP value (impact on model output)
RW
CNS_score
GCS
Cardiovascular_score
Comorbidity_score
RDW
Respiration_score
BUN
Respiratory_rate
SOFA_score
AST
Pao2
Lymphocyte_percent
Temperature
Hemoglobin
WBC
ALT
Age
Renal_score
Systolic_ABP
Creatinine
BMI
SO2
Platelet
INR
Chloride
Glucose
Sodium
Heart_rate
Bilirubin
Albumin
Lymphocyte_count
Bicarbonate
Liver_score
Troponin I
Coagulation_score
Bands
Lactate
CRP
−0.4 −0.2 0.0 0.2 0.4
SHAP value (impact on model output)
High
Feature value
Low

**Figure S5. Predictor contribution to the prediction of the predictive model in the NMEDW validation cohor**t. DI: Delayed Improving; RI: Rapidly Improving; DW: Delayed Worsening; RW: Rapidly Worsening.

Validation Cohort (NMEDW)

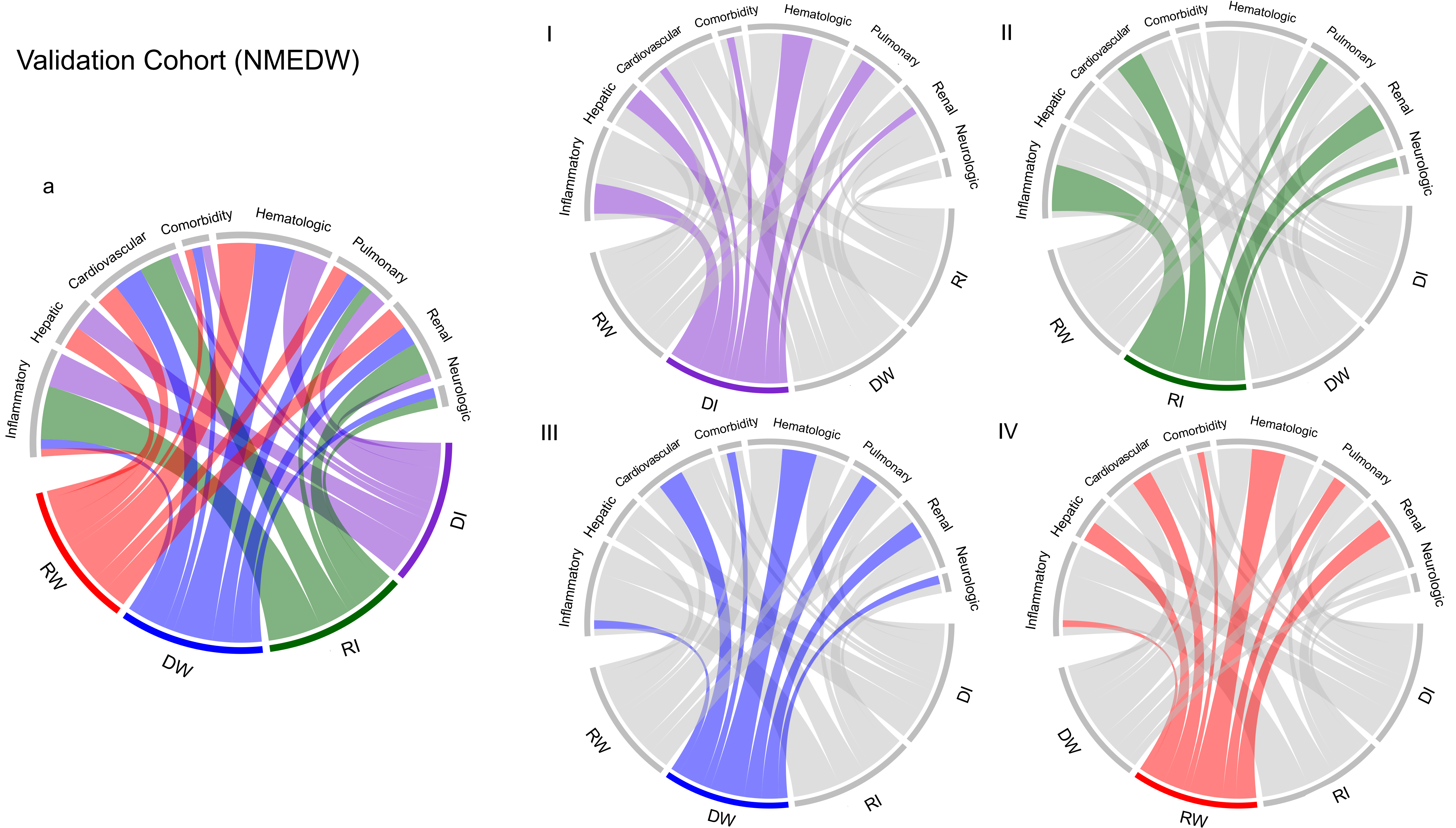

**Figure S6. Chord diagrams showing abnormal clinical variables by subphenotype in the NMEDW validation cohort**. a: abnormal biomarkers vs. all subphenotypes; I: abnormal biomarkers vs. DI; II: abnormal biomarkers vs. RI; III: abnormal biomarkers vs. DW; IV: abnormal biomarkers vs. RW. DI: Delayed Improving; RI: Rapidly Improving; DW: Delayed Worsening; RW: Rapidly Worsening.

### Validation Cohort (NMEDW)

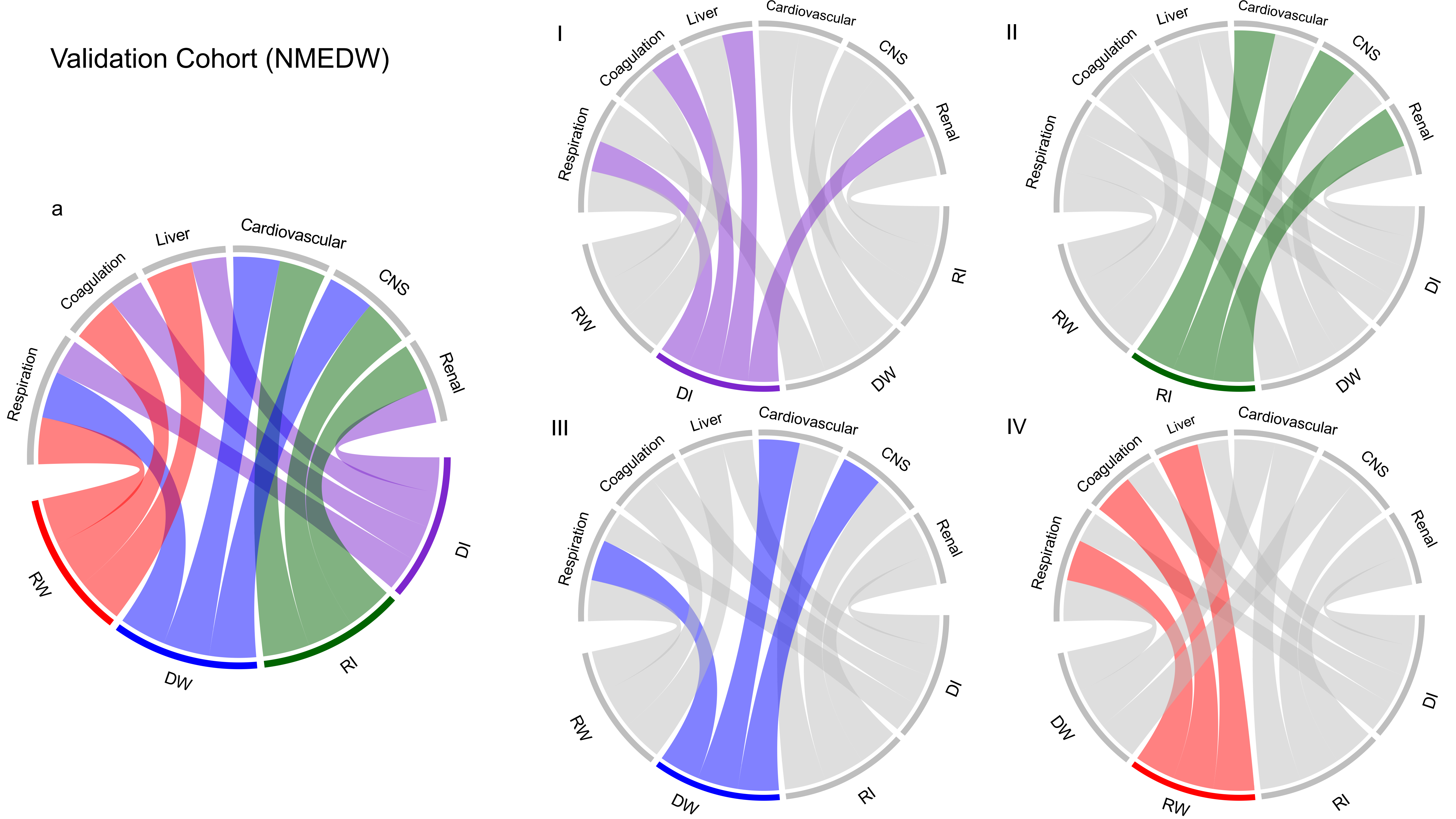

**Figure S7. Chord diagrams showing abnormal subscores by subphenotype in the NMEDW validation cohort**. a: abnormal subscores vs. all subphenotypes; I: abnormal subscores vs. DI; II: abnormal subscores vs. RI; III: abnormal subscores vs. DW; IV: abnormal subscores vs. RW. DI: Delayed Improving; RI: Rapidly Improving; DW: Delayed Worsening; RW: Rapidly Worsening.

A

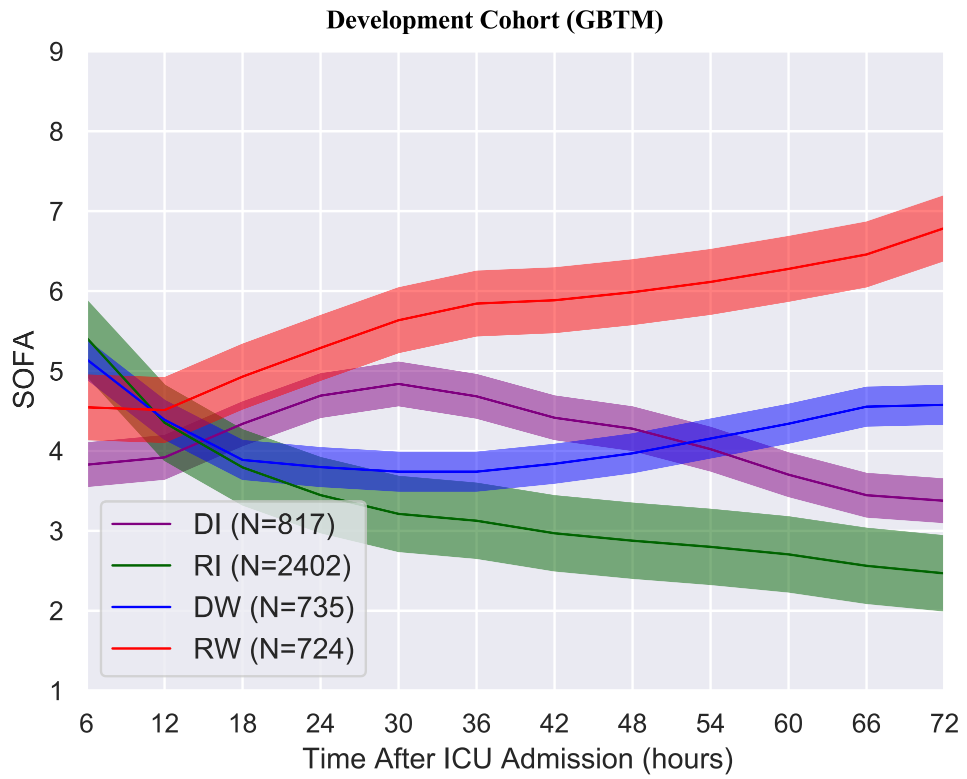
Development Cohort (GBTM)
SOFA
Time After ICU Admission (hours)
DI (N=817)
RI (N=2402)
DW (N=735)
RW (N=724)
9
8
7
6
5
4
3
2
1
6
12
18
24
30
36
42
48
54
60
66
72

B

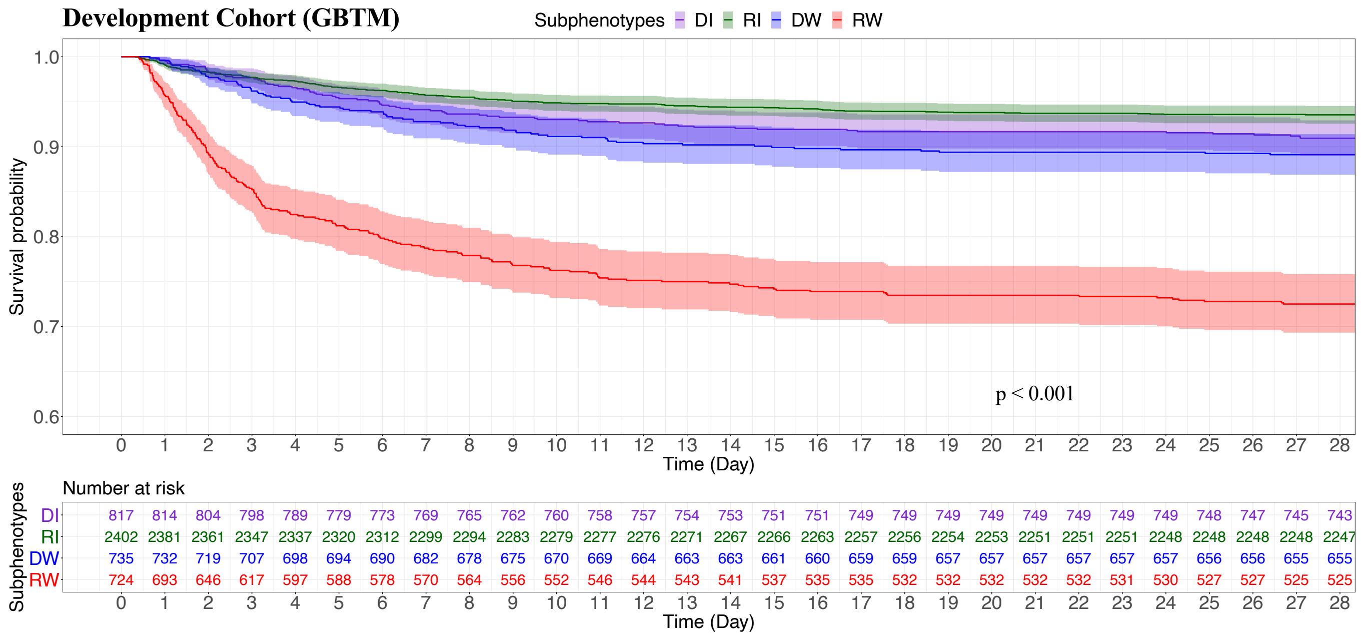
Development Cohort (GBTM)
Subphenotypes DI RI DW RW
Survival probability
1.0
0.9
0.8
0.7
0.6
p < 0.001
Time (Day)
Number at risk
Subphenotypes
DI 817 814 804 798 789 779 773 769 765 762 760 758 757 754 753 751 751 749 749 749 749 749 749 749 749 748 747 745 743
RI 2402 2381 2361 2347 2337 2320 2312 2299 2294 2283 2279 2277 2276 2271 2267 2266 2263 2257 2256 2254 2253 2251 2251 2251 2248 2248 2248 2248 2247
DW 735 732 719 707 698 694 690 682 678 675 670 669 664 663 663 661 660 659 659 657 657 657 657 657 657 656 656 655 655
RW 724 693 646 617 597 588 578 570 564 556 552 546 544 543 541 537 535 535 532 532 532 532 532 531 530 527 527 525 525
0 1 2 3 4 5 6 7 8 9 10 11 12 13 14 15 16 17 18 19 20 21 22 23 24 25 26 27 28
Time (Day)

**Figure S8. Sequential Organ Failure Assessment (SOFA) trajectories of the subphenotypes and survival analysis in terms of the identified subphenotypes by GBTM**. The (A) describes the SOFA trajectories of the subphenotypes re-derived by GBTM in development cohort. The (B) shows the survival analysis results in subphenotypes re-derived by GBTM in the development cohort. DI: Delayed Improving; RI: Rapidly Improving; DW: Delayed Worsening; RW: Rapidly Worsening.

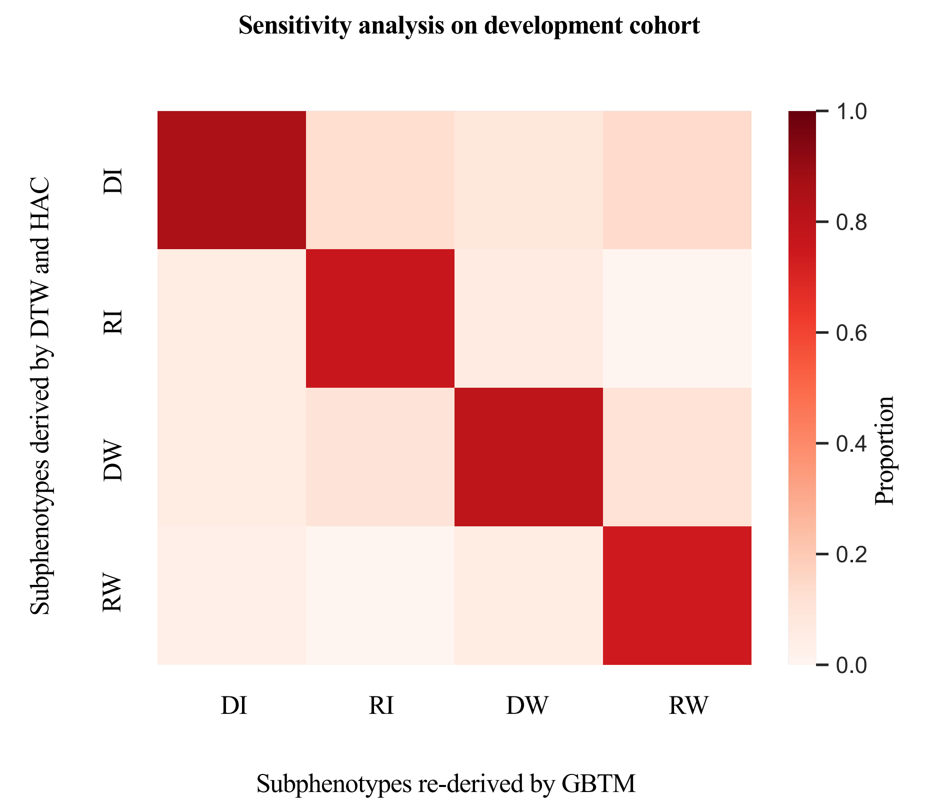

Sensitivity analysis on development cohort
Subphenotypes derived by DTW and HAC
DI
RI
DW
RW
Subphenotypes re-derived by GBTM
DI
RI
DW
RW
Proportion
1.0
0.8
0.6
0.4
0.2
0.0

**Figure S9. Confusion matrices for comparing the subphenotypes obtained by DTW and HAC, and GBTM in development cohort**. DI: Delayed Improving; RI: Rapidly Improving; DW: Delayed Worsening; RW: Rapidly Worsening. DTW: Dynamic Time Warping; HAC: Hierarchical Agglomerative Clustering. GBTM: Group-Based Trajectory Modeling.

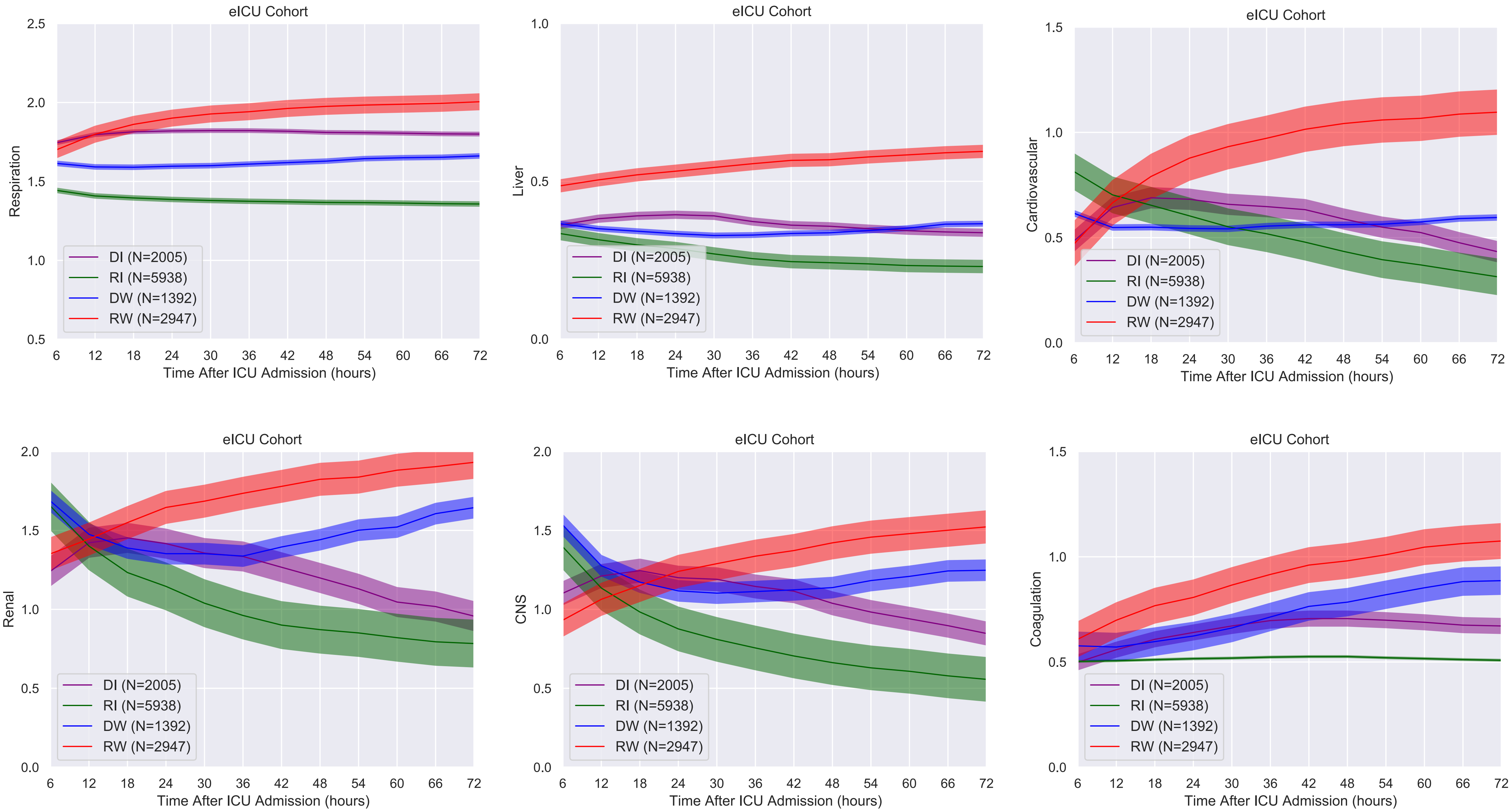

eICU Cohort
Respiration
Liver
Cardiovascular
Renal
CNS
Coagulation
DI (N=2005)
RI (N=5938)
DW (N=1392)
RW (N=2947)
Time After ICU Admission (hours)

**Figure S10. The trajectories of the subphenotypes in terms of subscores in the eICU validation cohort**. DI: Delayed Improving; RI: Rapidly Improving; DW: Delayed Worsening; RW: Rapidly Worsening.

eICU
Cohort

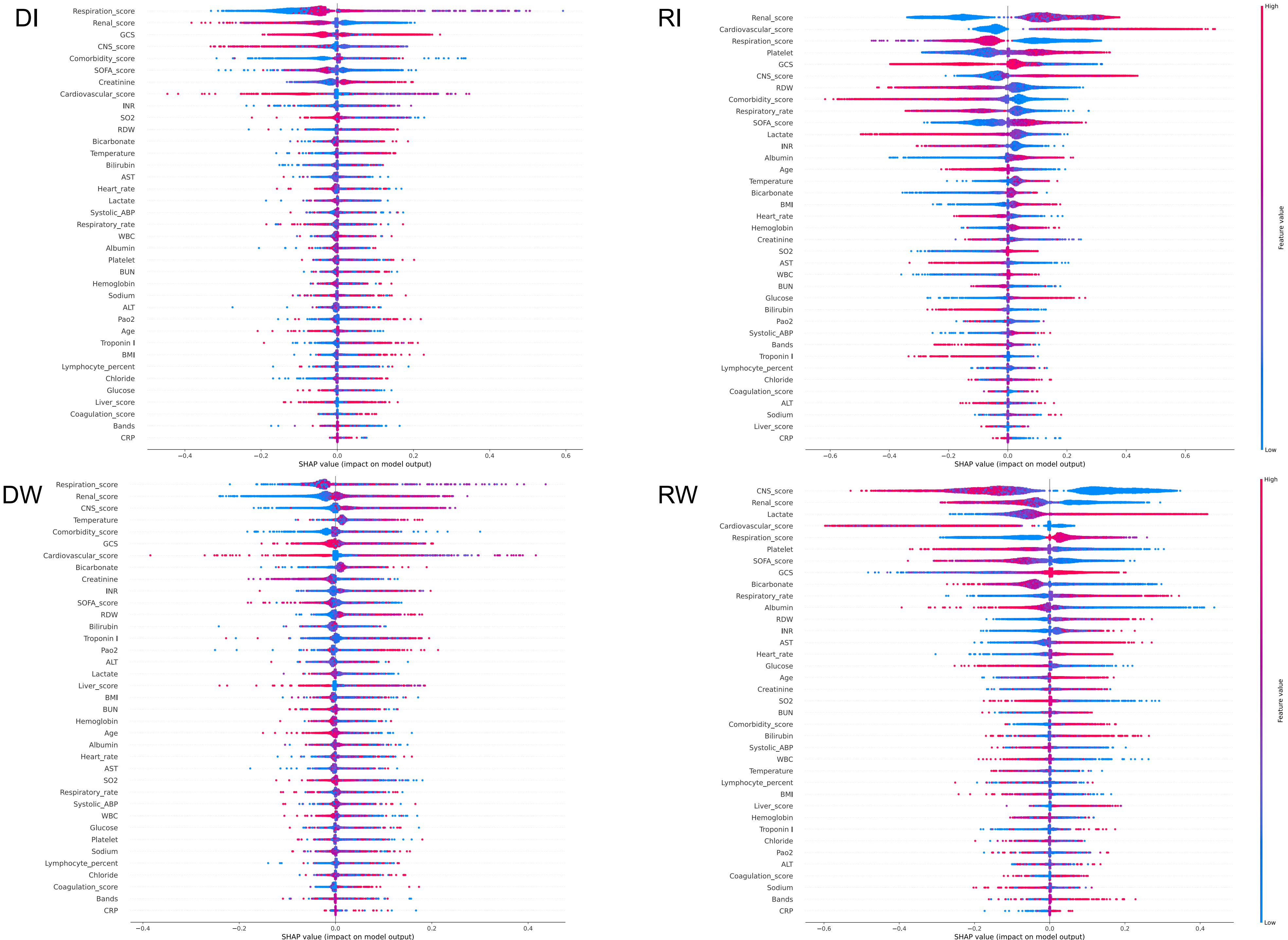
DI
Respiration_score
Renal_score
GCS
CNS_score
Comorbidity_score
SOFA_score
Creatinine
Cardiovascular_score
INR
SO2
RDW
Bicarbonate
Temperature
Bilirubin
AST
Heart_rate
Lactate
Systolic_ABP
Respiratory_rate
WBC
Albumin
Platelet
BUN
Hemoglobin
Sodium
ALT
Pao2
Age
Troponin I
BMI
Lymphocyte_percent
Chloride
Glucose
Liver_score
Coagulation_score
Bands
CRP
−0.4 −0.2 0.0 0.2 0.4 0.6
SHAP value (impact on model output)
RI
Renal_score
Cardiovascular_score
Respiration_score
Platelet
GCS
CNS_score
RDW
Comorbidity_score
Respiratory_rate
SOFA_score
Lactate
INR
Albumin
Age
Temperature
Bicarbonate
BMI
Heart_rate
Hemoglobin
Creatinine
SO2
AST
WBC
BUN
Glucose
Bilirubin
Pao2
Systolic_ABP
Bands
Troponin I
Lymphocyte_percent
Chloride
Coagulation_score
ALT
Sodium
Liver_score
CRP
−0.6 −0.4 −0.2 0.0 0.2 0.4 0.6
SHAP value (impact on model output)
High
Feature value
Low
DW
Respiration_score
Renal_score
CNS_score
Temperature
Comorbidity_score
GCS
Cardiovascular_score
Bicarbonate
Creatinine
INR
SOFA_score
RDW
Bilirubin
Troponin I
Pao2
ALT
Lactate
Liver_score
BMI
BUN
Hemoglobin
Age
Albumin
Heart_rate
AST
SO2
Respiratory_rate
Systolic_ABP
WBC
Glucose
Platelet
Sodium
Lymphocyte_percent
Chloride
Coagulation_score
Bands
CRP
−0.4 −0.2 0.0 0.2 0.4
SHAP value (impact on model output)
RW
CNS_score
Renal_score
Lactate
Cardiovascular_score
Respiration_score
Platelet
SOFA_score
GCS
Bicarbonate
Respiratory_rate
Albumin
RDW
INR
AST
Heart_rate
Glucose
Age
Creatinine
SO2
BUN
Comorbidity_score
Bilirubin
Systolic_ABP
WBC
Temperature
Lymphocyte_percent
BMI
Liver_score
Hemoglobin
Troponin I
Chloride
Pao2
ALT
Coagulation_score
Sodium
Bands
CRP
−0.6 −0.4 −0.2 0.0 0.2 0.4
SHAP value (impact on model output)
High
Feature value
Low

**Figure S11. Predictor contribution to the prediction of the predictive model in the eICU validation cohort**. DI: Delayed Improving; RI: Rapidly Improving; DW: Delayed Worsening; RW: Rapidly Worsening.

### Validation Cohort (eICU)

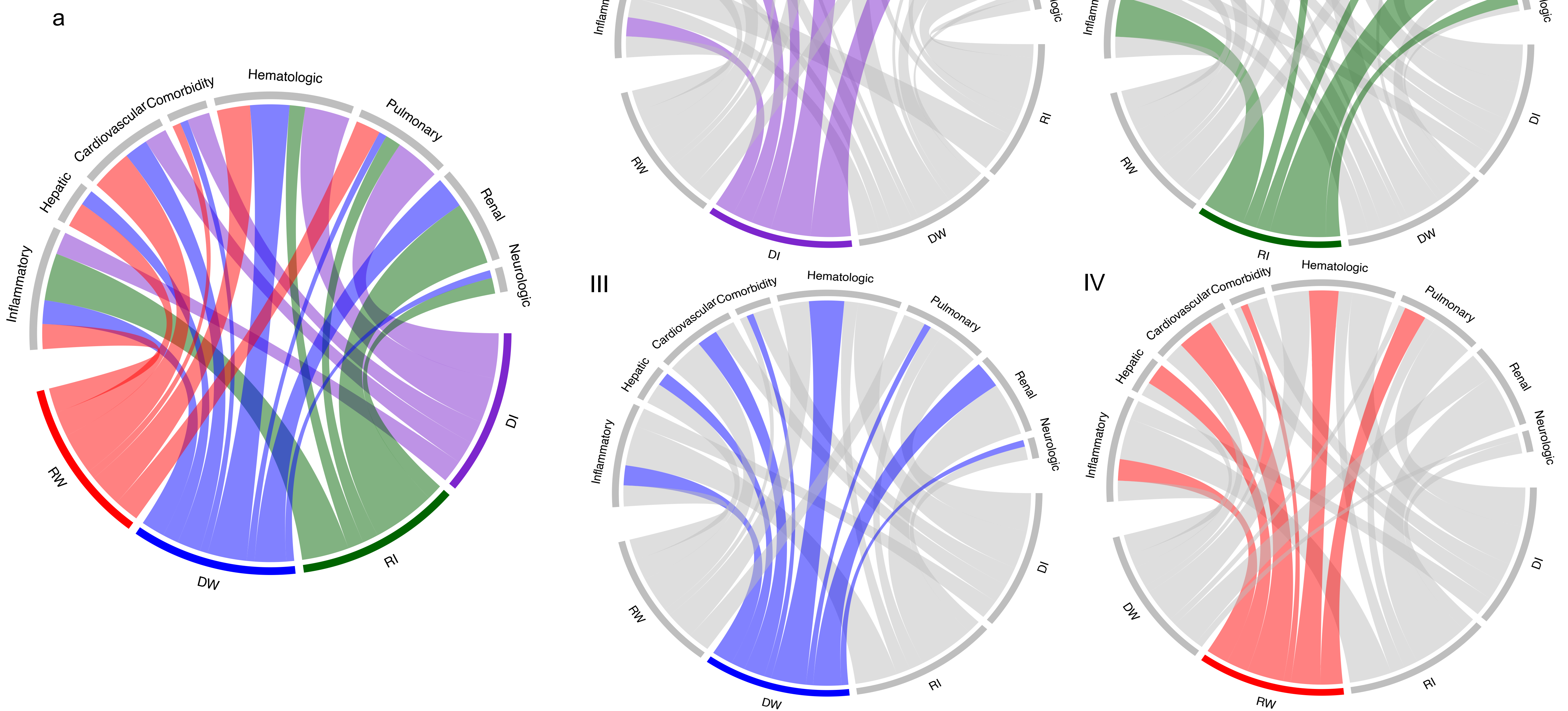

**Figure S12. Chord diagrams showing abnormal clinical variables by subphenotype in the eICU validation cohort**. a: abnormal biomarkers vs. all subphenotypes; I: abnormal biomarkers vs. DI; II: abnormal biomarkers vs. RI; III: abnormal biomarkers vs. DW; IV: abnormal biomarkers vs. RW. DI: Delayed Improving; RI: Rapidly Improving; DW: Delayed Worsening; RW: Rapidly Worsening.

Validation Cohort (eICU)

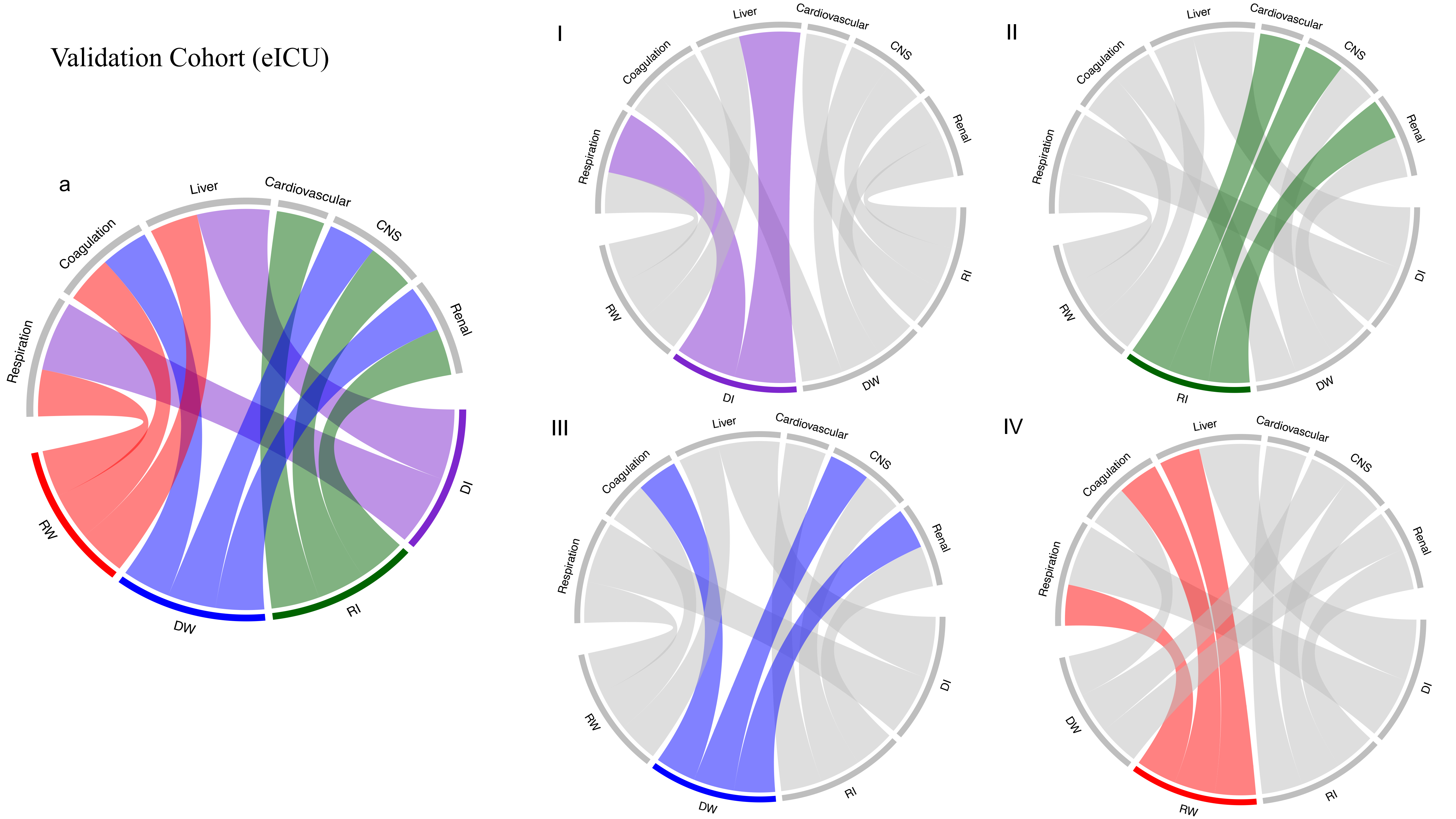

**Figure S13. Chord diagrams showing abnormal subscores by subphenotype in the eICU validation cohort**. a: abnormal subscores vs. all subphenotypes; I: abnormal subscores vs. DI; II: abnormal subscores vs. RI; III: abnormal subscores vs. DW; IV: abnormal subscores vs. RW. DI: Delayed Improving; RI: Rapidly Improving; DW: Delayed Worsening; RW: Rapidly Worsening.

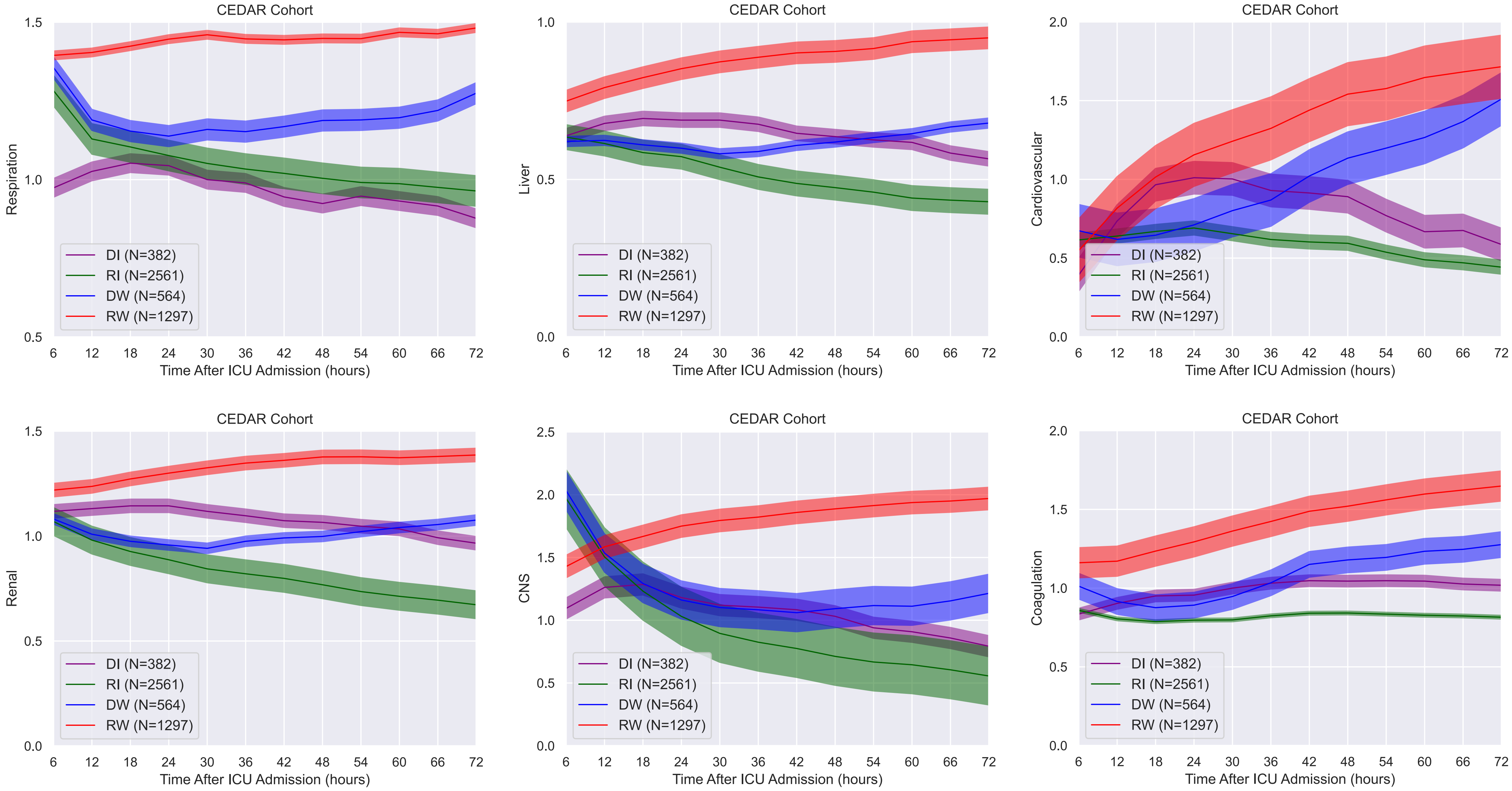
CEDAR Cohort
Respiration
Liver
Cardiovascular
Renal
CNS
Coagulation
Time After ICU Admission (hours)
DI (N=382)
RI (N=2561)
DW (N=564)
RW (N=1297)
6
12
18
24
30
36
42
48
54
60
66
72

**Figure S14. The trajectories of the subphenotypes in terms of subscores in the CEDAR validation cohort. DI**: Delayed Improving; RI: Rapidly Improving; DW: Delayed Worsening; RW: Rapidly Worsening.

CEDAR
Cohort

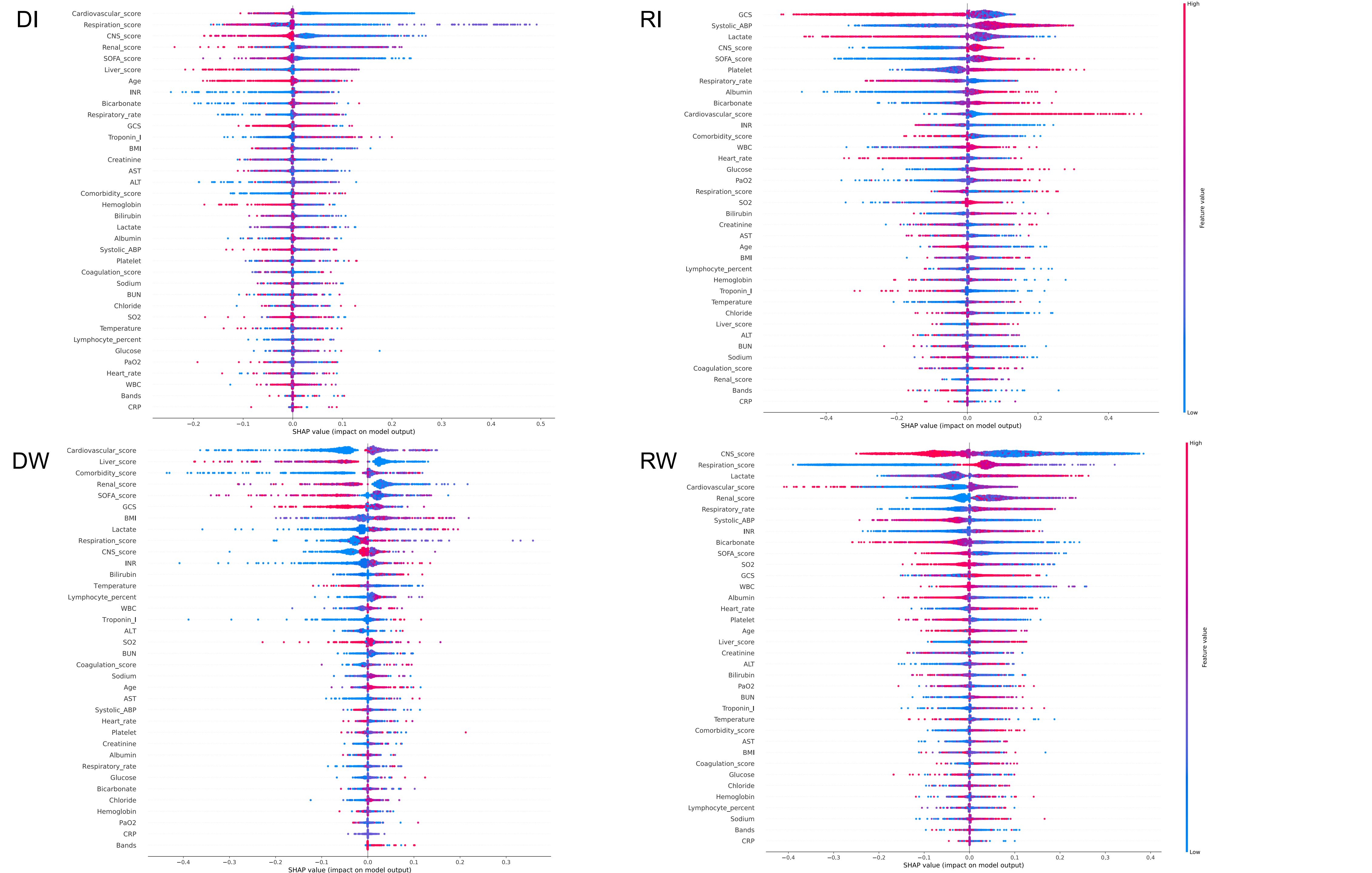

**Figure S15. Predictor contribution to the prediction of the predictive model in the CEDAR validation cohort**.
DI: Delayed Improving; RI: Rapidly Improving; DW: Delayed Worsening; RW: Rapidly Worsening.

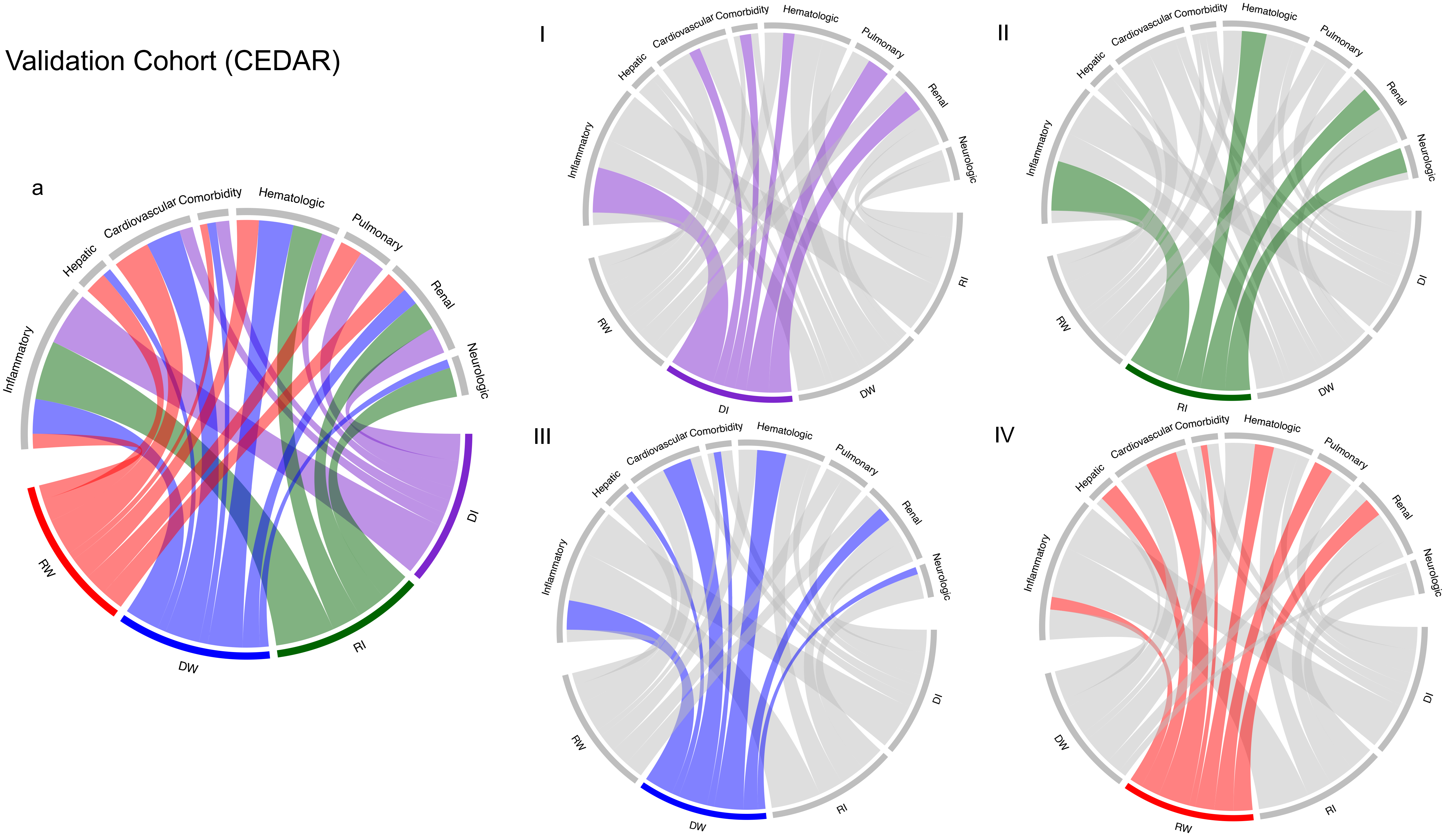
Validation Cohort (CEDAR)
a
Inflammatory
Hepatic
Cardiovascular Comorbidity
Hematologic
Pulmonary
Renal
Neurologic
DI
RI
DW
RW
I
II
III
IV

**Figure S16. Chord diagrams showing abnormal clinical variables by subphenotype in the CEDAR validation cohort.** a: abnormal biomarkers vs. all subphenotypes; I: abnormal biomarkers vs. DI; II: abnormal biomarkers vs. RI; III: abnormal biomarkers vs. DW; IV: abnormal biomarkers vs. RW. DI: Delayed Improving; RI: Rapidly Improving; DW: Delayed Worsening; RW: Rapidly Worsening.

### Validation Cohort (CEDAR)

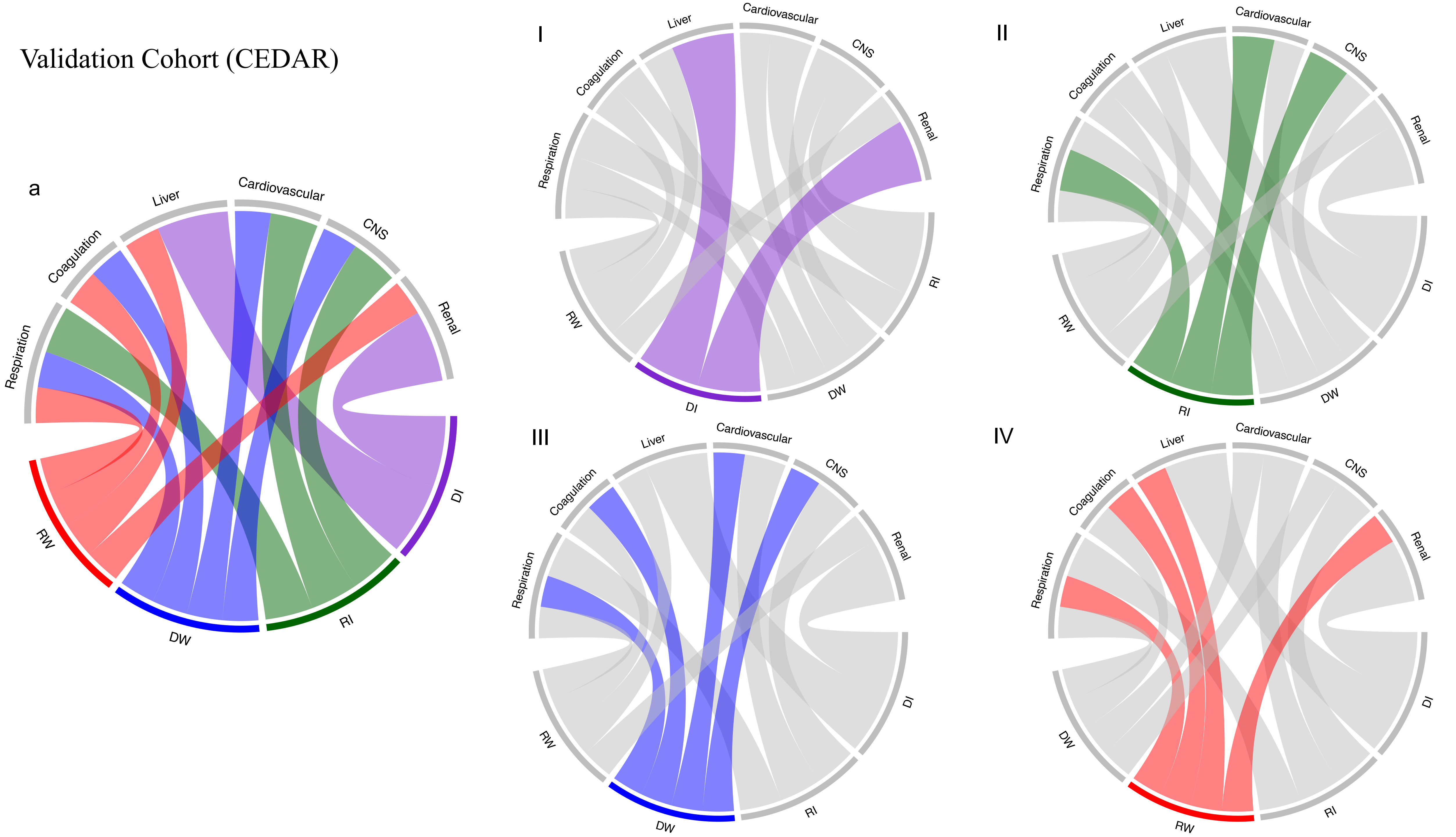

**Figure S17. Chord diagrams showing abnormal subscores by subphenotype in the CEDAR validation cohort.** a: abnormal subscores vs. all subphenotypes; I: abnormal subscores vs. DI; II: abnormal subscores vs. RI; III: abnormal subscores vs. DW; IV: abnormal subscores vs. RW. DI: Delayed Improving; RI: Rapidly Improving; DW: Delayed Worsening; RW: Rapidly Worsening.

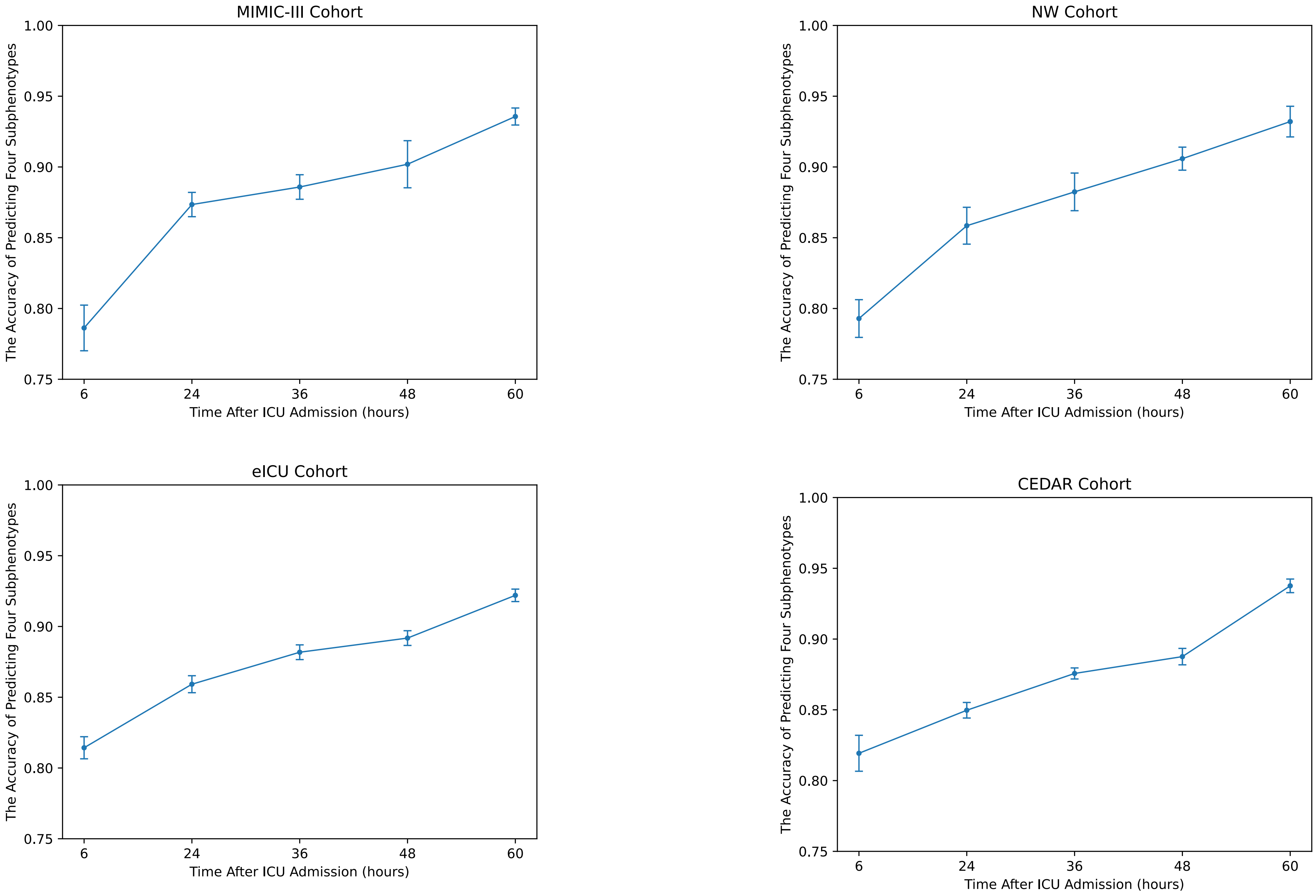
MIMIC-III Cohort
NW Cohort
eICU Cohort
CEDAR Cohort
The Accuracy of Predicting Four Subphenotypes
Time After ICU Admission (hours)
1.00
0.95
0.90
0.85
0.80
0.75
6
24
36
48
60

**Figure S18. The accuracy of predicting four subphenotypes at successive time points (hours 6, 24, 36, 48, 60) after ICU admission in development and validation cohorts.**

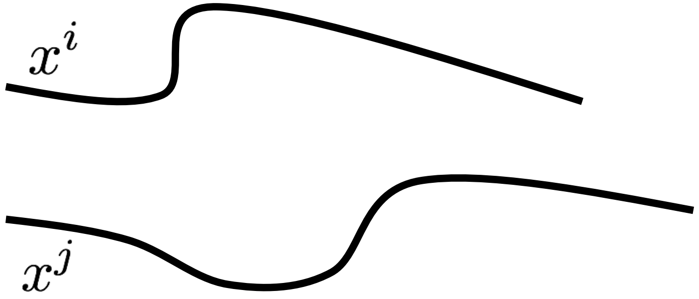


(a) Original Sequences

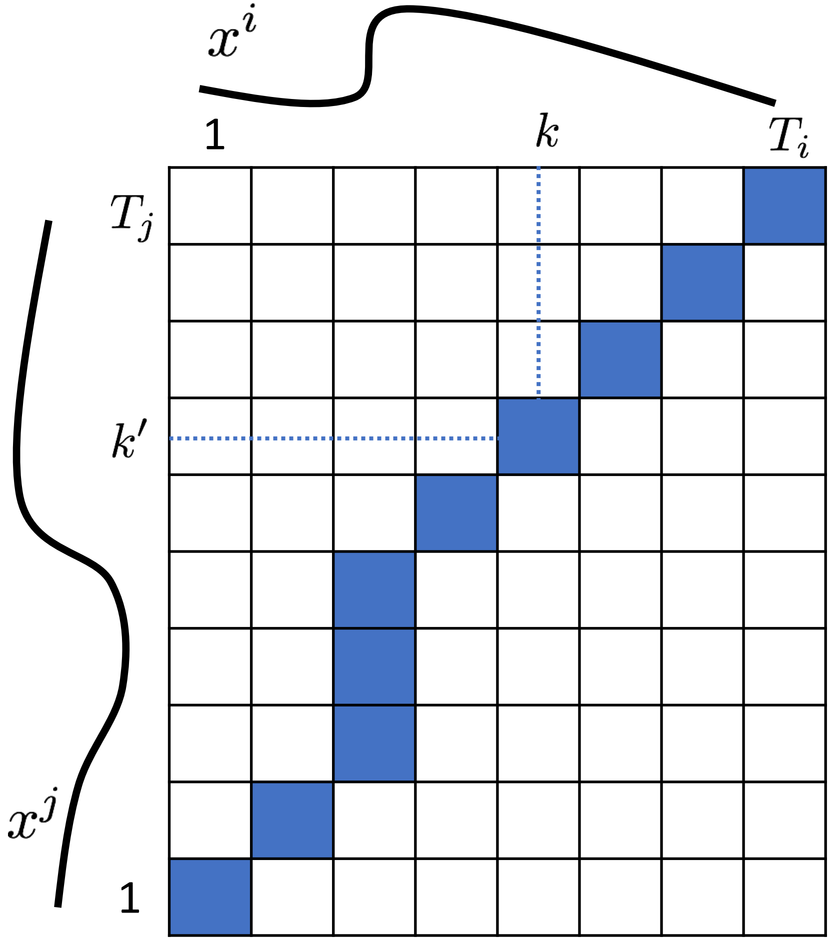


(b) Warping Path

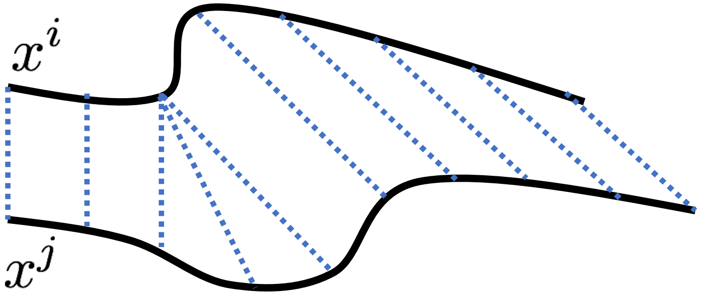


(c) DTW Matching

**Figure S19. The illustration of using DTW during obtaining matched sequences. (c) shows the warping path induced by the DTW matching, where the blue cells indicate the best matching pairs for the points in the two sequences.**

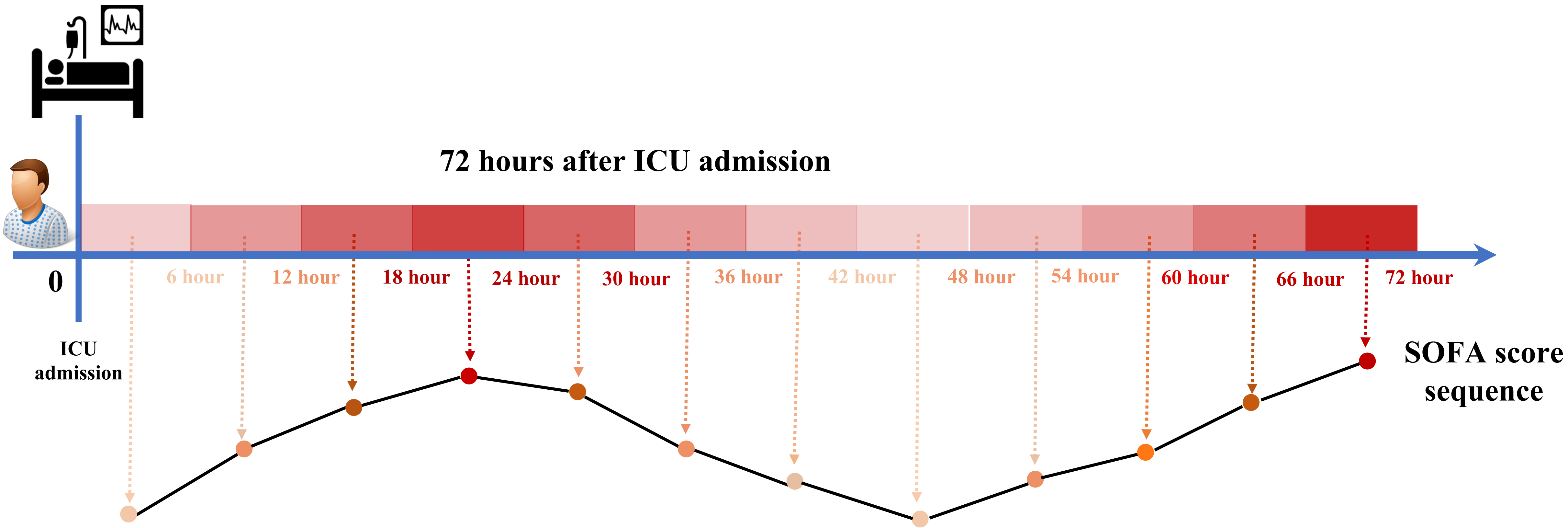

72 hours after ICU admission
0
6 hour
12 hour
18 hour
24 hour
30 hour
36 hour
42 hour
48 hour
54 hour
60 hour
66 hour
72 hour
ICU admission
SOFA score sequence

**Figure S20. An example: the illustration of patient's SOFA trajectory.**
